# Using DNA to predict behaviour problems from preschool to adulthood

**DOI:** 10.1101/2021.02.15.21251308

**Authors:** Agnieszka Gidziela, Kaili Rimfeld, Margherita Malanchini, Andrea G. Allegrini, Andrew McMillan, Saskia Selzam, Angelica Ronald, Essi Viding, Sophie von Stumm, Thalia C. Eley, Robert Plomin

## Abstract

**Background:** One goal of the DNA revolution is to predict problems in order to prevent them. We tested here if the prediction of behaviour problems from genome-wide polygenic scores (GPS) can be improved by creating composites across ages and across raters and by using a multi-GPS approach that includes GPS for adult psychiatric disorders as well as for childhood behaviour problems.

**Method:** Our sample included 3,065 genotyped unrelated individuals from the Twins Early Development Study who were assessed longitudinally for hyperactivity, conduct, emotional problems and peer problems as rated by parents, teachers and children themselves. GPS created from 15 genome-wide association studies were used separately and jointly to test the prediction of behaviour problems composites (general behaviour problems, externalizing and internalizing) across ages (from age 2 to age 21) and across raters in penalized regression models. Based on the regression weights, we created multi-trait GPS reflecting the best prediction of behaviour problems. We compared GPS prediction to twin heritability using the same sample and measures.

**Results:** Multi-GPS prediction of behaviour problems increased from less than 2% of the variance for observed traits to up to 6% for cross-age and cross-rater composites. Twin study estimates of heritability mirrored patterns of multi-GPS prediction as they increased from less than 40% to up to 83%.

**Conclusions:** The ability of GPS to predict behaviour problems can be improved by using multiple GPS, cross-age composites and cross-rater composites, although the effect sizes remain modest, up to 6%. Our results can be used in any genotyped sample to create multi-trait GPS predictors of behaviour problems that will be more predictive than polygenic scores based on a single age, rater or GPS.

**Key points:**

- Genome-wide polygenic scores (GPS) can be used to predict behaviour problems in childhood, but the effect sizes are generally less than 3.5%.
- DNA-based prediction models of achieve greater accuracy if holistic approaches are employed, that is cross-trait, longitudinal and trans-situational approaches.
- The prediction of childhood behaviour problems can be improved by using multiple GPS to predict composites that aggregate behaviour problems across ages and across raters.
- Our results yield weights that can be applied to GPS in any study to create multi-trait GPS predictors of behaviour problems based on cross-age and cross-rater composites.
- As compared to individuals in the lowest multi-trait GPS decile, nearly three times as many individuals in the highest internalizing multi-trait GPS decile were diagnosed with anxiety disorder and 25% more individuals in the highest general behaviour problems and externalizing multi-trait GPS deciles have taken medication for mental health.

---

Because all behaviour problems in childhood show moderate genetic influence (Cheesman et al., 2017), a next step in genetic research is to find inherited DNA variants responsible for their heritability. The ability to predict behaviour problems from DNA will facilitate research on topics such as how genetic risk unfolds developmentally, gene-environment interaction and correlation, and multivariate issues of genetic heterogeneity and co-morbidity. It will also advance clinical work by identifying problems on the basis of causes rather than symptoms, by moving away from diagnoses towards dimensions, by switching from one-size-fit-all treatments to individually tailored treatments, and by focusing on prevention rather than treatment (Plomin, 2019).

Genome-wide association (GWA) studies identify DNA variants such as single-nucleotide polymorphisms (SNPs) that are significantly associated with complex traits and common disorders (Visscher et al., 2017). Individual SNP associations have small effect sizes each, but thousands of SNP associations can be aggregated in genome-wide polygenic scores (GPS) to predict considerably more variance (GPS heritability, aka GPS prediction) for some traits (Martin, Daly, Robinson, Hyman & Neale, 2019).

The most predictive GPS for behavioural traits have been reported for educational achievement (Lee et al., 2018) and general cognitive ability (Savage et al., 2018), with GPS heritabilities up to 16% and 11%, respectively (Allegrini et al., 2019). However, despite substantial twin heritability (a mean of 60%; Cheesman et al., 2017), GPS heritabilities are modest for childhood behaviour problems such as autism spectrum disorder (2.5%; Grove et al., 2019) and ADHD (3.3%; de Bode et al., 2020). In a recent study, GPS prediction of childhood ADHD symptoms, internalizing and social problems was reported to be much lower for adult-based GPS of major depression (0.2%), neuroticism (0.1%), insomnia (0.05%) and subjective wellbeing (0.06%) (Akingbuwa et al., 2020). A recent GWA study of childhood and adolescence internalizing symptoms predicted 0.4% of the variance in internalizing at age 7 and 0.03% at ages 13-18 (Jami et. al., 2020). GPS will become more predictive as GWA sample sizes increase and as whole-genome sequencing identifies all DNA variants, rare as well as common, that contribute to heritability (Visscher et al, 2017).

Using existing GPS, we explored ways to increase the prediction of childhood behaviour problems from DNA. Research suggests that using multiple GPS in a multivariate framework can improve prediction (Allegrini et al., 2019, 2020a; Krapohl et al., 2018). To test the hypothesis that the multi-GPS approach will yield greater GPS heritability than the single-GPS approach, we assessed the joint prediction of 15 GPS in penalised regression models with out-of-sample evaluation of prediction accuracy (multi-GPS) (Zou & Hastie, 2005). In addition to GPS for childhood behaviour problems (ADHD; autism spectrum disorder), we included GPS derived from the much larger GWA studies of adult psychiatric disorders such as schizophrenia, bipolar disorder and major depressive disorder and traits such as neuroticism, well-being and risk-taking as they have been shown to predict a variety of childhood phenotypes, including psychopathology (Allegrini et al., 2020b) and behaviour problems (Akingbuwa et al., 2020).

In both phenotypic and DNA-based analyses of behaviour problems, a general factor of psychopathology has been observed that is known as a ‘p-factor’ or ‘p’ (Allergini et al., 2020b; Caspi et al., 2014), suggesting that diverse behaviour problems share common genetic influences. Accordingly, we created latent composites of general behaviour problems (BPp, externalizing and internalizing) and used the multi-GPS approach to test two other hypotheses to improve GPS prediction.

First, because age-to-age stability is largely driven genetically (Nivard et al., 2015; Plomin et al., 2019), we hypothesised that longitudinal composites of behaviour problems composites would yield greater GPS heritability than age-specific observed variables, as suggested by previous genomic research (Cheesman et al., 2018).

Second, building on the assumption that behaviour problems that emerge across situations are more heritable than situation-specific problems, we hypothesised that GPS heritability is greater for behaviour problems composites across raters such as parents, teachers and children themselves who see behaviour problems in different settings than behaviour problems assessed only by one rater (Bartels et al., 2004; Cheesman et al., 2018).

We tested these hypotheses in a sample of 3,065 unrelated individuals from the Twins Early Development Study (TEDS; Rimfeld et al., 2019) for whom we had genotypes and ratings of behaviour problems from early childhood to early adulthood for parents, teachers and the children themselves from age 2 to 21. Because these unrelated individuals were members of twin pairs, we included their co-twins in analyses to estimate heritability using the twin method, testing the hypotheses that cross-age and cross-rater composites increase twin heritability, mirroring the patterns of GPS heritability.

## Methods

Our hypotheses and analyses were preregistered in Open Science Framework (OSF) (https://osf.io/27tpj/) prior to accessing the data. Please see **Supporting information 1** for details. Scripts will be made available on the OSF website.

### Participants

Our sample consists of twins born in England and Wales between 1994 and 1996 enrolled in the Twins Early Development Study (TEDS; for a detailed description of the sample, please refer to the **Supporting information 2** and Rimfeld et al., 2019).

In the current study we investigated heritability of behaviour problems, using data collected when the twins were aged approximately 2, 3, 4, 7, 9, 12, 16 and 21 years old. The sample selected for construction of composites included twins who had at least half of the data on behaviour problems complete across ages and raters. Patterns of missing data were addressed using the full information maximum likelihood. This resulted in a sample of 4,778 twin pairs.

DNA has been genotyped for a subsample of 7,026 unrelated individuals from TEDS (i.e., one twin per pair), out of which 3,065 individuals were included in the present study, which provides a sample size adequate to detect a correlation of 0.10 with more than 99% power (Hulley, Cummings, Browner, Grady & Newman, 2013). For details on sample sizes per composite, please refer to **Supporting table S1**.

Genotyping took place on two different genotyping platforms (AffymetrixGeneChip 6.0 and Illumina HumanOmniExpressExome-8v1.2) in two separate waves. For a detailed genotyping protocol, see Selzam et al. (2018) and Rimfeld et al. (2019).

### Measures

#### Polygenic scores

Our methods for obtaining DNA, genotyping, quality control and constructing polygenic scores have been described previously (Rimfeld et al., 2019; Selzam et al., 2018). In the present analyses, we included 15 genome-wide polygenic scores (GPS) of behaviour problems and psychopathology, derived from the most powerful genome-wide association (GWA) studies, which were used in our previous research (Allegrini et al., 2019; Allegrini et al., 2020a). For the list of polygenic scores, please refer to **Supporting information 3**.

#### Behaviour problems

We assessed hyperactivity, conduct, emotional problems and peer problems from early childhood to early adulthood as rated by parents, teachers and the twins themselves. The Preschool Behaviour Questionnaire (PBQ; Behar, 1997) was used to rate hyperactivity, conduct and emotional problems at ages 2 and 3. At ages 4, 7, 9, 12, 16 and 21, the Strengths and Difficulties Questionnaire (SDQ; Goodman, 1997) assessed peer problems in addition to hyperactivity, conduct and emotional problems. For a description of measure administration and scoring, and an illustration of the four behaviour problems domains across development, please refer to **Supporting information 4**. We also assessed mental health outcomes reported by the twins at age 21, such as mental health diagnoses and whether they have ever taken a medication for mental health.

### Composites

Composites across ages and raters (**Figure 1**) were constructed using the hierarchical latent factor model, where the two first-order factors (externalizing and internalizing) loaded on a second-order factor of BPp. The hierarchical modelling was conducted using confirmatory factor analysis, based upon the results of exploratory factor analyses. For details on the exploratory and confirmatory factor analyses and composite construction, please refer to **Supporting information 5 and 6**.

**Figure 1.**
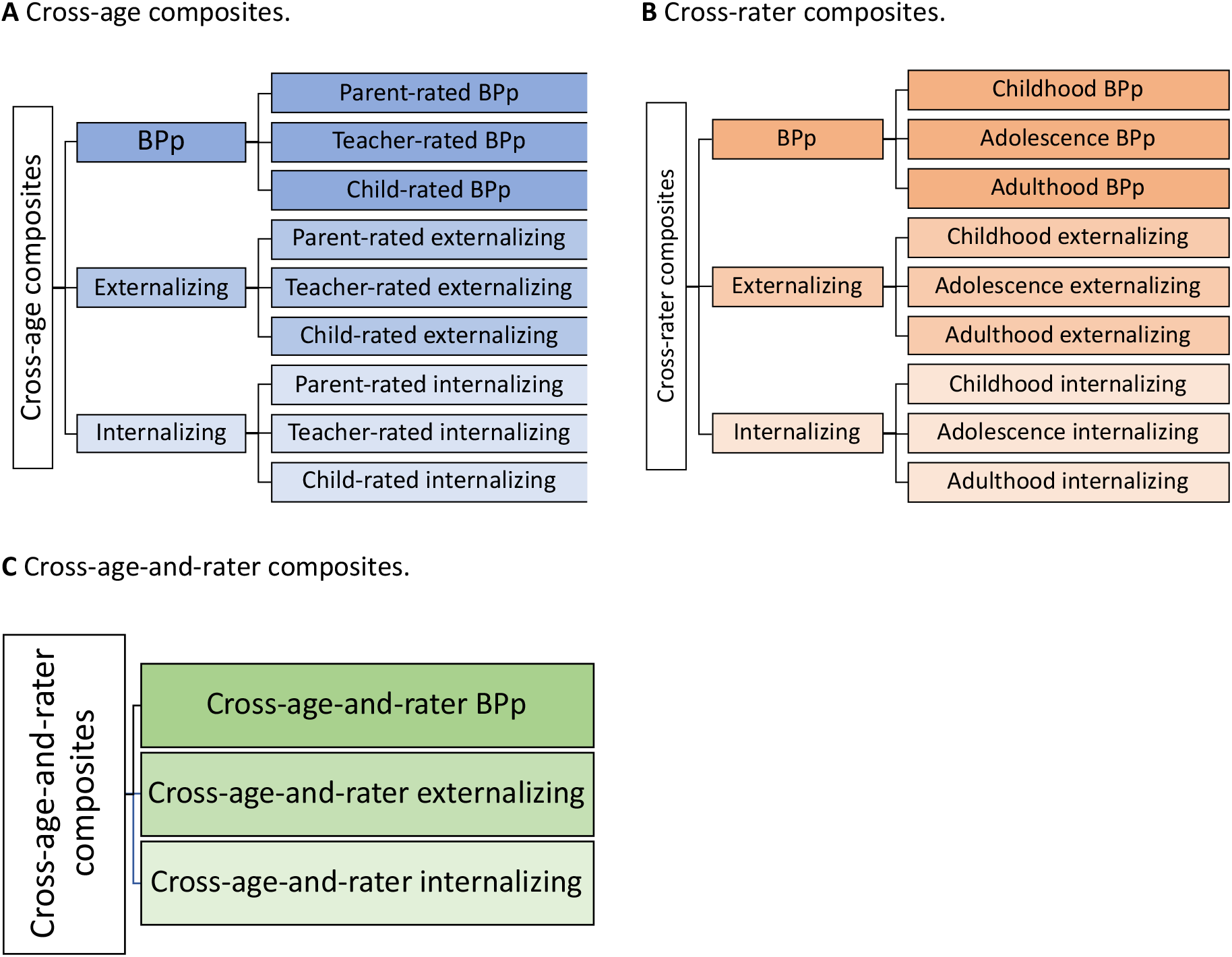
Summary of the construction of the cross-age, cross-rater and cross-age-and-rater composites. *Note*. This figure illustrates the components of the cross-age and cross-rater composites; it is not the hierarchical model used to create composites.

Using hierarchical confirmatory factor analysis, we constructed cross-age and cross-rater composites of BPp, externalizing and internalizing. We created cross-age composites from age 2 to 21 separately for each rater, which yielded nine cross-age composites (three rater-specific composites of BPp, three rater-specific composites of externalizing, three rater-specific composites of internalizing). Cross-rater composites were constructed separately in childhood (ages 2-9), adolescence (ages 12 and 16) and early adulthood (age 21), which yielded nine cross-rater composites (i.e., three age-specific composites each of BPp, externalizing and internalizing). The construction of the cross-age and cross-rater composites is summarised in **Figure 1A and 1B**, respectively. Phenotypic and genetic correlations between the cross-age and cross-rater composites are presented in **Supporting information 7**.

To explore whether simultaneously aggregating cross-age and cross-rater effects improves GPS heritability, we constructed cross-age-and-rater composites of BPp, externalizing and internalizing, using a three-level hierarchical model. In this model, we analysed behaviour problems at all ages (2-21) rated by parent, teacher and child (cross-age approach) to create the first-order factors of cross-age externalizing and internalizing, which were then combined across raters to create the second-order factors of cross-age-and-rater externalizing and internalizing, which subsequently gave rise to the third-order cross-age-and-rater BPp factor (**Figure 1C**). We validated this approach by combining the behaviour problems scales across raters, but separately in childhood, adolescence and adulthood (cross-rater approach) on the first-order factor level, which yielded similar results.

In addition, we created single-trait composites for the four behaviour problems (hyperactivity, conduct, emotional problems and peer problems) in order to compare the effects of single-trait composites to BPp, externalizing and internaliizng composites. Construction and results for the single-trait composites are presented in **Supporting information 8**.

### Analyses

All polygenic scores were regressed on 10 genetic principal components of population structure, genotyping chip and genotyping batch (Allegrini et al., 2019). The standardized residuals from these regressions were used in all downstream analyses.

#### Genome-wide polygenic scores (GPS heritability)

Genome-wide polygenic scores (GPS) are the estimated effects of thousands of genetic variants on a trait and are calculated as a weighted sum of alleles associated with the trait based on summary statistics from genome-wide association studies (Dudbridge, 2013). The GPS were constructed using LD-pred (Vilhjálmsson et al., 2015), with the 1000 Genomes phase 1 sample as a reference for linkage disequilibrium structure. A detailed description of our LD-pred analytic strategy used to calculate GPS has been published (Allegrini et al., 2019). We report results for GPS created at p-value threshold of 1.0, although results for GPS p-value thresholds of 0.3 and 0.01 are presented in **Supporting tables S2-S5**. In addition, we reported the GPS results separately for males and females (**Supporting table S4**).

We estimated the joint prediction of the 15 GPS (multi-GPS heritability) in a penalized regression elastic net model with out-of-sample evaluation of prediction accuracy. For details on the elastic net regularization analytic procedure, please refer to **Supporting information 9** and Allegrini et al. (2020a).

#### Multi-GPS effects

To investigate whether a multi-GPS approach improved prediction as compared to a single-GPS approach, we compared the joint prediction of behaviour problems by the 15 GPS (multi-GPS heritability) to individual predictions yielded by each of the 15 GPS alone (single-GPS heritability). The multi-GPS heritability was estimated in elastic net regularization models and multiple regression models (using adjusted R^2^). Single-GPS heritability was estimated using squared correlations (r^2^) between each of the 15 GPS and composites.

#### Compositing effects

We compared the multi-GPS heritability for the composites to the mean multi-GPS heritability for the individual constituent behaviour problem traits that comprise these composites (that is, the age and rater-specific traits which we will refer to as observed traits) (**Supporting tables S2 and S5**). For example, the multi-GPS heritability of the cross-age parent-rated externalizing composite was compared to the mean of multi-GPS heritabilities of parent-rated hyperactivity and conduct scales across ages 2 to 21. Although the focus of this paper is to present a broad picture of the effect sizes, rather than formally testing for significant differences, in order to present the 95% confidence intervals of the estimates that index significance of differences, we also used a meta-analytic approach (**Supporting information 10**).

#### Analysis of extremes

In addition to continuous analyses, we investigated the ability of GPS to predict differences in behaviour problems at the decile extremes of the multi-trait GPS, using the cross-age-and-rater composites of BPp, externalizing and internalizing as an example. We created multi-trait GPS scores based on the individual predictor GPS coefficients from the elastic net regularization models (**Supporting table S9**), using the following formula:

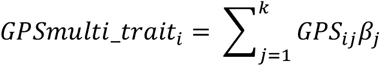

where *GPS multi_trait* is the multi-trait GPS for individual *i* in the full sample, *j ∈* {1, 2, …, 15} and denotes the GPS value for the k GPS for individual *i* and ß indicates the elastic net coefficient of the association between the *jth* predictor GPS and the composite that was learnt in the training set (see **Supporting information 9** for details).

After assigning multi-trait GPS scores to each individual for BPp, externalizing and internalizing, we divided the sample into deciles and compared their mean phenotypic scores for BPp, externalizing and internalizing as well as for other mental health outcomes.

#### Twin heritability

We compared the multi-GPS heritability results to heritability results from twin analyses (**Supporting tables S6** and **S8**). The classical univariate twin design was employed to estimate broad heritability (additive and non-additive genetic variance) for individual behaviour problems as compared to composites. We performed twin analyses using OpenMx 2.0 for R (Neale et al., 2016; R Core Team, 2020). Additionally, we report the univariate twin model estimates separately for males and females (**Supporting table S7**).

In order to investigate the impact of compositing on twin heritability, we contrasted twin heritability estimates for composites to the mean twin heritabilities for the observed traits. Significance of these differences was assessed using a meta-analytic approach (**Supporting information 10**).

## Results

### Multi-GPS heritability: cross-age and cross-rater composites

Results of the multi-GPS prediction with elastic net regularization are shown in **Figure 2** for cross-age composites (**Figure 2A**) and cross-rater composites (**Figure 2B**). As shown in **Supporting information 7**, cross-age and cross-rater composites were substantially correlated phenotypically and genetically.

**Figure 2.**
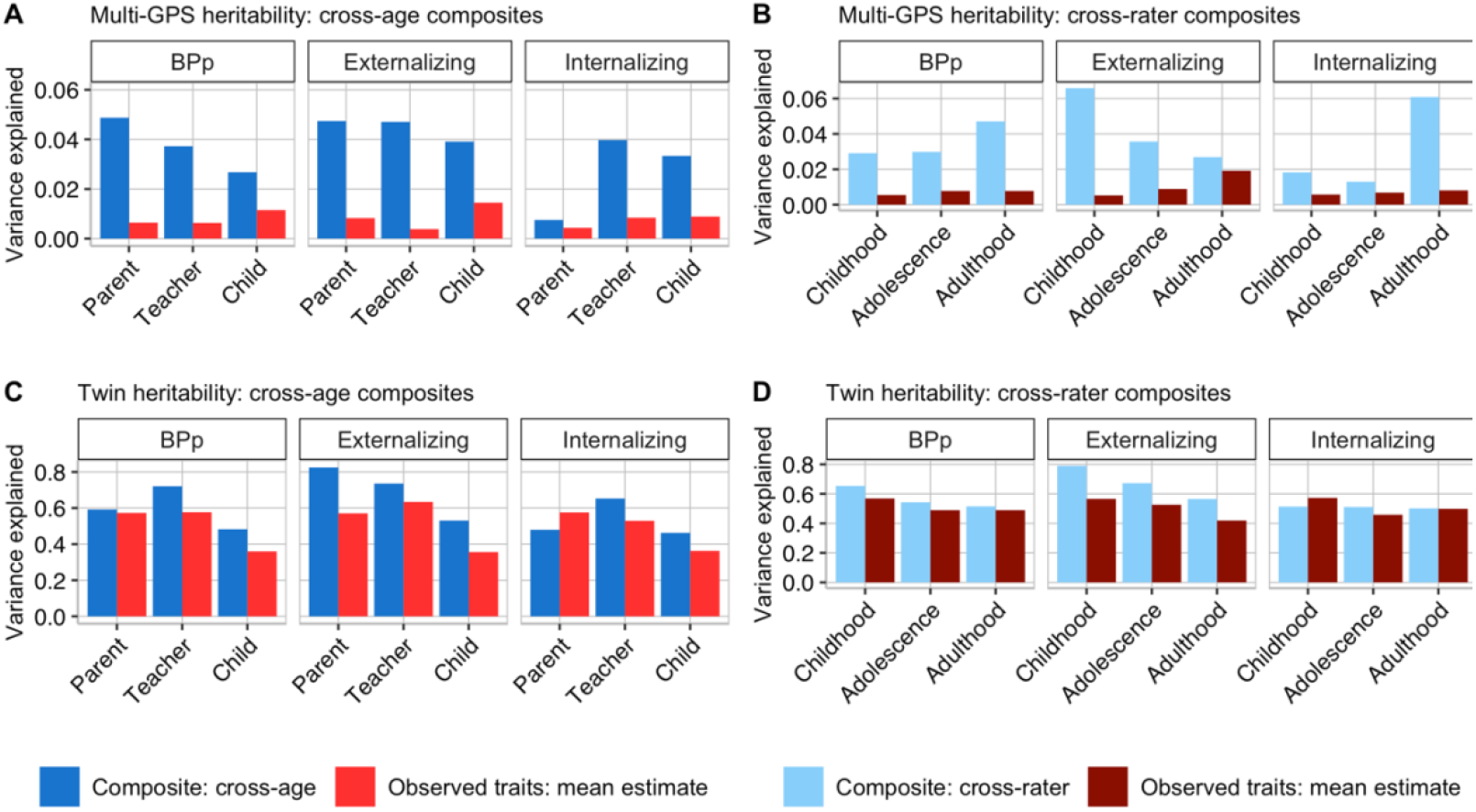
Multi-GPS and twin heritability of cross-age composites and cross-rater composites, compared to the mean multi-GPS and twin heritability of observed traits.

Compositing across ages increased multi-GPS heritability as compared to the mean multi-GPS heritability of observed traits for BPp, externalizing and internalizing (**Figure 2A**). The greatest cross-age effect was found for parent-rated BPp, with the multi-GPS predicting 4.9% of the variance, as compared to the mean estimate of 0.6% when considering observed behaviour problems. Parent-rated multi-GPS heritabilities were 4.7% vs 0.8% for externalizing, but only 0.7% vs 0.4% for internalizing. For teacher ratings, the multi-GPS heritabilities were 3.7% vs 0.6% for BPp, 4.7% vs 0.4% for externalizing problems and 4% vs 0.8% for internalizing problems. Finally, for child ratings, the multi-GPS heritabilities were 2.7% vs 1.2% for BPp, 3.9% vs 1.4% for externalizing problems and 3.3% vs 0.9% for internalizing problems.

Compositing across raters also increased multi-GPS heritability in childhood, adolescence and adulthood, as shown in **Figure 2B**. For BPp, multi-GPS heritability for cross-rater composites was 2.9% as compared to the mean of 0.5% for the observed traits in childhood, 3.0% vs 0.8% in adolescence and 4.7% vs 0.8% in adulthood. For externalizing problems, multi-GPS heritabilities were 6.6% vs 0.5% in childhood, 3.6% vs 0.9% in adolescence and 2.7% vs 1.9% in adulthood. For internalizing problems, multi-GPS heritabilities were 1.8% vs 0.6% in childhood, 1.3% vs 0.7% in adolescence and 6.0% vs 0.8% in adulthood. The greatest cross-rater effect was found for externalizing problems in childhood, with the multi-GPS prediction of 6.6% as compared to 0.8% for observed traits.

**Figure 3** compares the multi-GPS approach to the single-GPS approach in prediction of cross-age and cross-rater composites. The first row of each of the six panels in **Figure 3** repeats the results in **Figure 2** showing the multi-GPS prediction using elastic net regularization for cross-age composites (**Figure 3A**) and cross-rater composites (**Figure 3B**). The second row shows that in most cases the elastic net regularization performed better than adjusted R^2^ from simple multiple regressions. The rest of each panel shows the variance explained (correlation squared) by each of the 15 GPS alone.

**Figure 3.**
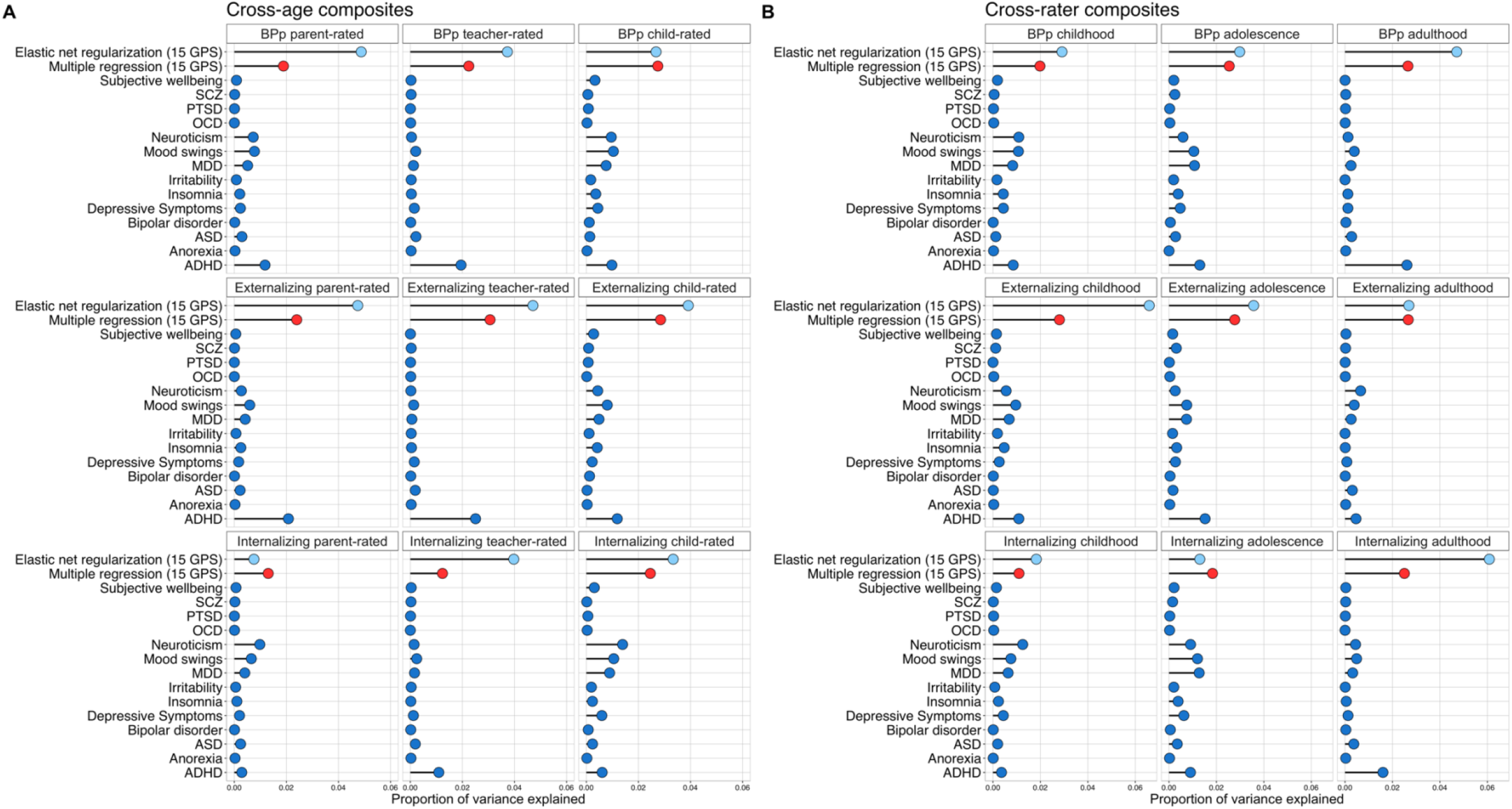
Multi-GPS prediction as compared to single-GPS prediction of cross-age composites and cross-rater composites.

For BPp and externalizing problems, the ADHD GPS was the most predictive GPS for cross-age and cross-rater composites, predicting up to 2.6% of the variance in the cross-rater composite of adulthood p and 2.5% of the variance in the cross-age composite of teacher-rated externalizing. Other than the ADHD GPS, none of the individual GPS predicted more than 1.5% of the variance. For internalizing problems, the most predictive GPS was the neuroticism GPS which predicted up to 1.4% of the variance in cross-age composites of child-rated internalizing and cross-rater childhood internalizing and 1.3% in cross-rater composites of childhood and adolescence internalizing.

### Twin heritability: cross-age and cross-rater composites

**Figure 2C and 2D** summarises twin heritability estimates for cross-age composites (**Figure 2C**) and cross-rater composites (**Figure 2D**) as compared to the mean estimates of twin heritability of the observed traits. In general, cross-age and cross-rater composites yielded greater twin heritability estimates than the observed traits.

The average heritability for cross-age composites was 61% as compared to 50% for the observed traits (**Figure 2C)**. The largest difference was found for parent-rated externalizing problems (82% vs 57%). The pattern of cross-age effects for twin heritability largely mirrored the multi-GPS heritability results, with the notable exception that twin heritability showed no cross-age effect for parent ratings of BPp, whereas this was one of the largest cross-age effects for multi-GPS heritability.

For cross-rater composites, the average heritability was 58% as compared to 51% for the observed traits (**Figure 2D**). The average cross-rater effect across the three ages was strongest for externalizing problems (68% vs 55%), weaker for BPp (57% vs 54%) and absent for internalizing problems (51% vs 53%). The strongest cross-rater effect was observed for externalizing problems in childhood (79% vs 57%), which is consistent with the multi-GPS results. Similar to multi-GPS heritability, twin heritability for cross-rater externalizing problems decreased from childhood (79%) to adolescence (67%) to adulthood (57%).

### Aggregated cross-age-and-rater effects

**Figure 4** compares the multi-GPS heritability and twin heritability obtained for the combined cross-age-and-rater composites. Multi-GPS heritabilities of the cross-age-and-rater composites were similar to multi-GPS heritability of the cross-age composites (**Figure 4A**) and cross-rater composites (**Figure 4B**). Combining traits across ages and raters did not significantly improve GPS heritability. The variance explained by the GPS in the combined cross-age-and-rater composites (3.3%) was similar to the average prediction yielded by cross-age and cross-rater composites (3.6%).

The twin analyses also showed that the benefits of cross-age and cross-rater compositing are not additive (**Figure 4C** and **4D**, respectively). The twin heritability for cross-age-and-rater composites (63%) was similar to the mean twin heritability yielded by cross-age and cross-rater composites (60%).

**Figure 4.**
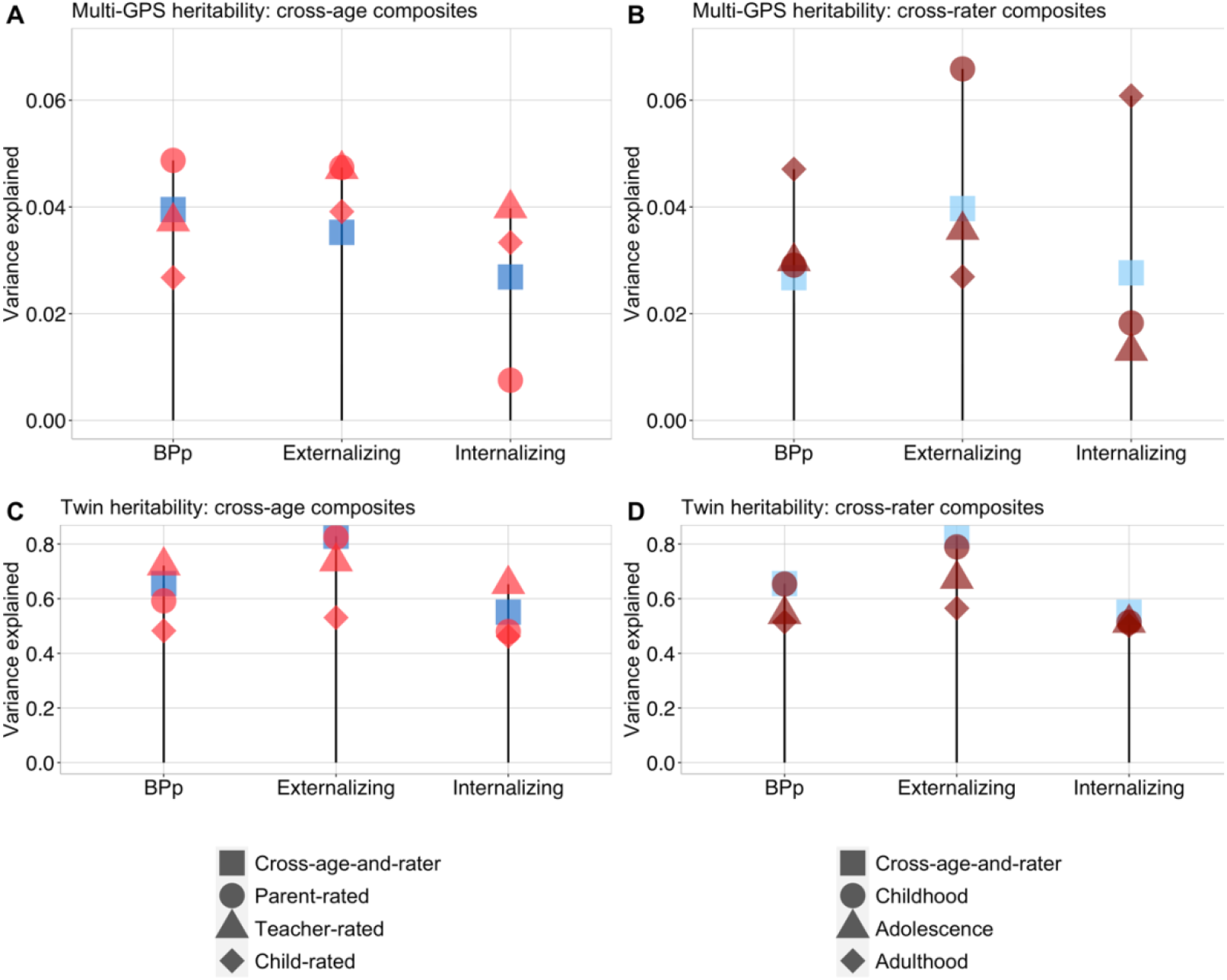
Multi-GPS heritability and twin heritability of cross-age-and-rater composites as compared to cross-age composites and to cross-rater composites. *Note*. Dark blue squares signifies the cross-age-and-rater composites constructed using the cross-age approach; lights blue squares signifies the cross-age-and-rater composites constructed using the cross-rater approach (see **Methods**).

### Analysis of multi-trait GPS decile extremes

Multi-trait GPS scores for BPp, externalizing and internalizing were created for each individual as explained earlier. We used these multi-trait GPS scores to divide the sample into deciles. **Figure 5** shows box plots, presenting the z-standardized scores for cross-age-and-rater BPp, externalizing and internalizing as a function of the multi-trait GPS deciles. Mean behaviour problems increase linearly from the lowest to the highest GPS deciles, with a scatterplot of scores as expected from the modest correlations between the multi-trait GPS and BPp (r = 0.19), externalizing (r = 0.20) and internalizing (r = 0.16). At the lowest and highest decile extremes, the differences are substantial: the mean standard score difference between the lowest and highest GPS deciles is 0.61 for BPp, 0.67 for externalizing and 0.51 for internalizing.

**Figure 5.**
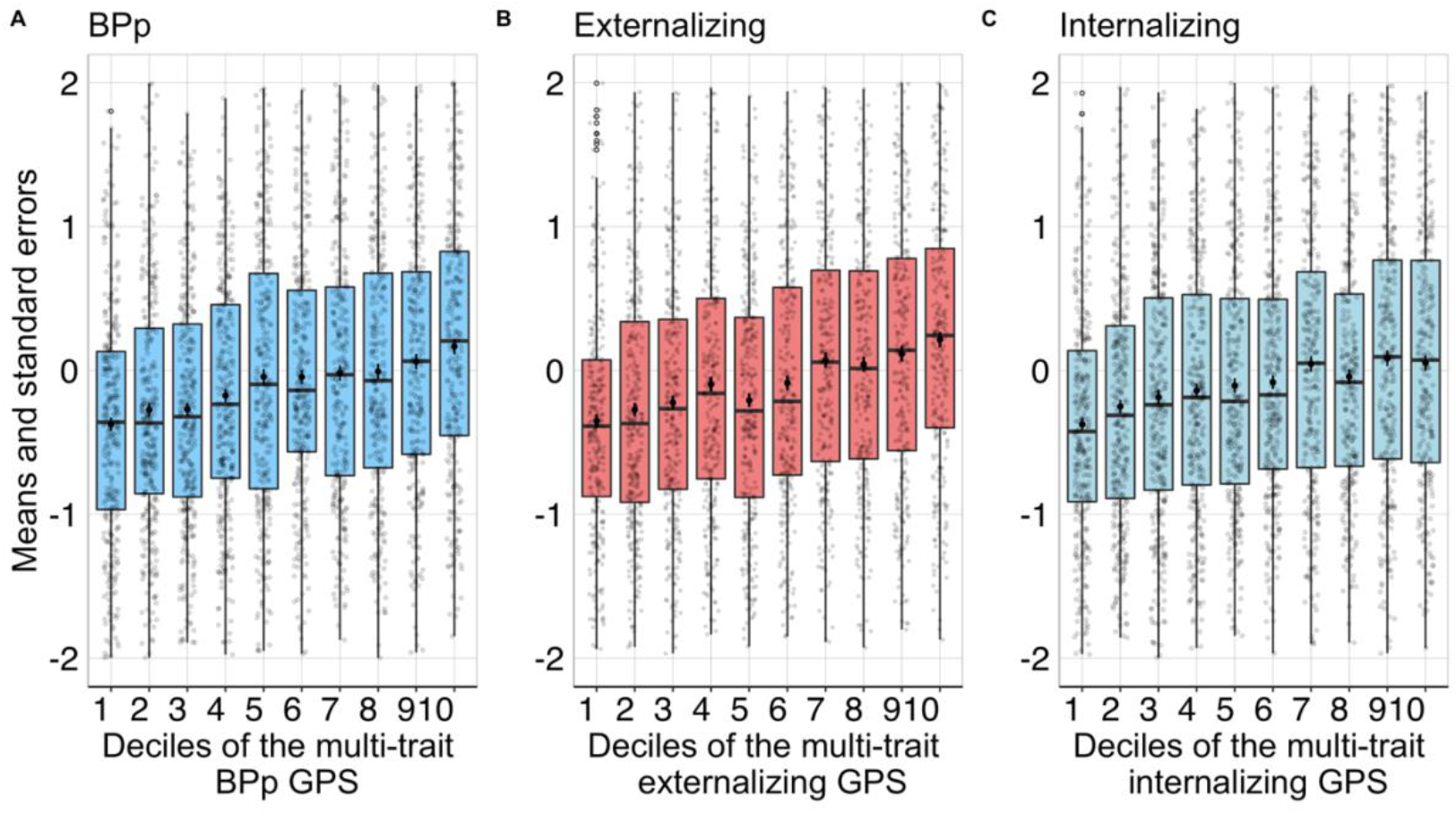
Box plots showing z-standardized means and distributions of cross-age-and-rater multi-trait GPS scores for BPp, externalizing and internalizing (**C**). *Note*. The boxes enclose 50% of the distribution of each GPS decile. Horizontal lines in boxes indicate the median values. Dots and error bars in boxes indicate means and standard errors. Vertical lines outside the boxes indicate the normal distribution of GPS deciles. Point contours indicate outliers.

Differences between the lowest and highest deciles were reflected in mental health outcomes. For example, 15% of individuals in the lowest multi-trait GPS decile for BPp and 15% in the lowest multi-trait GPS decile for externalizing have taken medication for mental health, compared to 20% in the highest BPp and 21% in the highest externalizing decile, although these differences are not statistically significant (odds ratio and 95% confidence intervals: 1.47 (0.84, 2.57) for p and 1.01 (0.58, 1.78) for externalizing). For the multi-trait internalizing GPS, 12% of individuals in the lowest decile have been diagnosed with depression, compared to 19% in the highest decile (odds ratio and 95% confidence intervals: 1.82 (1.03, 3.24)), while 7% of individuals in the lowest decile have been diagnosed with anxiety disorder, compared to 19% in the highest decile (odds ratio and 95% confidence intervals: 2.90 (1.53, 5.75)).

## Discussion

Our findings indicate that a multi-GPS approach using cross-age and cross-rater composites doubles the prediction estimates for general behaviour problems, however with the modest effect sizes there is still a long way to go from prediction to prevention. These results are bolstered by twin analyses showing, although to a lesser extent, increased heritability for cross-age and cross-rater composites. The twin design can be viewed as the prediction ceiling because it assesses the effect of all inherited DNA differences, not just SNPs shown to be associated with behavioural problems.

The multi-GPS weights for our cross-age-and-rater composites that simultaneously composite across age and across raters provide the best genetic estimates currently available for children’s BPp, externalizing and internalizing problems (**Supporting table S9**). These multi-GPS weights will be useful as genetic predictors of behaviour problems for any sample with DNA regardless of whether any behaviour problems data are available. Just as the 15 GPS can be created from DNA for any sample, our sets of multi-GPS weights can be used to create the best possible genetic estimates of BPp, externalizing and internalizing based on cross-age, cross-rater and cross-age-and-rater composites. These multi-trait GPS will facilitate developmental, multivariate and gene-environment interplay research because they are more predictive of behaviour problems than polygenic scores based on a single age, rater or trait. Our goal of increasing GPS heritability led us to focus on compositing across ages, raters and traits, which should not be seen to denigrate the continued search for specific genetics effects for each age, rater or trait. Although we present results for the cross-age-and-rater multi-trait GPS for conceptual consistency, we report weights for all of the composites that will allow researchers to construct developmental stage-specific and rater-specific multi-trait GPS.

In order to condense the results, we focused on the second-order factors of externalizing and internalizing and a third-order factor representing BPp. However, we also present multi-GPS weights for the single-trait cross-age and cross-rater composites of hyperactivity, conduct, emotional and peer problems (**Supporting table S9**). Although these traits generally showed increased GPS and twin heritability for cross-age and cross-rater composites, results for these trait-specific factors are subject to more measurement error, hence the results are less consistent than for the general factors representing BPp, externalizing and internalizing problems.

More research is needed to identify the mechanisms by which compositing increases GPS prediction. We had assumed that compositing across ages captures new genetic effects that come on board at later ages and that compositing across raters captures trans-situational genetic effects in the home for parent ratings and in school for teacher ratings. However, if different mechanisms are responsible for increasing GPS prediction for cross-age and cross-rater composites, we would expect that the effects of compositing across ages and across raters would be additive. However, the combined cross-age-and-rater composites do not show increased GPS heritability nor increased twin heritability as compared to the cross-age and cross-rater composites. Notably, we found that cross-age effects differ depending on rater, and, similarly, cross-rater effects depend on developmental stage. These interactions might explain in part why cross-age and cross-rater effects do not add up. However, going against this interaction hypothesis is the strong phenotypic overlap (∼0.60) and genetic overlap (∼0.65) between cross-age and cross-rater effects (**Supporting information 7**), which suggests that the same mechanisms are responsible for increasing heritability for cross-age and cross-rater composites. A likely candidate is increased reliability, which could increase heritability for both cross-age and cross-rater composites. However, this reliability hypothesis requires the added assumption that compositing either across ages or across raters reaches a ceiling of reliability, so that there is no additional increase in heritability for cross-age-and-rater composites.

Although compositing doubles the predictive power of GPS, the effect sizes remain modest, less than 6%. Nonetheless, we show (**Figure 5**) that multi-trait GPS with these effect sizes result in sizeable (*Cohen’s d* ∼ .5) mean differences in behaviour problems at the multi-trait GPS extremes as well as doubled odds ratios for mental health outcomes. Increasing the power of GPS to predict behaviour problems ultimately depends on bigger GWA studies. Our results imply that GWA studies can increase their power to detect effects by conducting GWA analyses using cross-age or cross-rater composites.

Our results are limited to existing GWA studies and will need to be updated as new GWA studies are reported. A more specific limitation is that we focused on the 15 most powerful GWA of psychopathology regardless of whether the GWA analysis targeted childhood disorders (autism spectrum disorder and ADHD) or disorders in adulthood (e.g., schizophrenia and depression). It is reasonable to expect that GWA studies targeted on childhood disorders will add disproportionately to the multi-GPS prediction of childhood behaviour problems. Supporting this expectation is our finding that the ADHD GPS was by far the strongest single GPS predictor of behaviour problems, especially for parent and teacher ratings of the BPp factor and externalizing problems (**Figure 3A**). Nonetheless, multi-GPS predicted twice as much variance, with adult-based GPS for neuroticism, mood swings, and major depressive disorder contributing to the prediction from the ADHD GPS. Our approach is atheoretical and empirical in the sense that we would include any GPS, child-based or adult-based, that adds to the multi-GPS prediction of behaviour problems.

There is special value in focusing on GPS derived from adult-based GWA studies because they predict adult psychiatric disorders from childhood regardless of their associations with childhood behaviour problems. We chose not to do this at this time because our aim was to increase the DNA prediction of childhood behaviour problems, and we show that multi-GPS limited to extant adult-based GWA studies are weak predictors of childhood behaviour problems.

## Data Availability

Scripts are available on the OSF site.

https://osf.io/27tpj/

## Acknowledgements

We gratefully acknowledge the on-going contribution of the participants in the Twins Early Development Study (TEDS) and their families. TEDS is supported by a programme grant to R.P. from the UK Medical Research Council (MR/M021475/1 and previously G0901245), with additional support from the US National Institutes of Health (AG046938) and the European Commission (602768; 295366).

## Abbreviations

GPS: genome-wide polygenic scores

