## Supplementary material for "Using DNA to predict behaviour problems from preschool to adulthood": SI.docx

| **Table of contents** | |
| --- | --- |
| **Supporting information** | |
| *Supporting information 1.* | *OSF statement.* |
| *Supporting information 2.* | Sample description. |
| *Supporting information 3.* | Polygenic scores. |
| *Supporting information 4.* | Additional information about the behaviour problems measures: SDQ and PBQ. |
| *Supporting information 5.* | Exploratory factor analysis (EFA). |
| *Supporting information 6.* | Confirmatory factor analysis (CFA). |
| *Supporting information 7.* | Phenotypic and genetic correlations between cross-age and cross-rater composites. |
| *Supporting information 8.* | Construction and results for the single-trait composites. |
| *Supporting information 9.* | Analytic procedure of the elastic net regularization. |
| *Supporting information 10.* | The meta-analytic approach to comparing multi-GPS heritability between composites and constituent traits. |
| **Supporting figures** | |
| *Supporting figure S1.* | Observed variables at each age. |
| *Supporting figure S2.* | Exploratory factor analysis models. |
| *Supporting figure S3.* | Hierarchical and bifactor models. |
| *Supporting figure S4.* | Correlations between hierarchical and bifactor composites. |
| *Supporting figure S5.* | Phenotypic and genetic correlations between the cross-age and cross-rater composites. |
| *Supporting figure S6.* | Construction of the single-trait cross-age and single-trait cross-rater composites. |
| *Supporting figure S7.* | Multi-GPS and twin results for the single-trait cross-age and cross-rater composites. |
| *Supporting figure 8.* | Multi-GPS correlation and twin heritability results for cross-age composites and cross-rater composites. |
| *Supporting figure 9.* | Multi-GPS correlation and twin heritability results for single-trait cross-age composites and single-trait cross-rater composites. |
| **Supporting tables** | |
| *Supporting table S1.* | Sample characteristics. |
| *Supporting table S2.* | Model fit indices and predictions of behaviour problems composites  from elastic net regularization. |
| *Supporting table S3.* | Model fit indices and predictions of behaviour problems composites  from multiple regression. |
| *Supporting table S4.* | Model fit indices and predictions of behaviour problems composites  from multiple regression,  for males and females separately. |
| *Supporting table S5.* | Model fit indices and predictions of observed behaviour problems  from elastic net regularization. |
| *Supporting table S6.* | Univariate twin model fitting results for behaviour problems composites. |
| *Supporting table S7.* | Univariate twin model fitting results for behaviour problems composites,  for males and females separately. |
| *Supporting table S8.* | Univariate twin model fitting results for observed behaviour problems. |
| *Supporting table S9.* | Weights for the polygenic scores from elastic net regularization. |
| *Supporting table S10.* | Bivariate twin model fitting results for cross-age and cross-rater composites. |
| *Supporting table S11.* | Representativeness of the selected sample. |
| *Supporting table S12.* | Polygenic scores and sample sizes. |

***Supporting information 1.***

*Hypotheses from our OSF (Open Science Framework) statement* (<https://osf.io/27tpj/>)*.*

Although there are as yet no multi-GPS analyses of behaviour problems from childhood to early adulthood, we base the following OSF-registered hypotheses on varying degrees of evidence from the results of previous genetic and genomic analyses (for example, Cheesman et al., 2018; Allegrini et al., 2020).

*Hypothesis I*

Variance in behaviour problems explained by multivariate GPS analysis (GPS heritability)

a) will be modest, less than 5%.
b) will be greater than GPS heritability for individual GPS.
c) will increase during development as children grow closer to the target age of the mostly adult participants in the genome-wide association (GWA) studies from which the GPS are derived.
d) will be greater for externalizing problems composites than for internalizing problems composites.
e) will be greater for parent and teacher ratings than for self-reports.
f) will be greater for longitudinal and cross-rater composites than for rater-specific measures at each age.
g) will be greater for cross-rater composites than for single-rater composites.
h) will be greater for a general factor of behaviour problems (BPp) than for individual measures of behaviour problems.
i) will be substantially lower than for cognitive and anthropometric traits.
j) will be similar for girls and boys.

*Hypothesis II*

SNP heritability for behaviour problems

a) will be substantially higher than GPS heritability.
b) will be substantially lower than heritability estimated from twin model-fitting analyses.
c) will be substantially lower than for cognitive and anthropometric traits.

*Hypothesis III*

Twin model-fitting heritability for behaviour problems

a) will be similar for parent and teacher ratings and lower for self-reports.
b) will be similar to cognitive traits but lower than anthropometric traits.
c) will reveal nonadditive genetic effects for behaviour problems but not for cognitive and anthropometric traits.

We addressed all these hypotheses in the current paper, apart from hypotheses regarding SNP heritability (II a, b, c) and cognitive/anthropometric traits (I a, III b, III c).

Our hypotheses were pre-registered prior to obtaining the dataset; hence we were not able to estimate the precise sample sizes for composites at that stage. Once we examined sample sizes of the obtained dataset, we did not have sufficient power to detect reliable SNP heritability estimates, according to the genome-wide complex trait analysis (GCTA) power calculator (Hermani & Yang, 2017). The power analysis revealed that with the sample of 3065 individuals we only had 16% of power to detect the SNP heritability of 10% (alpha= 0.05) (Hermani et al., 2017; for computation details refer to Vissher et al., 2014). Having acknowledged our SNP heritability estimates are unreliable due to low power, we have decided not to present them in this paper.

The other theme not covered in the paper but mentioned in the OSF pre-registration is to compare behaviour problems and cognitive and anthropometric traits. Increasing GPS heritability of cognitive/anthropometric traits could only be addressed from the longitudinal and not trans-situational perspective due to cognitive/anthropometric data only being collected on the level of a single-rater. Also, while having 15 polygenic scores available for behaviour problems, only 3 were available for cognitive traits (educational achievement, intelligence and income) and only 2 for anthropometric traits (height and body mass index). In order to overcome these discrepancies and enable comparability, as well as address the *nonadditive hypothesis* (III c), major extensions to the study would be needed, which currently are beyond the scope of this paper, which is already considerably complex and lengthy.

***Supporting information 2*.**

Sample description.

Our sample consists of twins born in England and Wales between 1994 and 1996 enrolled in the Twins Early Development Study (TEDS; Rimfeld et al., 2019). Parents of twins were invited to participate in TEDS when the twins were about one year old. The invitations were sent to families by the UK Office for National Statistics after screening for infant mortality, and 16,810 families expressed interest in taking part.

The TEDS twins have been assessed a dozen times from infancy through early adulthood on a wide range of behavioural, psychological, cognitive and physical measures, including genome-wide genotyping for a sub-sample (Rimfeld et al., 2019). Data collection procedures included questionnaires and tests administered by post, by telephone and online, as described in detail in a recent overview of TEDS (Rimfeld et al., 2019; details can be found in the TEDS data dictionary: <https://www.teds.ac.uk/datadictionary/home.htm>).

The sample of TEDS twins represents a broad swathe of the UK population in terms of ethnicity and socioeconomic status (SES) (Rimfeld et al., 2019), so is our selected sample of 3,065 unrelated twins (**Supporting table S11**). Individuals with severe medical conditions that affected participation were excluded from analyses. Zygosity was assessed using a parent questionnaire of the twins’ physical similarity, which yielded 95% accuracy when compared to DNA tests (Price et al., 2000). For twin pairs with uncertain zygosity based on physical resemblance, DNA was used to establish reliable zygosity estimates (Price et al., 2000).

| **Supporting table S11**. Representativeness of the selected sample. | | | |
| --- | --- | --- | --- |
| Ethnicity and SES | **Selected sample** | **1st Contact sample** | **National equivalents^a,b^** |
| % white | 94.6% | 91.7% | 93% |
| % mother A-levels or higher | 45.3% | 35.5% | 35% |
| % father A-levels or higher | 51.7% | 44.8% | 47% |
| % mother employed | 47.9% | 43.1% | 50% |
| % father employed | 93.9% | 91.6% | 91% |
| Note. ^a^ including cohort of parents with children born in late 1990s and early 2000s;  ^b^ derived from Rimfeld et al. (2019). | | | |

***Supporting information 3.***

Polygenic scores.

The 15 polygenic scores used in the current study and the sample sizes are presented in **Supporting table S12**.

| **Supporting table S12**. Polygenic scores and sample sizes. | |
| --- | --- |
| Polygenic score | **Sample size** |
| Attention deficit hyperactivity disorder (ADHD) (Demontis et al., 2019) | 55374 |
| Anorexia nervosa (Watson et al., 2019) | 14477 |
| Autism spectrum disorder (ASD)  (Grove et al., 2019), | 46350 |
| Bipolar disorder (Stahl et al., 2019) | 51710 |
| Broad depression (Howard et al., 2019) | 331374 |
| Insomnia (Jansen et al., 2019) | 386533 |
| Irritability (Neale Lab, 2017) | 322668 |
| Major depressive disorder (MDD)  (Wray et al., 2018) | 173005 |
| Mood swings (Neale lab, 2017) | 329428 |
| Neuroticism (Luciano et al., 2018) | 329821 |
| Obsessive-compulsive disorder (OCD) (Genetics IOCDF et al., 2018) | 9725 |
| Post-traumatic stress disorder (PTSD) (Duncan et al., 2018) | 9537 |
| Risk-taking (Linnér et al., 2019) | 325821 |
| Schizophrenia (Pardinas et al., 2018) | 105318 |
| Wellbeing (Okbay et al., 2016) | 298420 |

***Supporting information 4.***

Additional information about the behaviour problems measures: SDQ and PBQ.

The SDQ (Strengths and difficulties questionnaire) and PBQ (Preschool behaviour questionnaire) were generally administered by post, in a form of paper questionnaires.

A small proportion of parent-report questionnaires were collected over the phone at age 7 and a larger proportion of parent and self-report questionnaires were collected online at age 21.

The PBQ questionnaire (Behar, 1977) was only administered to parents at ages 2 and 3 and included 18 items (4 items measuring hyperactivity, 8 items measuring conduct problems and 6 items measuring emotional problems). The SDQ questionnaire (Goodman, 1997) was administered to parents (ages 4- 21), teachers (ages 7- 12) and children (ages 9- 21) and included 20 items (5 items per domain, with peer problems being the fourth domain).

Both PBQ and SDQ questionnaires were rated on a three-point Likert scale (Certainly true; Sometimes true; Not true). All items were scored in the direction of problems (for example *Restless, overactive, cannot stay still for long*) or reversed where necessary, so that higher scores suggested more severe behaviour problems. The total hyperactivity, conduct, emotional problems and peer problems scores were computed as the mean of the non-missing items (provided at least half of those items to be complete) multiplied by the number of items within the scale.

The four SDQ domains from age 2 to age 21 are presented in **Supporting figure S1** below.

**Supporting figure S1**. The sunburst plot showing the observed variables at each age.

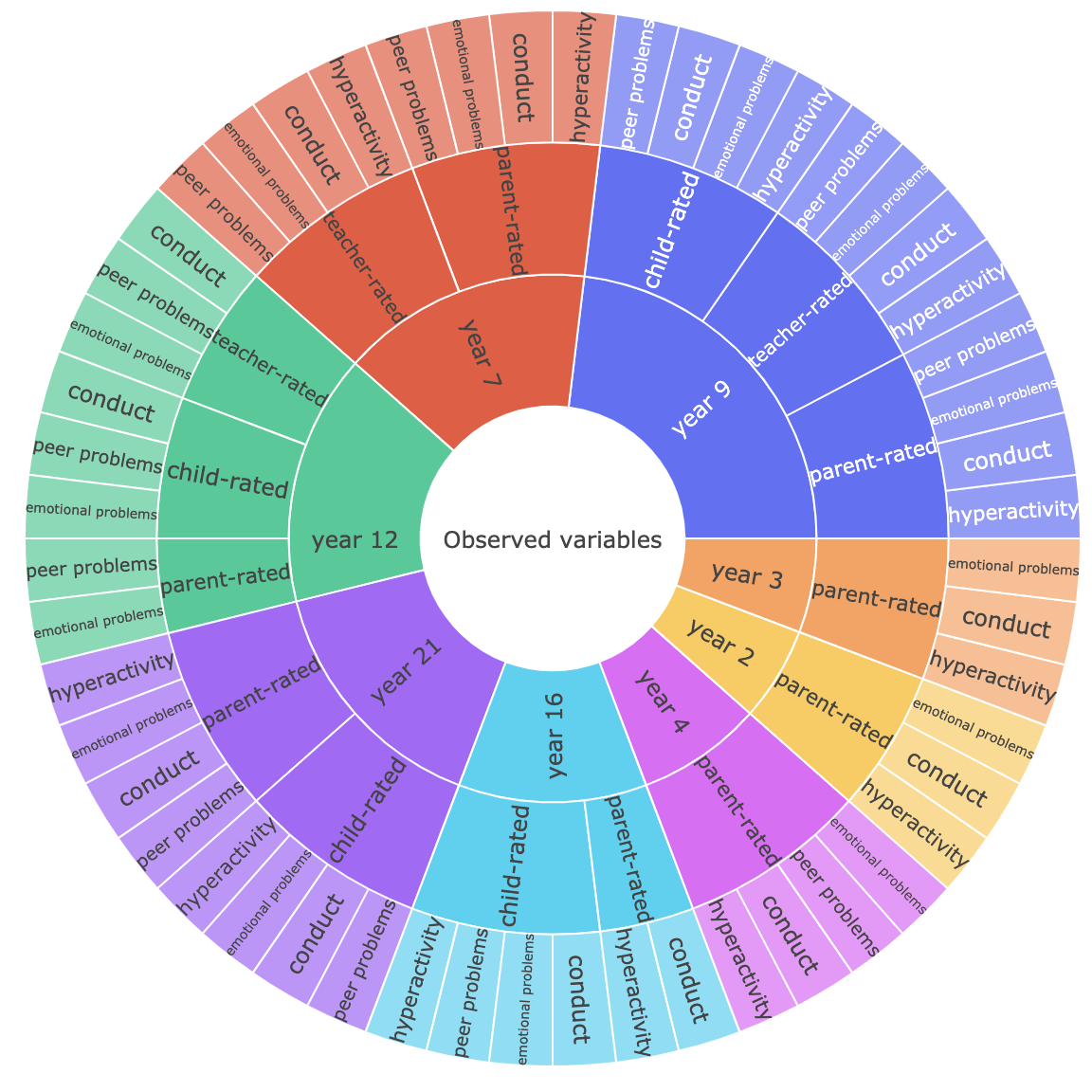

***Supporting information 5*.**

Exploratory factor analysis (EFA).

EFA analyses were conducted in *psych* for R (Revelle, 2020; R Core Team, 2020), after residualizing all variables on age and sex for twin 1 and twin 2 separately (McGue & Bouchard, 1984). To explore the factor structure of behaviour problems in childhood, we performed the EFA of the four SDQ scales (hyperactivity, conduct, emotional problems and peer problems) at age 9, which was selected due to availability of complete data for all scales and for all three raters: parents, teachers and children themselves. The exploratory factor analysis was performed separately for parent (**Supporting figure S2A**), teacher (**Supporting figure S2B**) and child ratings (**Supporting figure S2C**).

EFA suggested a two-factor structure for all three raters. Hyperactivity and conduct problems loaded strongly on factor 1 (henceforth externalizing), and emotional problems and peer problems loaded strongly on factor 2 (henceforth internalizing). The externalizing and internalizing factors were moderately correlated (0.27, 0.56 and 0.63 for parent, teacher and child-rated data respectively), suggesting a general factor of behaviour problems (henceforth BPp).

| **Supporting figure S2**. Exploratory factor analyses for parent, teacher and child-rated data. | | |
| --- | --- | --- |
| **A Parent ratings** | **B Teacher ratings** | **C Child ratings** |
| 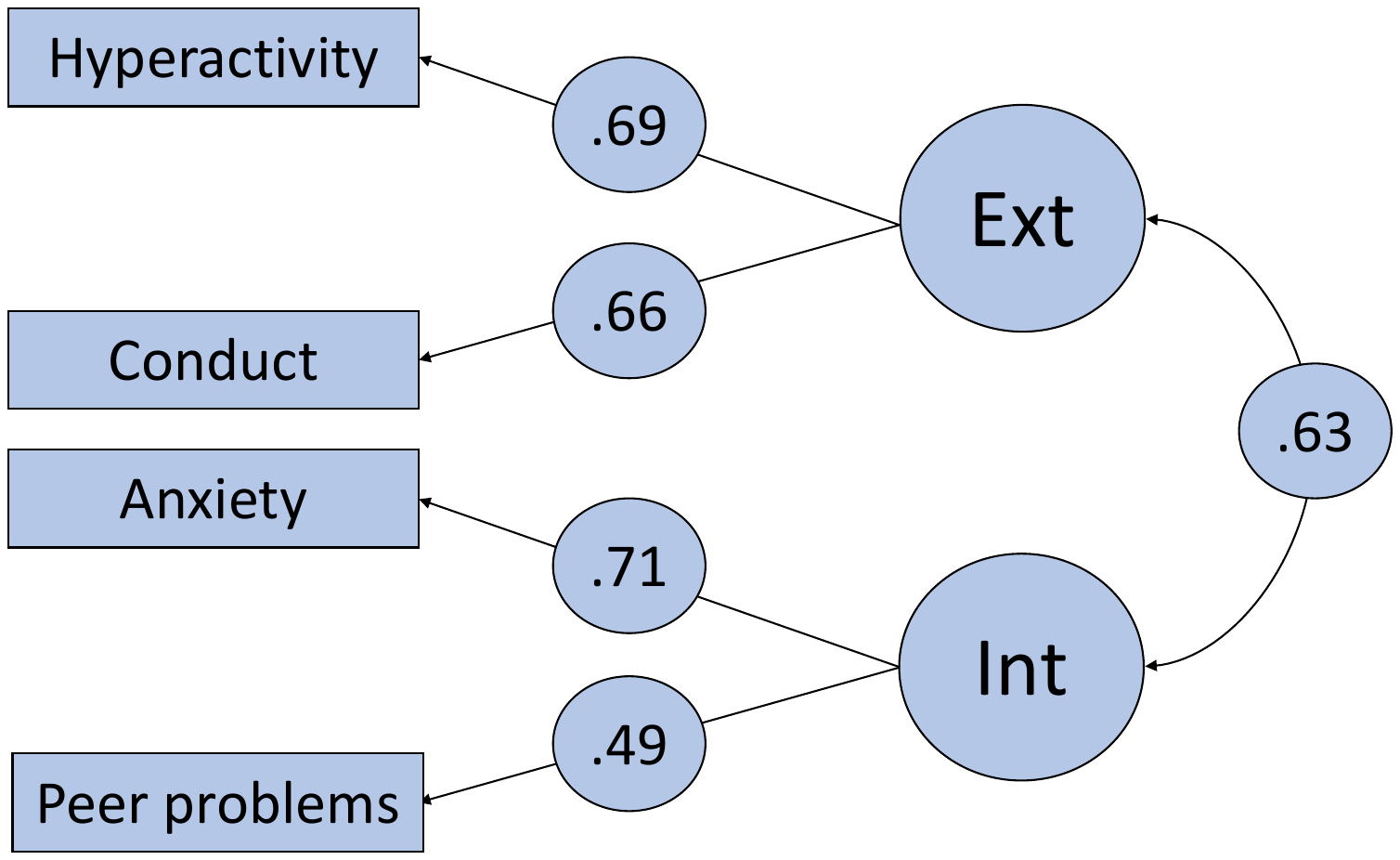 | 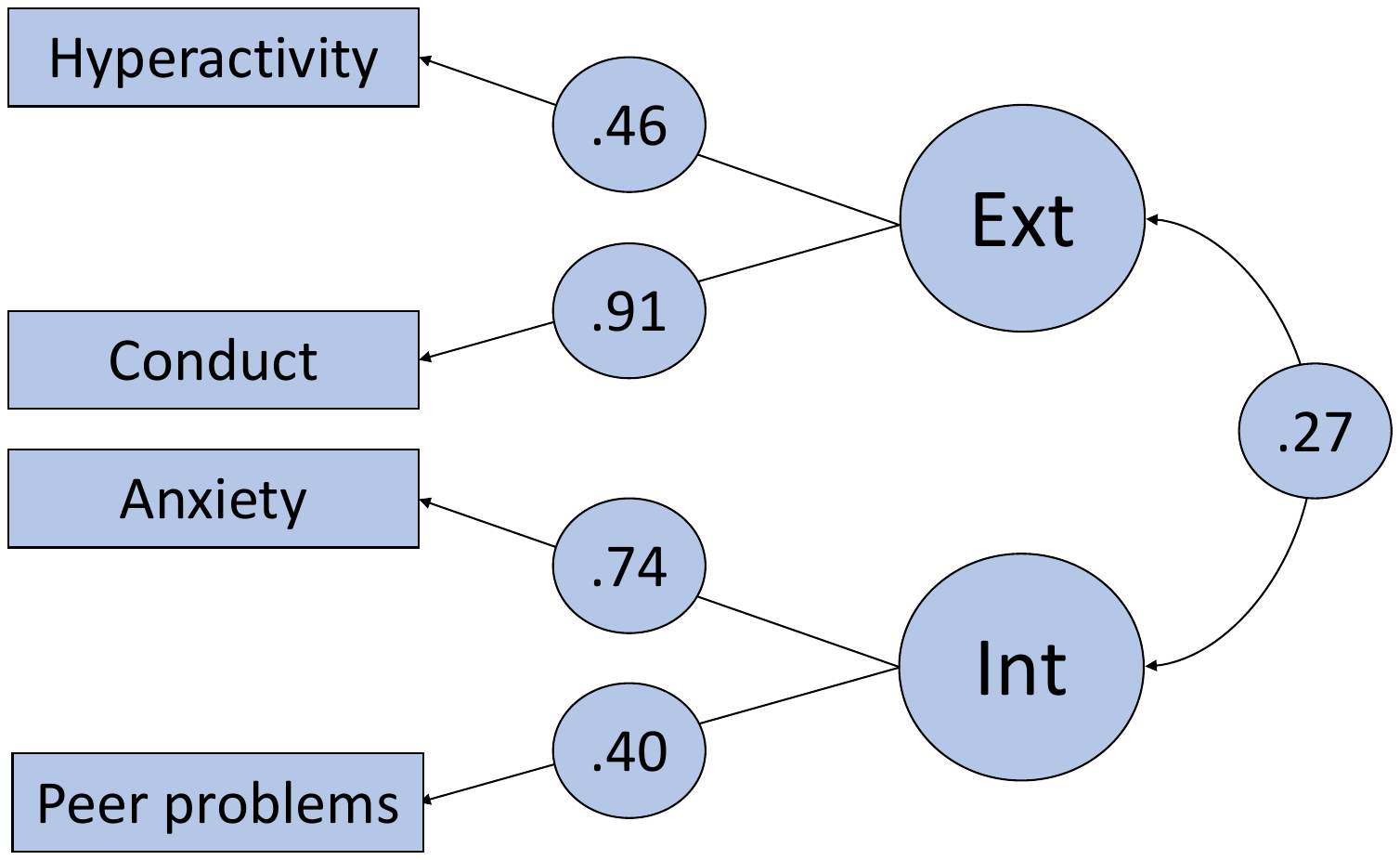 | 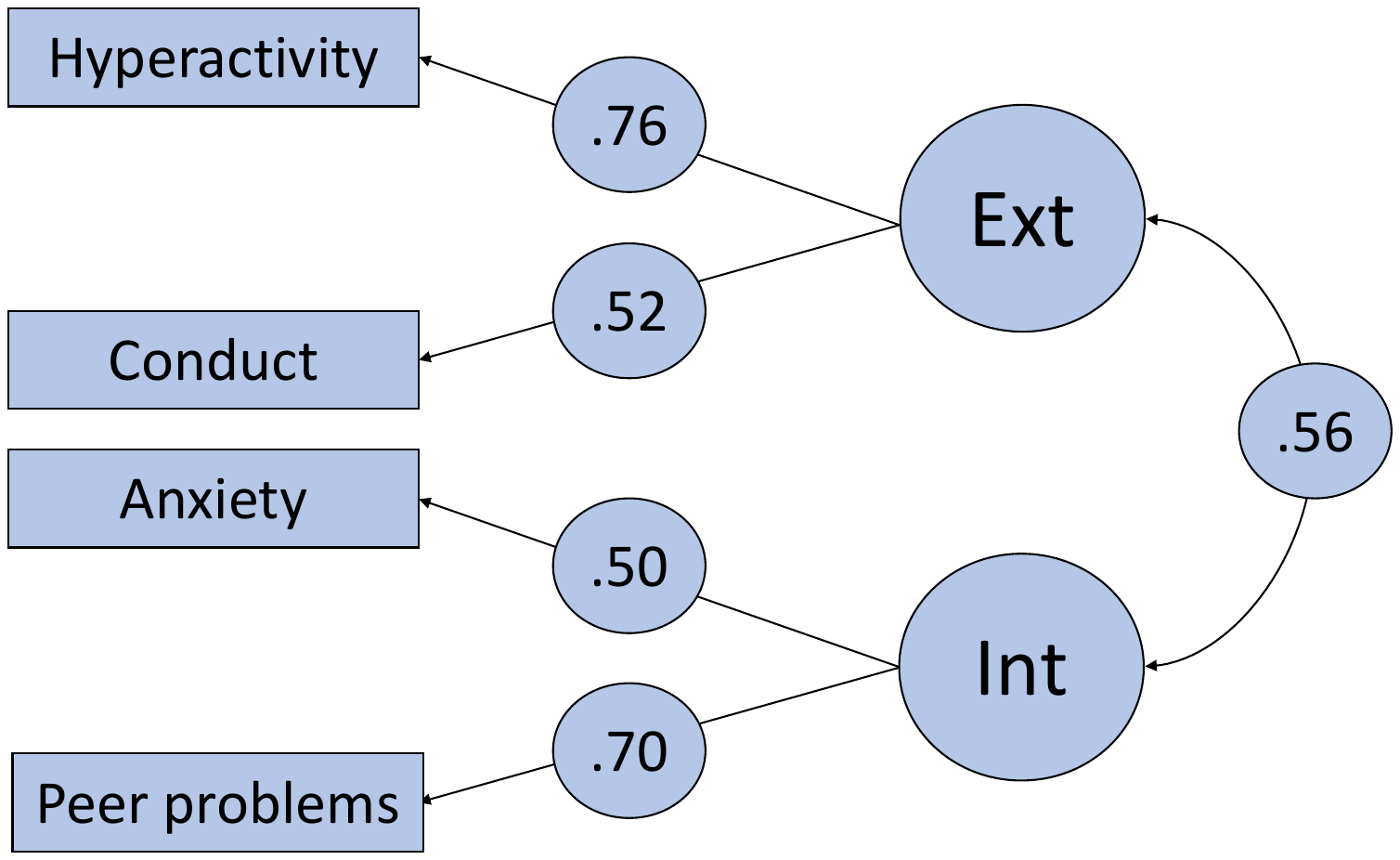 |
| *Note:* Ext= Externalizing behaviour problems factor; Int= Internalizing behaviour problems factor. | | |

***Supporting information 6.***

Confirmatory factor analysis (CFA).

To determine the best latent model for the data structure based on the two-factor structure yielded by the EFA, we created the BPp, externalizing and internalizing composites on the cross-age level, using the bifactor and hierarchical factor modelling.

Latent composites were created using the CFA data reduction technique that constructs the latent composites based on a pre-specified structure that underlies the data. CFA was conducted using *lavaan* for R (Rosseel, 2012; R Core Team, 2020). Full Information Maximum Likelihood was used to account for data missingness.

The hierarchical latent factor model includes two first-order factors (here, externalizing and internalizing) loading onto a second-order factor (here, general behaviour problems, which we refer to as BPp). In contrast, the bifactor model allows all constituent variables to index a general factor (here BPp) and specific factors that account for the residual variance (here, externalizing and internalizing). **Supporting figure S3** presents the structure of the hierarchical (**Supporting figure S3A**) and bifactor (**Supporting figure S3B**) models of BPp.

**Supporting figure** **S4** shows that externalizing and internalizing factors yielded moderate-to-high correlations across models and raters (0.81-0.92 in externalizing and 0.53-0.94 in internalizing). The bifactor and hierarchical BPp were highly correlated for parent ratings (0.74; **Supporting figure S4A**), but not teacher ratings or children’s self-ratings (0.20 and -0.16, respectively; **Supporting figures S4B and S4C**, respectively).

Finally, based on model fit indices, the hierarchical model provided significantly better fit than the bifactor model, hence the hierarchical solution was selected for downstream analyses.

**Supporting figure S3.** Hierarchical and bifactor cross-age model of BPp, externalizing and internalizing.

**A Hierarchical cross-age model**

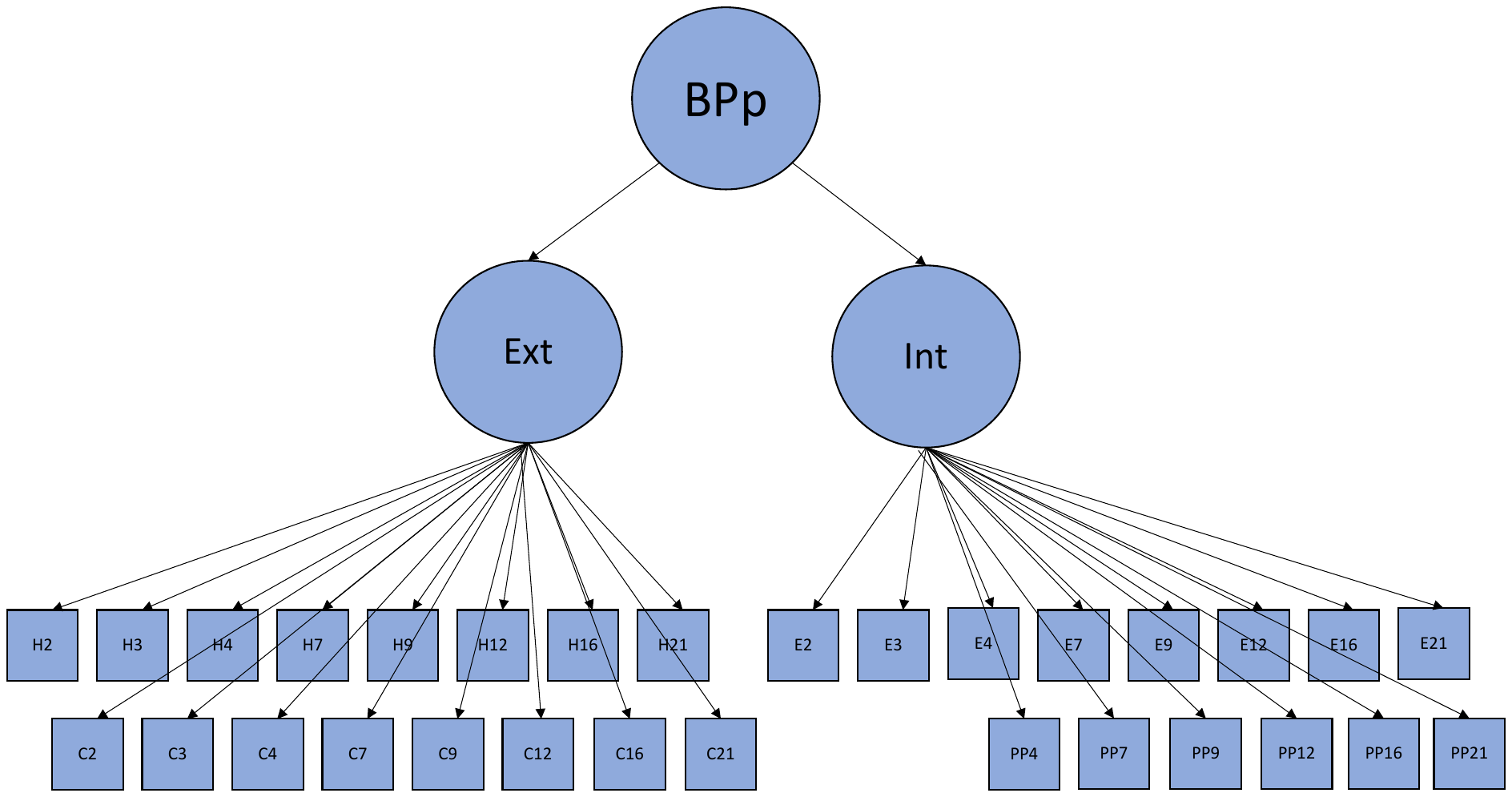

*Note.* We ran three separate hierarchical cross-age models: one for parent-report, one for teacher-report and one for self-report (child). H= hyperactivity; C= conduct; E= emotional problems; PP= peer problems; numbers indicate age of measurement.

**B Bifactor cross-age model**

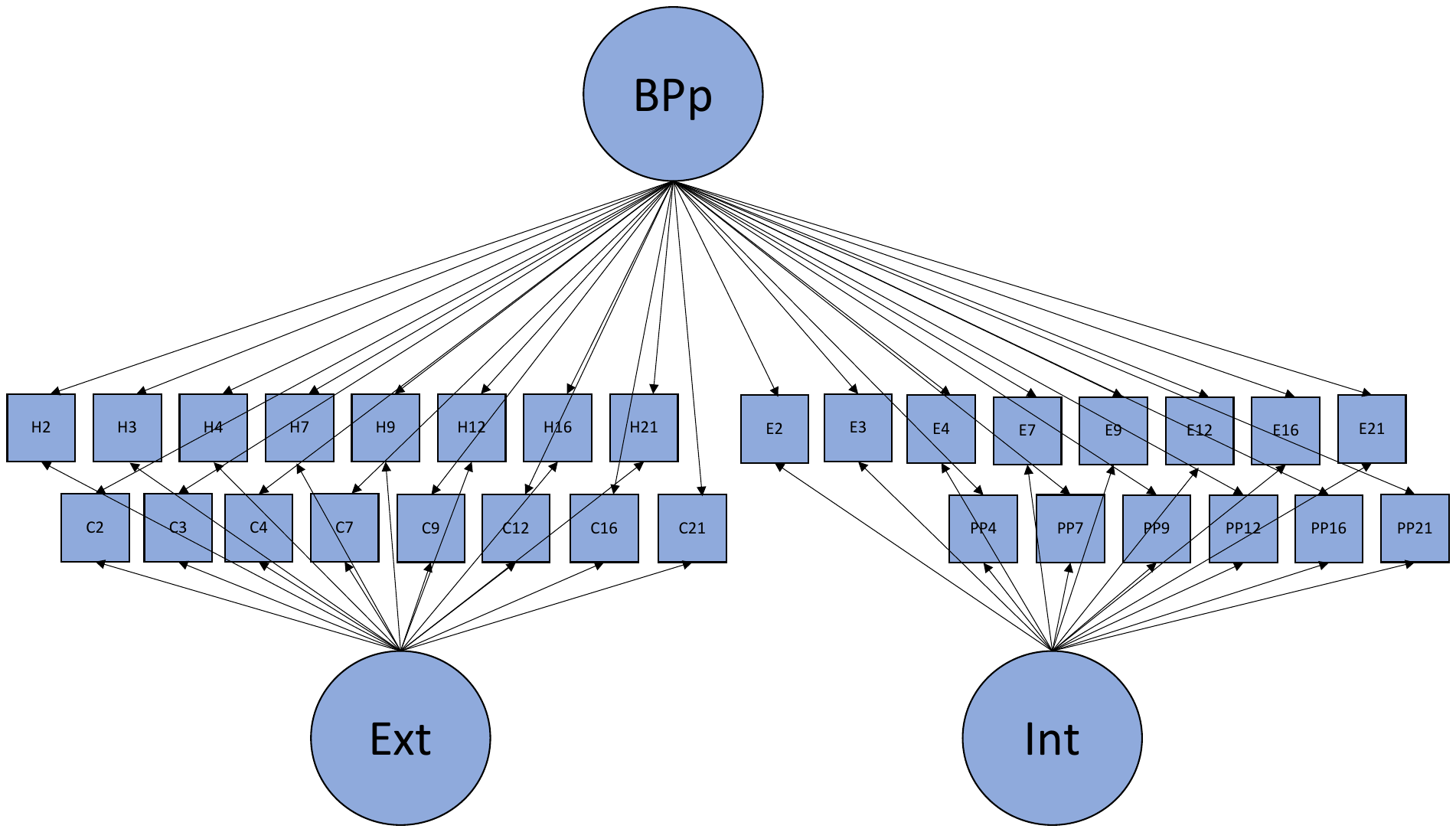

*Note.* We ran three separate bifactor cross-age models: one for parent-report, one for teacher-report and one for self-report (child). H= hyperactivity; C= conduct; E= emotional problems; PP= peer problems; numbers indicate age of measurement.

| **Supporting figure S4**. Correlations between hierarchical and bifactor composites of BPp, externalizing and internalizing for parent, teacher and child ratings. | | |
| --- | --- | --- |
| **A Parent ratings** | **B Teacher ratings** | **C Child ratings** |
| 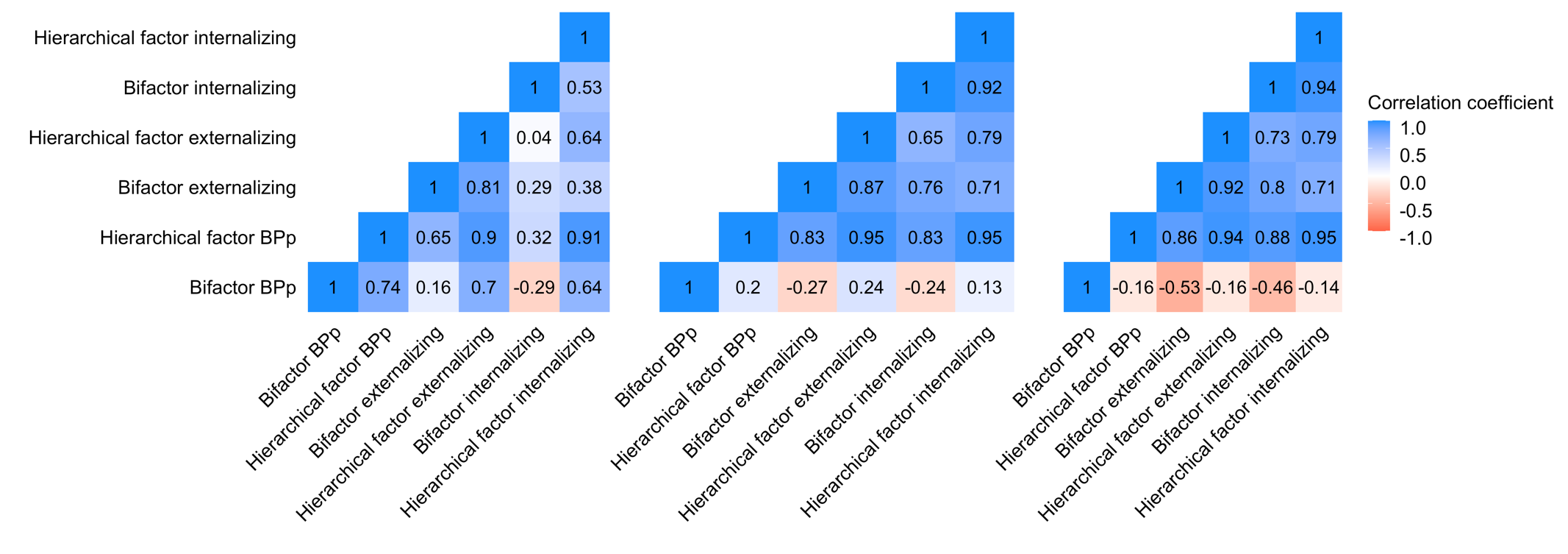 | | |

***Supporting information 7.***

Phenotypic and genetic correlations between cross-age and cross-rater composites.

Genetic correlations were estimated using the twin method, extended to the exploration of the covariance between pairs of traits, using OpenMx 2.0 for R (Neale et al., 2016; R Core Team, 2020). Bivariate genetic analysis allows for the decomposition of the covariance between multiple traits into genetic and environmental sources of variance, by modelling the cross-twin cross-trait covariances. Cross-twin cross-trait covariances describe the association between two variables, with twin 1’s score on variable 1 correlated with twin 2’s score on variable 2, which are calculated separately for monozygotic and dizygotic twins.

As presented in **Supporting figure S5**, cross-age and cross-rater effects were strongly correlated both phenotypically (range of r= 0.28- 0.93 for BPp, r= 0.26- 0.94 for externalizing and 0.27- 0.91 for internalizing) and genetically (range of r= 0.41- 0.94 for BPp, r= 0.40- 0.96 for externalizing and 0.39- 0.90 for internalizing). 95% confidence intervals for genetic correlations are presented in **Supporting table S10**.

**Supporting figure S5.** Phenotypic and genetic correlations between cross-age and cross-rater composites of BPp, externalizing and internalizing.

**Phenotypic correlations	Genetic correlations**

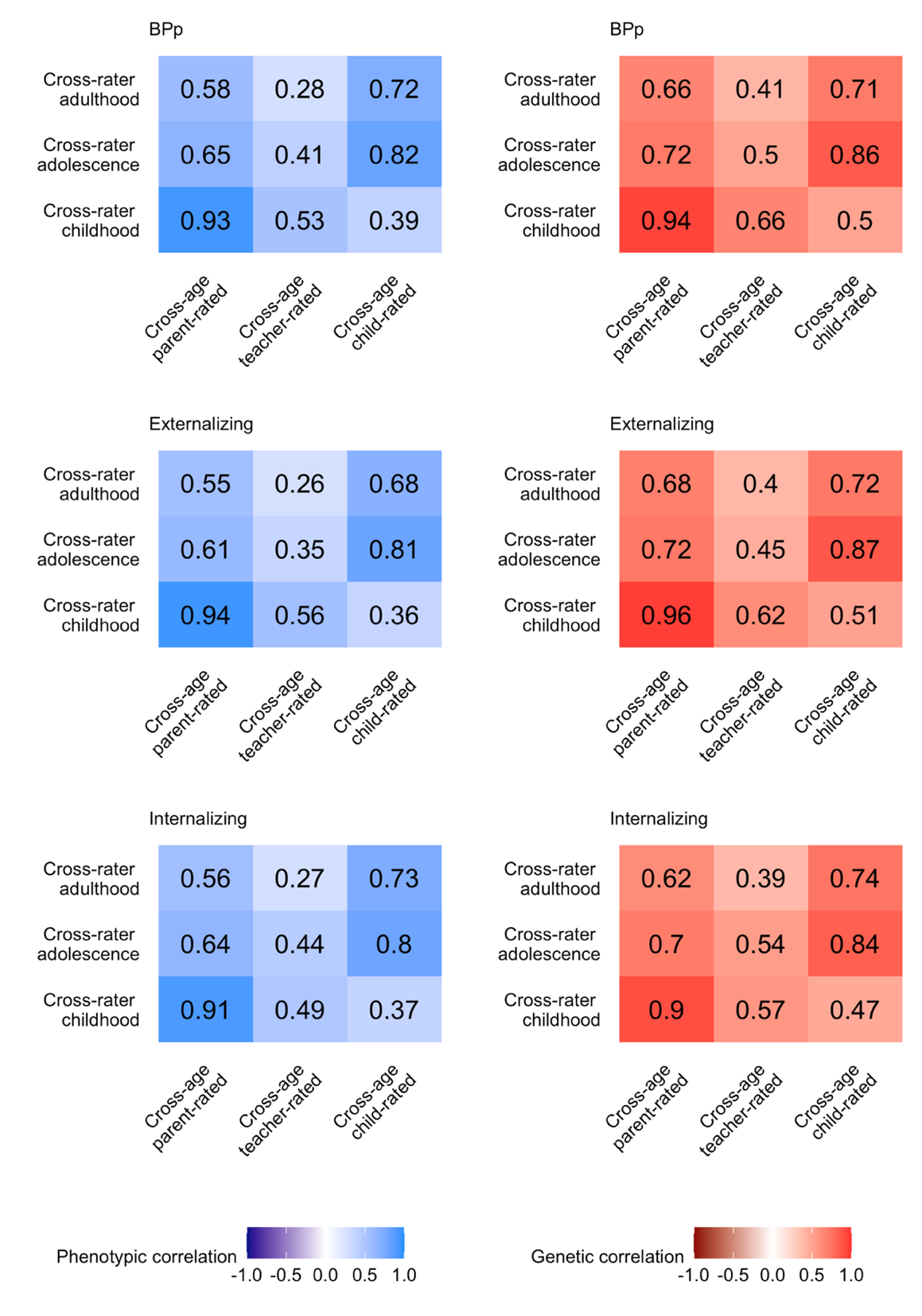

***Supporting information 8.***

Construction and results for the single-trait composites.

*Construction of the single-trait composites*

As a comparison to cross-age composites, we created separate measures of the four behaviour problems across ages but separately for parent, teacher and child ratings. As a comparison to cross-rater composites, we created separate measures of the four behaviour problems across raters but separately in childhood, adolescence and adulthood. We constructed the single-trait composites using one-factor confirmatory factor analysis, where the latent constructs of cross-age (**Supporting figure S6A**) and cross-rater (**Supporting figure S6B**) hyperactivity, conduct, emotional problems and peer problems were estimated in separate models.

**Supporting figure S6.** Summary of the construction of the single-trait cross-age and single-trait cross-rater composites.

**A** Single-trait cross-age composites.

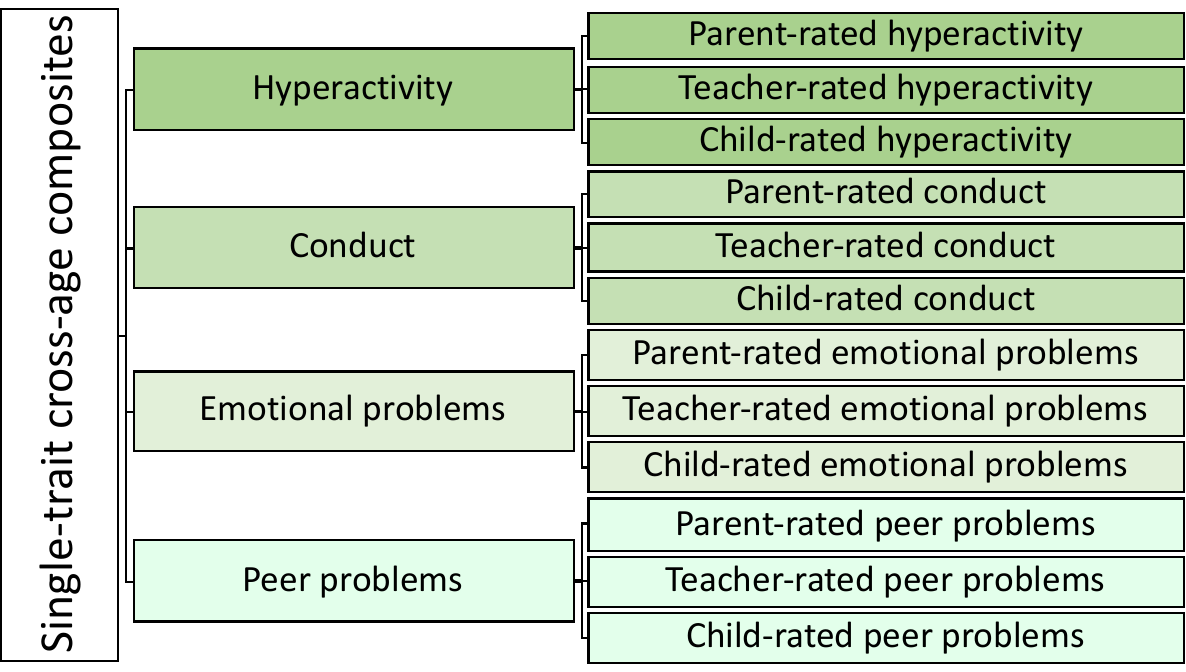

**B** Single-trait cross-rater composites.

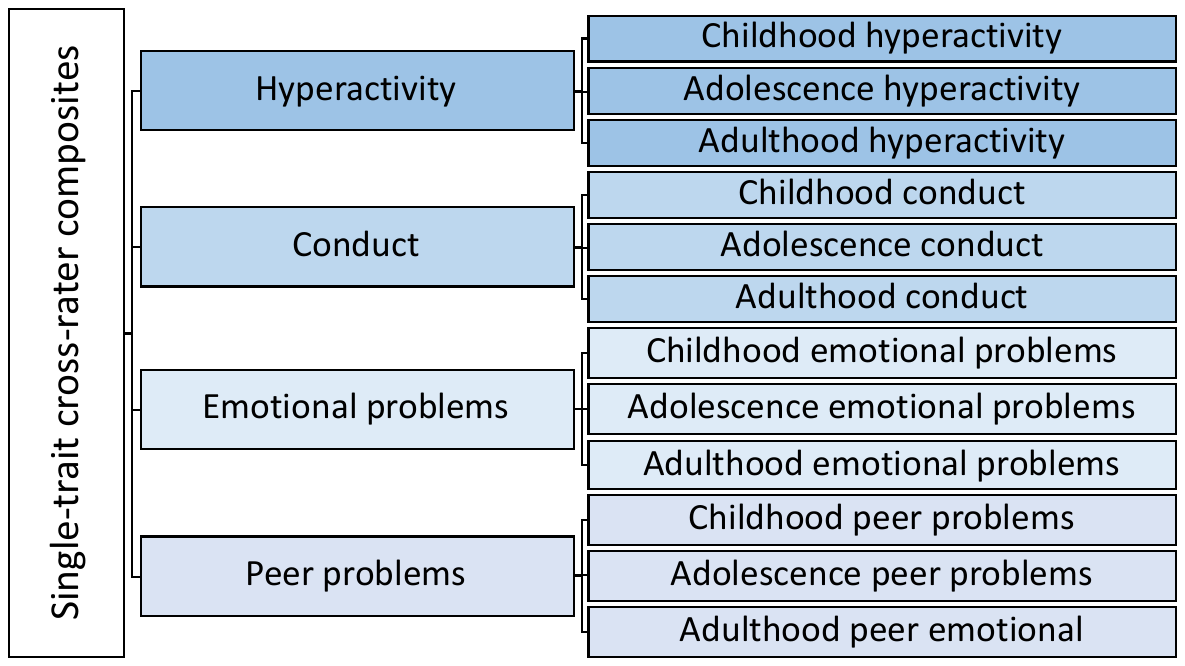

*Multi-GPS heritability: single-trait composites of behaviour problems*

**Supporting figure S7** focuses on the effects of cross-age and cross-rater compositing on the four individual SDQ scales rather than BPp, externalizing and internalizing. In general, we found similar but weaker results, suggesting the usefulness of compositing across traits as well as across ages and across raters.

For cross-age comparisons (**Supporting figure S7A**), we found that multi-GPS heritability increased for teacher-rated hyperactivity (3% vs 0.3%), parent and teacher-rated conduct (2.5% vs 0.9% and 2.5% vs 0.5%, respectively) and teacher and child-rated emotional problems (1.8% vs 0.1% and 2.2% vs 1.2%, respectively). While single-trait cross-age composites yielded a mean multi-GPS heritability of only 1.3%, compositing across ages and traits (BPp, externalizing, internalizing) yielded a mean multi-GPS heritability that was almost three times as high (3.6%). In addition, due to lack of the cross-age effect for parent-rated hyperactivity and peer problems, the highest cross-age effect observed for parent-rated BPp may be driven entirely by conduct and emotional problems.

Compositing across raters also generally increased multi-GPS heritability of single-trait composites (**Supporting figure 7B**). The multi-GPS heritability showed a notable drop from childhood to adulthood for conduct (4.8% in childhood to 3.7% in adolescence to 1.1% in adulthood), which was consistent with the pattern GPS results for cross-rater externalizing. An opposite trend was found for emotional problems, with the multi-GPS heritability increasing from childhood to adolescence and adulthood (1.3% in childhood to 3.3% in adolescence to 2% in adulthood), which also mirrored the direction of cross-rater internalizing. multi-GPS heritability for cross-rater composites of hyperactivity and peer problems remained relatively stable across development (the mean of 1.5% and 0.9%, respectively). Compared to the improvement of GPS prediction for the single-trait cross-rater composites (mean of 1.6% vs 0.9% for individual traits), the cross-rater effect was more than quadrupled for the cross-rater composites of BPp, externalizing and internalizing (mean of 3.6% vs 0.8% for individual traits).

*Twin heritability: single-trait composites of behaviour problems*

The twin results for the single-trait composites showed both cross-age (**Supporting figure S7C**) and cross-rater (**Supporting figure S7D**) effects, mirroring the twin results for BPp, externalizing and internalizing. However, the magnitudes of these effects were lower for single-trait composites, with the mean of 57% for cross-age composites vs 51% for observed traits and 61% for cross-rater composites vs 51% for observed traits.

**Supporting figure S7.** Multi-GPS and twin heritability results for single-trait cross-age composites and single-trait cross-rater composites as compared to the mean multi-GPS and twin heritability of observed traits.

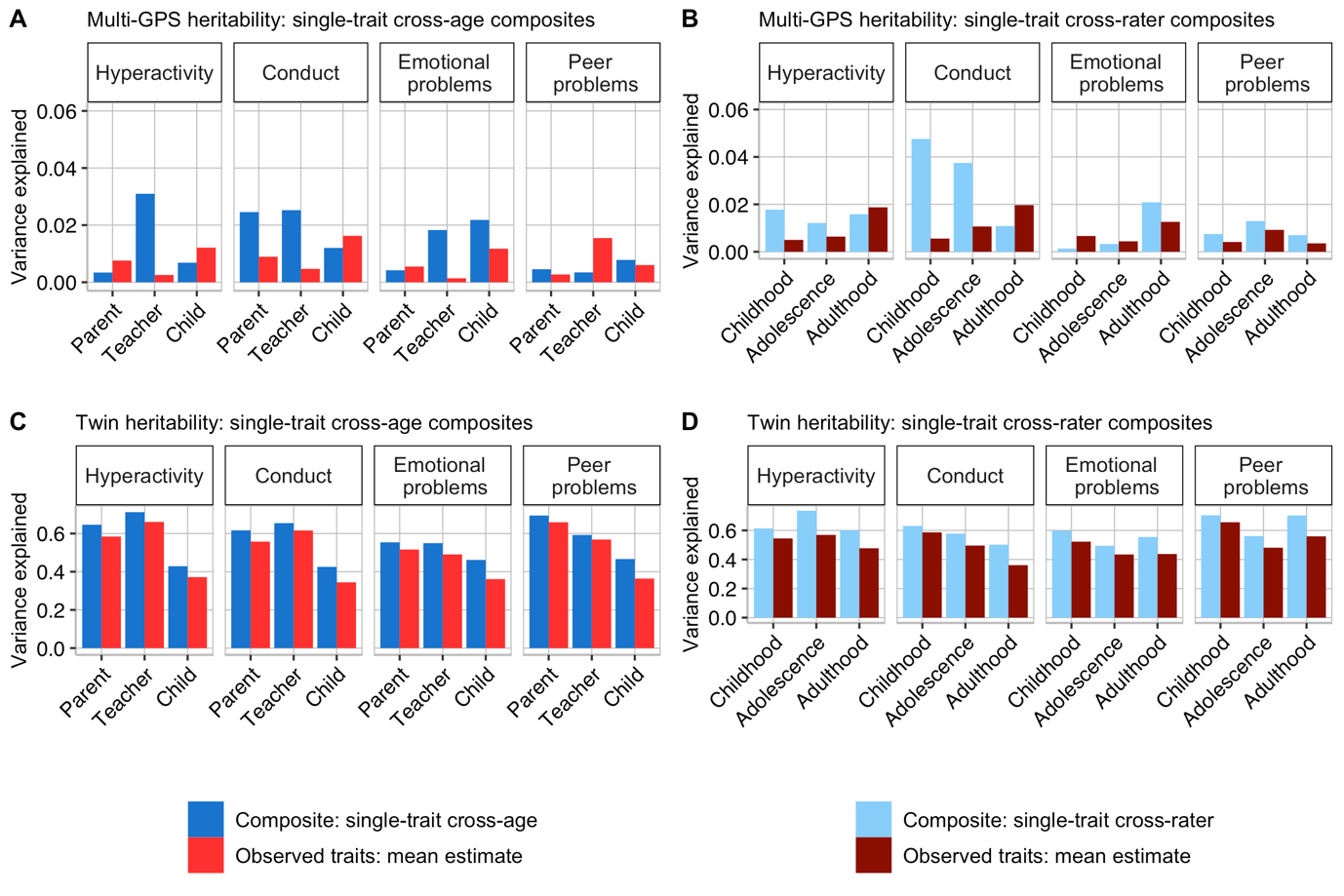

***Supporting information 9.***

Elastic net regularization.

We estimated the joint prediction of the 15 GPS in a penalized regression elastic net model with out-of-sample tests of prediction accuracy, using the R package glmnet (Hastie & Qian, 2014) implemented in caret (Kuhn, 2012; R Core Team, 2020). We used a shrinkage model referred to as elastic net regularization to overcome problems of multicollinearity and overfitting (Zou et al., 2005). Elastic net produces regression models that are penalised with both the L1-norm (Lasso) and L2-norm (Ridge) penalties in order to omit highly correlated predictors and reduce the risk of multicollinearity and overfitting (Pavlou et al., 2016).

Elastic net regularization tries to minimise the following loss function:

||y − X𝛽||2 + λ(α*|β|1 + (1−α)*|β|2)

where ||y–X’β||2 is the residual sum of squares, |β|2 is the sum of the squared betas (the L2 penalty), |β|1 is the sum of the absolute betas (the L1 penalty) and X is an N*P (‘N’ observations and ‘P’ predictors) matrix of polygenic scores (for details, see Allegrini et al., 2020).

For every model tested, we randomly split the sample into an independent training set (80%) and a hold-out set (20%). In the training set, we performed the 10-fold cross-validation repeated 100 times to select the model that minimises the Root Mean Square Error (RMSE), which indicates the smallest cross-validation error (Fushiki, 2011). To account for patterns of missingness in the phenotypic data and to use all available data we employed the Full Information Maximum Likelihood (FIML) method. We then estimated variance explained (R2) in the hold-out test set.

***Supporting information 10.***

Meta-analytic approach to comparing multi-GPS heritability between composites and observed traits.

In the meta-analytic approach, we used a random effects meta-regression model to estimate the grand mean of multi-GPS and twin heritability of the observed traits. For multi-GPS analyses, we used the square root of R^2^ (that is, the multiple correlation between predictors and a composite in the hold-out set) and the 95% confidence intervals of R as a measure of effect size. The square root of R^2^ was computed in order to estimate 95% confidence intervals, which could not be estimated from the R^2^_._

For twin analyses, the effect size was measured as twin heritability with 95% confidence intervals. Meta-analysis was conducted in the R package *metafor* (Viechtbauer & Viechtbauer, 2015; R Core Team, 2020).

**Supporting figure 8** shows the multi-GPS heritability results for cross-age composites (**Supporting figure S8A**) and cross-rater composites (**Supporting figure S8B**), as compared to grand mean of observed traits. Twin heritability results are presented in **Supporting figure S8C** for cross-age composites and **Supporting figure S8D** for cross-rater composites, as compared to grand mean of observed traits.

**Supporting figure 9** shows the multi-GPS heritability results for single-trait cross-age composites (**Supporting figure S9A**) and single-trait cross-rater composites (**Supporting figure S9B**), as compared to grand mean of observed traits. Twin heritability results are presented in **Supporting figure S9C** for single-trait cross-age composites and **Supporting figure S9D** for single-trait cross-rater composites, as compared to grand mean of observed traits.

**Supporting figure S8.** Multi-GPS correlation and twin heritability results for cross-age composites and cross-rater composites as compared to the grand mean multi-GPS correlation and twin heritability of observed traits.

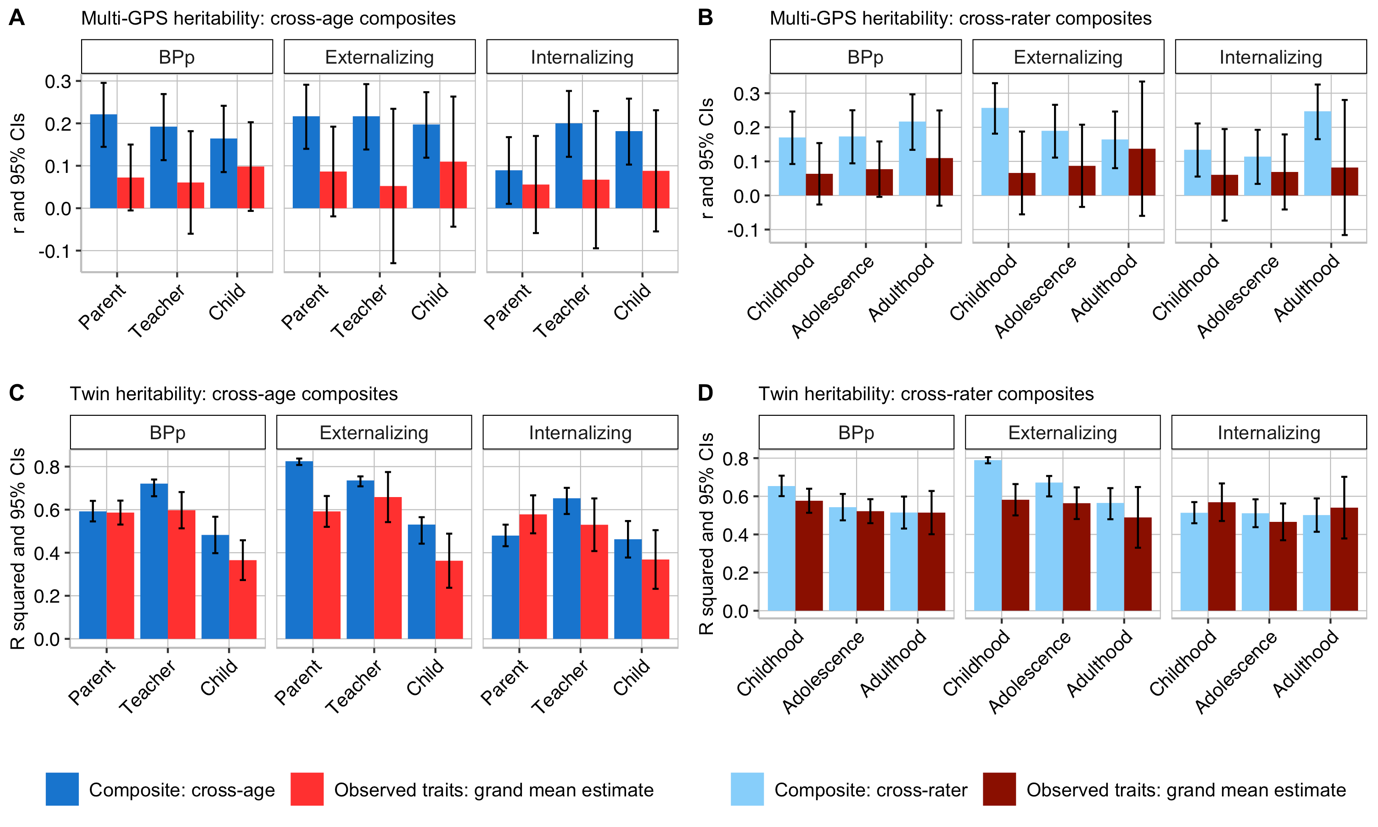

*Note.* R squared= variance explained; r= correlation coefficient; 95% CIs= confidence intervals.

**Supporting figure S9.** Multi-GPS correlation and twin heritability results for single-trait cross-age composites and single-trait cross-rater composites as compared to the grand mean multi-GPS correlation and twin heritability of observed traits.

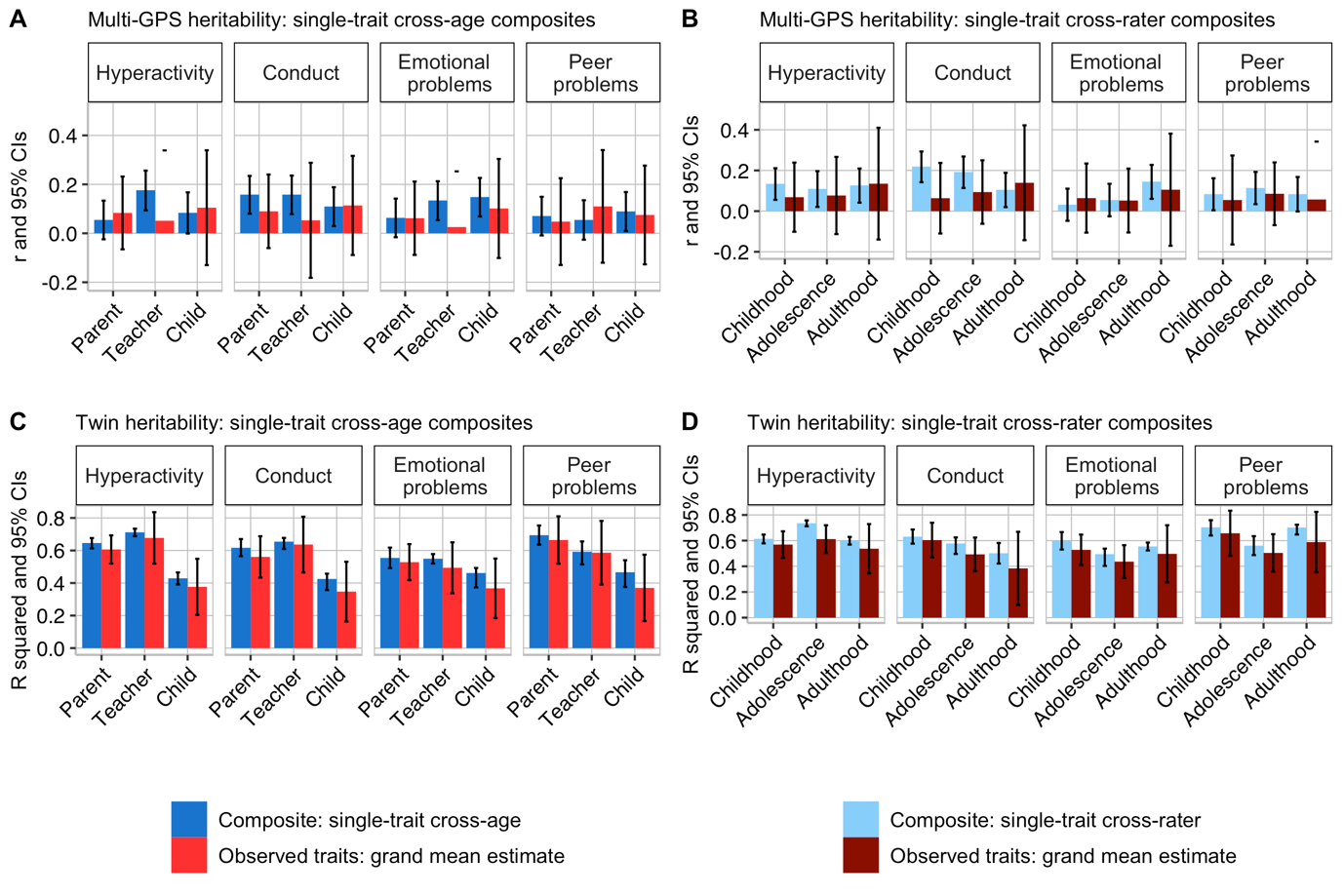

*Note.* R squared= variance explained; r= correlation coefficient; 95% CIs= confidence intervals.

***Supporting tables***

| **Supporting table S1**. Behaviour problems composites: sample characteristics. | | | | |
| --- | --- | --- | --- | --- |
| Note. | | | | |
| N= sample size; MZ= monozygotic; DZ= dizygotic. | | | | |
| * cross-age approach: combined individual behaviour problems at all ages (2-21) for parent, teacher and child ratings to create the first order factors of cross-age externalizing and internalizing, which were then combined across raters to create the second-order factors of cross-age-and-rater externalizing and internalizing. | | | | |
| ** cross-rater approach: combined individual behaviour problems in adolescence and adulthood to create the first order factors of cross-rater externalizing and internalizing, which were then combined across developmental stages to create the second-order factors of cross-age-and-rater externalizing and internalizing. | | | | |
| **Composites** | **N twin pairs MZ** | **N twin pairs DZ** | **N total twin pairs** | **N genotyped** |
| **Cross-age** |  |  |  |  |
| BPp parent-rated | 1753 | 3025 | 4778 | 3065 |
| BPp teacher-rated | 1675 | 2892 | 4567 | 2953 |
| BPp child-rated | 1753 | 3025 | 4778 | 2986 |
| Externalizing parent-rated | 1753 | 3025 | 4778 | 3065 |
| Externalizing teacher-rated | 1675 | 2892 | 4567 | 2953 |
| Externalizing child-rated | 1753 | 3025 | 4778 | 2986 |
| Internalizing parent-rated | 1753 | 3025 | 4778 | 3065 |
| Internalizing teacher-rated | 1675 | 2892 | 4567 | 2953 |
| Internalizing child-rated | 1753 | 3025 | 4778 | 2986 |
| **Cross-rater** |  |  |  |  |
| BPp childhood | 1753 | 3025 | 4778 | 3065 |
| BPp adolescence | 1711 | 2920 | 4631 | 2986 |
| BPp adulthood | 1472 | 2474 | 3946 | 2648 |
| Externalizing childhood | 1753 | 3025 | 4778 | 3065 |
| Externalizing adolescence | 1711 | 2920 | 4631 | 2986 |
| Externalizing adulthood | 1472 | 2474 | 3946 | 2648 |
| Internalizing childhood | 1753 | 3025 | 4778 | 3065 |
| Internalizing adolescence | 1711 | 2920 | 4631 | 2986 |
| Internalizing adulthood | 1472 | 2474 | 3946 | 2648 |
| **Cross-age-and-rater (cross-age approach) *** |  |  |  |  |
| BPp | 1753 | 3025 | 4778 | 3065 |
| Externalizing | 1753 | 3025 | 4778 | 3065 |
| Internalizing | 1753 | 3025 | 4778 | 3065 |
| **Cross-age-and-rater (cross-rater approach) **** |  |  |  |  |
| BPp | 1753 | 3025 | 4778 | 3065 |
| Externalizing | 1753 | 3025 | 4778 | 3065 |
| Internalizing | 1753 | 3025 | 4778 | 3065 |
| **Single-trait cross-age** |  |  |  |  |
| Hyperactivity parent-rated | 1753 | 3025 | 4778 | 3065 |
| Hyperactivity teacher-rated | 1547 | 2665 | 4212 | 2744 |
| Hyperactivity child-rated | 1753 | 3025 | 4778 | 2691 |
| Conduct parent-rated | 1753 | 3025 | 4778 | 3065 |
| Conduct teacher-rated | 1675 | 2892 | 4567 | 2953 |
| Conduct child-rated | 1753 | 3025 | 4778 | 2986 |
| Emotional problems parent-rated | 1753 | 3025 | 4778 | 3065 |
| Emotional problems teacher-rated | 1674 | 2891 | 4565 | 2952 |
| Emotional problems child-rated | 1753 | 3025 | 4778 | 2986 |
| Peer problems parent-rated | 1753 | 3025 | 4778 | 3065 |
| Peer problems teacher-rated | 1674 | 2892 | 4566 | 2953 |
| Peer problems child-rated | 1753 | 3025 | 4778 | 2986 |
| **Single-trait cross-rater** |  |  |  |  |
| Hyperactivity childhood | 1753 | 3025 | 4778 | 3065 |
| Hyperactivity adolescence | 1370 | 2342 | 3712 | 2432 |
| Hyperactivity adulthood | 1753 | 3025 | 4778 | 2648 |
| Conduct childhood | 1753 | 3025 | 4778 | 3065 |
| Conduct adolescence | 1710 | 2916 | 4626 | 2983 |
| Conduct adulthood | 1753 | 3025 | 4778 | 2648 |
| Emotional problems childhood | 1753 | 3025 | 4778 | 3065 |
| Emotional problems adolescence | 1709 | 2915 | 4624 | 2984 |
| Emotional problems adulthood | 1753 | 3025 | 4778 | 2647 |
| Peer problems childhood | 1750 | 3022 | 4772 | 3060 |
| Peer problems adolescence | 1709 | 2915 | 4624 | 2984 |
| Peer problems adulthood | 1750 | 3022 | 4772 | 2648 |

| **Supporting table S2**. Behaviour problems composites: model fit indices and predictions from elastic net regularization. | | | | | | | | | | | |
| --- | --- | --- | --- | --- | --- | --- | --- | --- | --- | --- | --- |
| Note. | | | | | | | | | | | |
| FRCT= fraction of SNPs; CV= cross-validated; SD= standard deviation; R2= variance explained; RMSE= root mean square error; train= training set (80%); test= hold-out set (20%). | | | | | | | | | | | |
| * cross-age approach: combined individual behaviour problems at all ages (2-21) for parent, teacher and child ratings to create the first order factors of cross-age externalizing and internalizing, which were then combined across raters to create the second-order factors of cross-age-and-rater externalizing and internalizing. | | | | | | | | | | | |
| ** cross-rater approach: combined individual behaviour problems in adolescence and adulthood to create the first order factors of cross-rater externalizing and internalizing, which were then combined across developmental stages to create the second-order factors of cross-age-and-rater externalizing and internalizing. | | | | | | | | | | | |
| **Composites** | **Elastic net regularization** | | | | | | | | | | |
|  | **FRCT** | **Mean CV RMSE (train)** | **SD CV**  **RMSE (train)** | **Mean CV R2 (train)** | **SD CV**  **R2 (train)** | **RMSE** | **R2** | **Alpha** | **Lambda** | **N (train)** | **N (test)** |
| **Cross-age** |  |  |  |  |  |  |  |  |  |  |  |
| BPp parent-rated | 1 | 0.973 | 0.001 | 0.023 | 0.001 | 0.981 | 0.049 | 0.6 | 0.04 | 2453 | 612 |
| BPp teacher-rated | 1 | 0.977 | 0.002 | 0.028 | 0.002 | 0.941 | 0.037 | 1.0 | 0.02 | 2365 | 588 |
| BPp child-rated | 1 | 0.981 | 0.002 | 0.030 | 0.003 | 0.979 | 0.027 | 0.2 | 0.04 | 2390 | 596 |
| Externalizing  parent-rated | 1 | 0.972 | 0.002 | 0.034 | 0.002 | 0.948 | 0.047 | 0.9 | 0.02 | 2453 | 612 |
| Externalizing  teacher-rated | 1 | 0.957 | 0.002 | 0.035 | 0.002 | 0.990 | 0.047 | 0.8 | 0.02 | 2365 | 588 |
| Externalizing  child-rated | 1 | 0.978 | 0.002 | 0.030 | 0.002 | 0.985 | 0.039 | 0.1 | 0.09 | 2390 | 596 |
| Internalizing  parent-rated | 1 | 0.984 | 0.001 | 0.019 | 0.001 | 0.976 | 0.008 | 0.8 | 0.02 | 2453 | 612 |
| Internalizing  teacher-rated | 1 | 0.978 | 0.001 | 0.014 | 0.001 | 0.996 | 0.040 | 0.5 | 0.03 | 2365 | 588 |
| Internalizing  child-rated | 1 | 0.977 | 0.002 | 0.024 | 0.002 | 1.007 | 0.033 | 0.1 | 0.05 | 2390 | 596 |
| **Cross-rater** |  |  |  |  |  |  |  |  |  |  |  |
| BPp childhood | 1 | 0.964 | 0.002 | 0.030 | 0.002 | 0.989 | 0.029 | 0.1 | 0.11 | 2453 | 612 |
| BPp adolescence | 1 | 0.983 | 0.002 | 0.029 | 0.003 | 0.958 | 0.030 | 1.0 | 0.02 | 2390 | 596 |
| BPp adulthood | 1 | 0.982 | 0.002 | 0.029 | 0.002 | 0.959 | 0.047 | 0.1 | 0.10 | 2120 | 528 |
| Externalizing  childhood | 1 | 0.965 | 0.002 | 0.034 | 0.002 | 0.941 | 0.066 | 0.1 | 0.13 | 2453 | 612 |
| Externalizing adolescence | 1 | 0.988 | 0.003 | 0.038 | 0.003 | 0.948 | 0.036 | 0.9 | 0.03 | 2390 | 596 |
| Externalizing  adulthood | 1 | 0.980 | 0.002 | 0.035 | 0.003 | 0.978 | 0.027 | 0.1 | 0.11 | 2120 | 528 |
| Internalizing  childhood | 1 | 0.987 | 0.001 | 0.016 | 0.001 | 0.960 | 0.018 | 0.7 | 0.03 | 2453 | 612 |
| Internalizing adolescence | 1 | 0.984 | 0.001 | 0.022 | 0.001 | 1.004 | 0.013 | 0.2 | 0.05 | 2390 | 596 |
| Internalizing  adulthood | 1 | 0.978 | 0.001 | 0.021 | 0.002 | 0.968 | 0.061 | 0.7 | 0.04 | 2120 | 528 |
| **Cross-age-and-rater (cross-age approach) *** |  |  |  |  |  |  |  |  |  |  |  |
| BPp | 1 | 0.974 | 0.002 | 0.028 | 0.002 | 0.957 | 0.040 | 0.1 | 0.05 | 2453 | 612 |
| Externalizing | 1 | 0.974 | 0.002 | 0.035 | 0.002 | 0.926 | 0.035 | 0.7 | 0.03 | 2453 | 612 |
| Internalizing | 1 | 0.982 | 0.001 | 0.022 | 0.001 | 0.979 | 0.027 | 0.5 | 0.04 | 2453 | 612 |
| **Cross-age-and-rater (cross-rater**  **approach) **** |  |  |  |  |  |  |  |  |  |  |  |
| BPp | 1 | 0.974 | 0.002 | 0.032 | 0.001 | 0.971 | 0.027 | 0.1 | 0.05 | 2453 | 612 |
| Externalizing | 1 | 0.968 | 0.002 | 0.033 | 0.002 | 0.986 | 0.040 | 0.8 | 0.02 | 2453 | 612 |
| Internalizing | 1 | 0.976 | 0.001 | 0.024 | 0.002 | 0.995 | 0.028 | 0.2 | 0.04 | 2453 | 612 |
| **Single-trait cross-age** |  |  |  |  |  |  |  |  |  |  |  |
| Hyperactivity  parent-rated | 1 | 0.985 | 0.001 | 0.019 | 0.002 | 0.968 | 0.003 | 0.6 | 0.05 | 2453 | 612 |
| Hyperactivity  teacher-rated | 1 | 0.994 | 0.001 | 0.029 | 0.000 | 0.980 | 0.031 | 0.6 | 0.03 | 2196 | 548 |
| Hyperactivity  child-rated | 1 | 0.992 | 0.001 | 0.016 | 0.001 | 0.986 | 0.007 | 0.8 | 0.04 | 2155 | 536 |
| Conduct  parent-rated | 1 | 0.985 | 0.001 | 0.023 | 0.000 | 0.971 | 0.025 | 0.1 | 0.05 | 2453 | 612 |
| Conduct  teacher-rated | 1 | 0.958 | 0.001 | 0.021 | 0.001 | 0.910 | 0.025 | 0.6 | 0.04 | 2364 | 589 |
| Conduct  child-rated | 1 | 0.986 | 0.002 | 0.023 | 0.002 | 1.029 | 0.012 | 1.0 | 0.02 | 2390 | 596 |
| Emotional problems parent-rated | 1 | 0.988 | 0.001 | 0.013 | 0.000 | 0.977 | 0.004 | 0.4 | 0.04 | 2453 | 612 |
| Emotional problems teacher-rated | 1 | 0.996 | 0.000 | 0.008 | 0.000 | 0.965 | 0.018 | 0.2 | 0.06 | 2364 | 588 |
| Emotional problems child-rated | 1 | 0.988 | 0.001 | 0.016 | 0.000 | 1.010 | 0.022 | 0.1 | 0.09 | 2390 | 596 |
| Peer problems  parent-rated | 1 | 1.008 | 0.001 | 0.007 | 0.000 | 0.991 | 0.005 | 0.4 | 0.06 | 2453 | 612 |
| Peer problems  teacher-rated | 1 | 1.008 | 0.001 | 0.003 | 0.000 | 1.012 | 0.003 | 0.6 | 0.05 | 2365 | 588 |
| Peer problems  child-rated | 1 | 1.002 | 0.001 | 0.008 | 0.001 | 1.018 | 0.008 | 0.3 | 0.06 | 2390 | 596 |
| **Single-trait cross-rater** |  |  |  |  |  |  |  |  |  |  |  |
| Hyperactivity  childhood | 1 | 0.980 | 0.001 | 0.022 | 0.001 | 0.967 | 0.018 | 0.2 | 0.05 | 2453 | 612 |
| Hyperactivity adolescence | 1 | 0.990 | 0.001 | 0.014 | 0.001 | 1.012 | 0.012 | 0.7 | 0.03 | 1948 | 484 |
| Hyperactivity  adulthood | 1 | 1.004 | 0.001 | 0.013 | 0.001 | 0.985 | 0.016 | 0.2 | 0.09 | 2120 | 528 |
| Conduct childhood | 1 | 0.980 | 0.001 | 0.017 | 0.001 | 0.974 | 0.048 | 0.6 | 0.04 | 2453 | 612 |
| Conduct adolescence | 1 | 0.990 | 0.001 | 0.021 | 0.001 | 1.006 | 0.037 | 0.1 | 0.09 | 2387 | 596 |
| Conduct adulthood | 1 | 0.984 | 0.001 | 0.015 | 0.001 | 1.000 | 0.011 | 0.5 | 0.04 | 2120 | 528 |
| Emotional problems childhood | 1 | 0.998 | 0.001 | 0.010 | 0.001 | 0.934 | 0.001 | 0.8 | 0.04 | 2453 | 612 |
| Emotional problems adolescence | 1 | 0.987 | 0.001 | 0.020 | 0.001 | 1.042 | 0.003 | 0.6 | 0.02 | 2388 | 596 |
| Emotional problems adulthood | 1 | 0.987 | 0.001 | 0.022 | 0.002 | 0.979 | 0.021 | 0.7 | 0.05 | 2119 | 528 |
| Peer problems childhood | 1 | 1.010 | 0.001 | 0.005 | 0.000 | 0.992 | 0.007 | 0.5 | 0.05 | 2448 | 612 |
| Peer problems adolescence | 1 | 1.018 | 0.000 | 0.006 | 0.000 | 0.980 | 0.013 | 0.4 | 0.05 | 2388 | 596 |
| Peer problems adulthood | 1 | 0.985 | 0.001 | 0.010 | 0.001 | 0.989 | 0.007 | 0.3 | 0.07 | 2120 | 528 |
| **Cross-age** |  |  |  |  |  |  |  |  |  |  |  |
| BPp parent-rated | 0.3 | 0.977 | 0.001 | 0.015 | 0.000 | 0.989 | 0.031 | 0.7 | 0.04 | 2453 | 612 |
| BPp teacher-rated | 0.3 | 0.979 | 0.001 | 0.023 | 0.001 | 0.944 | 0.032 | 0.2 | 0.05 | 2365 | 588 |
| BPp child-rated | 0.3 | 0.984 | 0.002 | 0.025 | 0.003 | 0.983 | 0.018 | 0.1 | 0.04 | 2390 | 596 |
| Externalizing  parent-rated | 0.3 | 0.979 | 0.001 | 0.019 | 0.001 | 0.952 | 0.046 | 1.0 | 0.02 | 2453 | 612 |
| Externalizing  teacher-rated | 0.3 | 0.959 | 0.002 | 0.030 | 0.001 | 0.995 | 0.033 | 0.8 | 0.01 | 2365 | 588 |
| Externalizing  child-rated | 0.3 | 0.980 | 0.002 | 0.027 | 0.002 | 0.994 | 0.022 | 0.2 | 0.04 | 2390 | 596 |
| Internalizing  parent-rated | 0.3 | 0.986 | 0.001 | 0.014 | 0.001 | 0.978 | 0.004 | 0.4 | 0.04 | 2453 | 612 |
| Internalizing  teacher-rated | 0.3 | 0.979 | 0.001 | 0.012 | 0.001 | 1.002 | 0.028 | 0.8 | 0.04 | 2365 | 588 |
| Internalizing  child-rated | 0.3 | 0.978 | 0.001 | 0.021 | 0.002 | 1.015 | 0.018 | 0.1 | 0.04 | 2390 | 596 |
| **Cross-rater** |  |  |  |  |  |  |  |  |  |  |  |
| BPp childhood | 0.3 | 0.969 | 0.001 | 0.019 | 0.000 | 0.992 | 0.023 | 0.1 | 0.11 | 2453 | 612 |
| BPp adolescence | 0.3 | 0.987 | 0.001 | 0.021 | 0.001 | 0.960 | 0.027 | 0.2 | 0.09 | 2390 | 596 |
| BPp adulthood | 0.3 | 0.985 | 0.001 | 0.023 | 0.002 | 0.965 | 0.036 | 0.4 | 0.04 | 2120 | 528 |
| Externalizing  childhood | 0.3 | 0.969 | 0.001 | 0.025 | 0.002 | 0.952 | 0.038 | 1.0 | 0.03 | 2453 | 612 |
| Externalizing adolescence | 0.3 | 0.996 | 0.001 | 0.022 | 0.001 | 0.951 | 0.033 | 0.5 | 0.04 | 2390 | 596 |
| Externalizing  adulthood | 0.3 | 0.984 | 0.002 | 0.026 | 0.002 | 0.981 | 0.022 | 0.1 | 0.11 | 2120 | 528 |
| Internalizing  childhood | 0.3 | 0.990 | 0.001 | 0.009 | 0.000 | 0.963 | 0.013 | 0.9 | 0.03 | 2453 | 612 |
| Internalizing adolescence | 0.3 | 0.985 | 0.001 | 0.020 | 0.001 | 1.009 | 0.007 | 0.1 | 0.04 | 2390 | 596 |
| Internalizing  adulthood | 0.3 | 0.981 | 0.001 | 0.016 | 0.001 | 0.971 | 0.059 | 0.6 | 0.04 | 2120 | 528 |
| **Cross-age-and-rater (cross-age approach) *** |  |  |  |  |  |  |  |  |  |  |  |
| BPp | 0.3 | 0.974 | 0.002 | 0.028 | 0.001 | 0.957 | 0.039 | 0.1 | 0.05 | 2453 | 612 |
| Externalizing | 0.3 | 0.974 | 0.002 | 0.036 | 0.002 | 0.926 | 0.034 | 0.6 | 0.03 | 2453 | 612 |
| Internalizing | 0.3 | 0.982 | 0.001 | 0.021 | 0.001 | 0.979 | 0.026 | 0.5 | 0.04 | 2453 | 612 |
| **Cross-age-and-rater (cross-rater approach) **** |  |  |  |  |  |  |  |  |  |  |  |
| BPp | 0.3 | 0.974 | 0.002 | 0.031 | 0.001 | 0.971 | 0.028 | 0.1 | 0.05 | 2453 | 612 |
| Externalizing | 0.3 | 0.968 | 0.002 | 0.033 | 0.002 | 0.987 | 0.039 | 0.8 | 0.02 | 2453 | **612** |
| Internalizing | 0.3 | 0.977 | 0.001 | 0.023 | 0.001 | 0.997 | 0.024 | 0.2 | 0.04 | 2453 | 612 |
| **Single-trait cross-age** |  |  |  |  |  |  |  |  |  |  |  |
| Hyperactivity  parent-rated | 0.3 | 0.985 | 0.001 | 0.019 | 0.002 | 0.968 | 0.003 | 0.6 | 0.05 | 2453 | 612 |
| Hyperactivity  teacher-rated | 0.3 | 0.993 | 0.002 | 0.030 | 0.000 | 0.985 | 0.022 | 0.1 | 0.06 | 2196 | 548 |
| Hyperactivity  child-rated | 0.3 | 0.992 | 0.001 | 0.016 | 0.001 | 0.986 | 0.008 | 0.8 | 0.04 | 2155 | 536 |
| Conduct parent-rated | 0.3 | 0.985 | 0.001 | 0.023 | 0.000 | 0.971 | 0.025 | 0.1 | 0.05 | 2453 | 612 |
| Conduct teacher-rated | 0.3 | 0.957 | 0.001 | 0.022 | 0.001 | 0.909 | 0.024 | 0.1 | 0.04 | 2364 | 589 |
| Conduct child-rated | 0.3 | 0.986 | 0.001 | 0.023 | 0.002 | 1.030 | 0.010 | 1 | 0.02 | 2390 | 596 |
| Emotional problems parent-rated | 0.3 | 0.989 | 0.001 | 0.012 | 0.000 | 0.976 | 0.005 | 0.3 | 0.08 | 2453 | 612 |
| Emotional problems teacher-rated | 0.3 | 0.996 | 0.001 | 0.009 | 0.000 | 0.964 | 0.025 | 0.2 | 0.06 | 2364 | 588 |
| Emotional problems child-rated | 0.3 | 0.989 | 0.001 | 0.013 | 0.001 | 1.014 | 0.014 | 0.3 | 0.08 | 2390 | 596 |
| Peer problems  parent-rated | 0.3 | 1.009 | 0.001 | 0.006 | 0.000 | 0.992 | 0.004 | 0.4 | 0.06 | 2453 | 612 |
| Peer problems teacher-rated | 0.3 | 1.008 | 0.001 | 0.003 | 0.000 | 1.010 | 0.008 | 0.5 | 0.05 | 2365 | 588 |
| Peer problems  child-rated | 0.3 | 1.003 | 0.001 | 0.007 | 0.000 | 1.021 | 0.002 | 0.4 | 0.05 | 2390 | 596 |
| **Single-trait cross-rater** |  |  |  |  |  |  |  |  |  |  |  |
| Hyperactivity  childhood | 0.3 | 0.980 | 0.001 | 0.023 | 0.001 | 0.968 | 0.016 | 0.1 | 0.05 | 2453 | 612 |
| Hyperactivity adolescence | 0.3 | 0.991 | 0.001 | 0.011 | 0.002 | 1.011 | 0.016 | 0.8 | 0.03 | 1948 | 484 |
| Hyperactivity  adulthood | 0.3 | 1.004 | 0.001 | 0.012 | 0.001 | 0.986 | 0.014 | 0.2 | 0.09 | 2120 | 528 |
| Conduct childhood | 0.3 | 0.980 | 0.001 | 0.018 | 0.001 | 0.973 | 0.051 | 0.6 | 0.04 | 2453 | 612 |
| Conduct adolescence | 0.3 | 0.991 | 0.001 | 0.020 | 0.001 | 1.008 | 0.033 | 0.1 | 0.08 | 2387 | 596 |
| Conduct adulthood | 0.3 | 0.985 | 0.001 | 0.015 | 0.001 | 1.001 | 0.010 | 0.6 | 0.04 | 2120 | 528 |
| Emotional problems childhood | 0.3 | 0.999 | 0.001 | 0.008 | 0.001 | 0.934 | 0.002 | 0.9 | 0.03 | 2453 | 612 |
| Emotional problems adolescence | 0.3 | 0.986 | 0.001 | 0.022 | 0.001 | 1.043 | 0.003 | 0.6 | 0.02 | 2388 | 596 |
| Emotional problems adulthood | 0.3 | 0.988 | 0.001 | 0.020 | 0.002 | 0.976 | 0.028 | 0.7 | 0.05 | 2119 | 528 |
| Peer problems childhood | 0.3 | 1.010 | 0.001 | 0.005 | 0.000 | 0.993 | 0.003 | 0.4 | 0.05 | 2448 | 612 |
| Peer problems adolescence | 0.3 | 1.019 | 0.001 | 0.005 | 0.000 | 0.984 | 0.000 | 0.8 | 0.05 | 2388 | 596 |
| Peer problems adulthood | 0.3 | 0.985 | 0.001 | 0.010 | 0.001 | 0.989 | 0.008 | 0.3 | 0.06 | 2120 | 528 |
| **Cross-age** |  |  |  |  |  |  |  |  |  |  |  |
| BPp parent-rated | 0.01 | 0.978 | 0.001 | 0.012 | 0.001 | 0.993 | 0.015 | 0.4 | 0.04 | 2453 | 612 |
| BPp teacher-rated | 0.01 | 0.984 | 0.001 | 0.014 | 0.001 | 0.942 | 0.050 | 0.6 | 0.04 | 2365 | 588 |
| BPp child-rated | 0.01 | 0.989 | 0.001 | 0.015 | 0.001 | 0.990 | 0.005 | 0.1 | 0.08 | 2390 | 596 |
| Externalizing  parent-rated | 0.01 | 0.982 | 0.001 | 0.015 | 0.001 | 0.959 | 0.026 | 0.8 | 0.02 | 2453 | 612 |
| Externalizing  teacher-rated | 0.01 | 0.962 | 0.001 | 0.025 | 0.001 | 1.000 | 0.024 | 0.5 | 0.05 | 2365 | 588 |
| Externalizing  child-rated | 0.01 | 0.988 | 0.000 | 0.011 | 0.001 | 0.995 | 0.021 | 0.1 | 0.06 | 2390 | 596 |
| Internalizing  parent-rated | 0.01 | 0.992 | 0.001 | 0.005 | 0.000 | 0.976 | 0.009 | 0.5 | 0.05 | 2453 | 612 |
| Internalizing  teacher-rated | 0.01 | 0.978 | 0.001 | 0.013 | 0.001 | 1.005 | 0.012 | 0.5 | 0.03 | 2365 | 588 |
| Internalizing  child-rated | 0.01 | 0.983 | 0.001 | 0.011 | 0.001 | 1.022 | 0.005 | 0.6 | 0.03 | 2390 | 596 |
| **Cross-rater** |  |  |  |  |  |  |  |  |  |  |  |
| BPp childhood | 0.01 | 0.973 | 0.001 | 0.012 | 0.001 | 0.992 | 0.027 | 0.4 | 0.04 | 2453 | 612 |
| BPp adolescence | 0.01 | 0.993 | 0.001 | 0.009 | 0.001 | 0.968 | 0.012 | 0.9 | 0.03 | 2390 | 596 |
| BPp adulthood | 0.01 | 0.990 | 0.001 | 0.014 | 0.001 | 0.976 | 0.011 | 0.8 | 0.03 | 2120 | 528 |
| Externalizing childhood | 0.01 | 0.972 | 0.001 | 0.020 | 0.002 | 0.960 | 0.019 | 1.0 | 0.02 | 2453 | 612 |
| Externalizing adolescence | 0.01 | 1.002 | 0.001 | 0.009 | 0.001 | 0.956 | 0.026 | 0.3 | 0.06 | 2390 | 596 |
| Externalizing adulthood | 0.01 | 0.988 | 0.001 | 0.019 | 0.001 | 0.993 | 0.003 | 0.6 | 0.02 | 2120 | 528 |
| Internalizing  childhood | 0.01 | 0.993 | 0.001 | 0.006 | 0.000 | 0.966 | 0.006 | 0.4 | 0.06 | 2453 | 612 |
| Internalizing adolescence | 0.01 | 0.992 | 0.001 | 0.007 | 0.001 | 1.008 | 0.005 | 0.6 | 0.02 | 2390 | 596 |
| Internalizing adulthood | 0.01 | 0.985 | 0.000 | 0.009 | 0.001 | 0.984 | 0.026 | 0.5 | 0.06 | 2120 | 528 |
| **Cross-age-and-rater (cross-age**  **approach) *** |  |  |  |  |  |  |  |  |  |  |  |
| BPp | 0.01 | 0.979 | 0.001 | 0.019 | 0.001 | 0.9645 | 0.026 | 1.0 | 0.02 | 2453 | 612 |
| Externalizing | 0.01 | 0.979 | 0.001 | 0.024 | 0.001 | 0.9310 | 0.024 | 0.6 | 0.02 | 2453 | 612 |
| Internalizing | 0.01 | 0.987 | 0.001 | 0.011 | 0.001 | 0.9840 | 0.015 | 0.4 | 0.04 | 2453 | 612 |
| **Cross-age-and-rater (cross-rater**  **approach) **** |  |  |  |  |  |  |  |  |  |  |  |
| BPp | 0.01 | 0.979 | 0.001 | 0.021 | 0.001 | 0.9805 | 0.008 | 1.0 | 0.02 | 2453 | 612 |
| Externalizing | 0.01 | 0.976 | 0.001 | 0.018 | 0.001 | 0.9915 | 0.038 | 0.6 | 0.04 | 2453 | 612 |
| Internalizing | 0.01 | 0.982 | 0.001 | 0.013 | 0.001 | 1.0065 | 0.004 | 0.9 | 0.04 | 2453 | 612 |
| **Single-trait cross-age** |  |  |  |  |  |  |  |  |  |  |  |
| Hyperactivity  parent-rated | 0.01 | 0.987 | 0.001 | 0.015 | 0.002 | 0.967 | 0.003 | 0.7 | 0.05 | 2453 | 612 |
| Hyperactivity  teacher-rated | 0.01 | 0.995 | 0.002 | 0.026 | 0.001 | 0.990 | 0.012 | 1.0 | 0.02 | 2196 | 548 |
| Hyperactivity  child-rated | 0.01 | 0.995 | 0.001 | 0.010 | 0.001 | 0.987 | 0.004 | 0.6 | 0.03 | 2155 | 536 |
| Conduct parent-rated | 0.01 | 0.989 | 0.001 | 0.015 | 0.001 | 0.968 | 0.042 | 0.6 | 0.04 | 2453 | 612 |
| Conduct teacher-rated | 0.01 | 0.961 | 0.001 | 0.014 | 0.001 | 0.910 | 0.035 | 0.8 | 0.04 | 2364 | 589 |
| Conduct child-rated | 0.01 | 0.992 | 0.000 | 0.011 | 0.001 | 1.035 | 0.002 | 0.1 | 0.06 | 2390 | 596 |
| Emotional problems parent-rated | 0.01 | 0.995 | 0.001 | 0.004 | 0.000 | 0.978 | 0.002 | 1.0 | 0.03 | 2453 | 612 |
| Emotional problems teacher-rated | 0.01 | 1.000 | 0.001 | 0.004 | 0.000 | 0.972 | 0.002 | 0.6 | 0.05 | 2364 | 588 |
| Emotional problems child-rated | 0.01 | 0.993 | 0.001 | 0.006 | 0.000 | 1.018 | 0.008 | 0.6 | 0.06 | 2390 | 596 |
| Peer problems parent-rated | 0.01 | 1.008 | 0.000 | 0.007 | 0.001 | 0.992 | 0.004 | 0.2 | 0.05 | 2453 | 612 |
| Peer problems teacher-rated | 0.01 | 1.007 | 0.001 | 0.004 | 0.001 | 1.010 | 0.007 | 1.0 | 0.02 | 2365 | 588 |
| Peer problems  child-rated | 0.01 | 1.002 | 0.001 | 0.007 | 0.000 | 1.023 | 0.000 | 0.6 | 0.03 | 2390 | 596 |
| **Single-trait cross-rater** |  |  |  |  |  |  |  |  |  |  |  |
| Hyperactivity childhood | 0.01 | 0.983 | 0.001 | 0.016 | 0.001 | 0.969 | 0.013 | 0.7 | 0.05 | 2453 | 612 |
| Hyperactivity adolescence | 0.01 | 0.993 | 0.001 | 0.008 | 0.001 | 1.015 | 0.008 | 0.6 | 0.07 | 1948 | 484 |
| Hyperactivity adulthood | 0.01 | 1.007 | 0.002 | 0.008 | 0.002 | 0.991 | 0.002 | 0.6 | 0.03 | 2120 | 528 |
| Conduct childhood | 0.01 | 0.982 | 0.001 | 0.013 | 0.001 | 0.974 | 0.048 | 0.2 | 0.07 | 2453 | 612 |
| Conduct adolescence | 0.01 | 0.995 | 0.001 | 0.012 | 0.001 | 1.022 | 0.005 | 0.4 | 0.03 | 2387 | 596 |
| Conduct adulthood | 0.01 | 0.989 | 0.001 | 0.006 | 0.001 | 1.000 | 0.014 | 0.8 | 0.03 | 2120 | 528 |
| Emotional problems childhood | 0.01 | 1.003 | 0.001 | 0.003 | 0.000 | 0.935 | 0.001 | 1.0 | 0.03 | 2453 | 612 |
| Emotional problems adolescence | 0.01 | 0.994 | 0.001 | 0.007 | 0.001 | 1.042 | 0.001 | 0.6 | 0.03 | 2388 | 596 |
| Emotional problems adulthood | 0.01 | 0.992 | 0.001 | 0.012 | 0.001 | 0.981 | 0.018 | 0.2 | 0.09 | 2119 | 528 |
| Peer problems childhood | 0.01 | 1.010 | 0.001 | 0.006 | 0.001 | 0.995 | 0.001 | 0.3 | 0.05 | 2448 | 612 |
| Peer problems adolescence | 0.01 | 1.017 | 0.001 | 0.008 | 0.001 | 0.985 | 0.000 | 0.6 | 0.07 | 2388 | 596 |
| Peer problems adulthood | 0.01 | 0.986 | 0.000 | 0.008 | 0.001 | 0.990 | 0.005 | 1.0 | 0.02 | 2120 | 528 |

| **Supporting table S3**. Behaviour problems composites: model fit indices and predictions from multiple regression. | | | | | | | |
| --- | --- | --- | --- | --- | --- | --- | --- |
| Note. | | | | | | | |
| FRCT= fraction of SNPs; SE= standard error; P= p-value; R2= variance explained. | | | | | | | |
| * cross-age approach: combined individual behaviour problems at all ages (2-21) for parent, teacher and child ratings to create the first order factors of cross-age externalizing and internalizing, which were then combined across raters to create the second-order factors of cross-age-and-rater externalizing and internalizing. | | | | | | | |
| ** cross-rater approach: combined individual behaviour problems in adolescence and adulthood to create the first order factors of cross-rater externalizing and internalizing, which were then combined across developmental stages to create the second-order factors of cross-age-and-rater externalizing and internalizing. | | | | | | | |
|  | **Multiple regression** | | | | | | |
|  | **FRCT** | **BETA** | **SE** | **T** | **P** | **Adjusted R2** | **N** |
| **Cross-age** |  |  |  |  |  |  |  |
| BPp parent-rated | 1 | 0.089 | 0.019 | 4.673 | 0.000 | 0.019 | 3065 |
| BPp teacher-rated | 1 | 0.127 | 0.019 | 6.575 | 0.000 | 0.022 | 2953 |
| BPp child-rated | 1 | 0.073 | 0.019 | 3.779 | 0.000 | 0.027 | 2986 |
| Externalizing parent-rated | 1 | 0.125 | 0.019 | 6.612 | 0.000 | 0.024 | 3065 |
| Externalizing teacher-rated | 1 | 0.147 | 0.019 | 7.633 | 0.000 | 0.031 | 2953 |
| Externalizing child-rated | 1 | 0.086 | 0.019 | 4.420 | 0.000 | 0.028 | 2986 |
| Internalizing parent-rated | 1 | 0.038 | 0.019 | 1.993 | 0.046 | 0.013 | 3065 |
| Internalizing teacher-rated | 1 | 0.093 | 0.020 | 4.776 | 0.000 | 0.012 | 2953 |
| Internalizing child-rated | 1 | 0.053 | 0.019 | 2.732 | 0.006 | 0.025 | 2986 |
| **Cross-rater** |  |  |  |  |  |  |  |
| BPp childhood | 1 | 0.111 | 0.019 | 5.866 | 0.000 | 0.020 | 3065 |
| BPp adolescence | 1 | 0.065 | 0.019 | 3.389 | 0.001 | 0.025 | 2986 |
| BPp adulthood | 1 | 0.079 | 0.020 | 3.873 | 0.000 | 0.027 | 2648 |
| Externalizing childhood | 1 | 0.146 | 0.019 | 7.799 | 0.000 | 0.028 | 3065 |
| Externalizing adolescence | 1 | 0.078 | 0.019 | 4.005 | 0.000 | 0.028 | 2986 |
| Externalizing adulthood | 1 | 0.092 | 0.020 | 4.491 | 0.000 | 0.027 | 2648 |
| Internalizing childhood | 1 | 0.056 | 0.019 | 2.937 | 0.003 | 0.011 | 3065 |
| Internalizing adolescence | 1 | 0.040 | 0.019 | 2.063 | 0.039 | 0.018 | 2986 |
| Internalizing adulthood | 1 | 0.060 | 0.020 | 2.946 | 0.003 | 0.025 | 2648 |
| **Cross-age-and-rater (cross-age approach) *** |  |  |  |  |  |  |  |
| BPp | 1 | 0.114 | 0.019 | 6.051 | 0.000 | 0.033 | 3065 |
| Externalizing | 1 | 0.143 | 0.019 | 7.620 | 0.000 | 0.038 | 3065 |
| Internalizing | 1 | 0.072 | 0.019 | 3.766 | 0.000 | 0.026 | 3065 |
| **Cross-age-and-rater (cross-rater approach) **** |  |  |  |  |  |  |  |
| BPp | 1 | 0.099 | 0.019 | 5.210 | 0.000 | 0.033 | 3065 |
| Externalizing | 1 | 0.121 | 0.019 | 6.403 | 0.000 | 0.037 | 3065 |
| Internalizing | 1 | 0.060 | 0.019 | 3.161 | 0.002 | 0.026 | 3065 |
| **Single-trait cross-age** |  |  |  |  |  |  |  |
| Hyperactivity parent-rated | 1 | 0.118 | 0.019 | 6.163 | 0.000 | 0.018 | 3065 |
| Hyperactivity teacher-rated | 1 | 0.159 | 0.020 | 7.798 | 0.000 | 0.032 | 2744 |
| Hyperactivity child-rated | 1 | 0.088 | 0.021 | 4.256 | 0.000 | 0.018 | 2691 |
| Conduct parent-rated | 1 | 0.113 | 0.019 | 5.925 | 0.000 | 0.024 | 3065 |
| Conduct teacher-rated | 1 | 0.108 | 0.019 | 5.729 | 0.000 | 0.022 | 2953 |
| Conduct child-rated | 1 | 0.058 | 0.020 | 2.980 | 0.003 | 0.023 | 2986 |
| Emotional problems parent-rated | 1 | 0.013 | 0.019 | 0.680 | 0.496 | 0.011 | 3065 |
| Emotional problems teacher-rated | 1 | 0.023 | 0.020 | 1.161 | 0.246 | 0.011 | 2952 |
| Emotional problems child-rated | 1 | 0.020 | 0.020 | 1.006 | 0.314 | 0.018 | 2986 |
| Peer problems parent-rated | 1 | 0.028 | 0.020 | 1.417 | 0.156 | 0.006 | 3065 |
| Peer problems teacher-rated | 1 | 0.051 | 0.020 | 2.563 | 0.010 | 0.002 | 2953 |
| Peer problems child-rated | 1 | 0.031 | 0.020 | 1.543 | 0.123 | 0.011 | 2986 |
| **Single-trait cross-rater** |  |  |  |  |  |  |  |
| Hyperactivity childhood | 1 | 0.133 | 0.019 | 7.003 | 0.000 | 0.022 | 3065 |
| Hyperactivity adolescence | 1 | 0.072 | 0.022 | 3.327 | 0.001 | 0.014 | 2432 |
| Hyperactivity adulthood | 1 | 0.074 | 0.021 | 3.552 | 0.000 | 0.013 | 2648 |
| Conduct childhood | 1 | 0.116 | 0.019 | 6.082 | 0.000 | 0.023 | 3065 |
| Conduct adolescence | 1 | 0.068 | 0.020 | 3.471 | 0.001 | 0.025 | 2983 |
| Conduct adulthood | 1 | 0.083 | 0.021 | 4.028 | 0.000 | 0.014 | 2648 |
| Emotional problems childhood | 1 | 0.012 | 0.019 | 0.641 | 0.522 | 0.007 | 3065 |
| Emotional problems adolescence | 1 | 0.018 | 0.020 | 0.899 | 0.369 | 0.015 | 2984 |
| Emotional problems adulthood | 1 | 0.024 | 0.021 | 1.179 | 0.239 | 0.024 | 2647 |
| Peer problems childhood | 1 | 0.023 | 0.020 | 1.181 | 0.238 | 0.003 | 3060 |
| Peer problems adolescence | 1 | 0.033 | 0.020 | 1.647 | 0.100 | 0.008 | 2984 |
| Peer problems adulthood | 1 | 0.046 | 0.021 | 2.267 | 0.023 | 0.012 | 2648 |
| **Cross-age** |  |  |  |  |  |  |  |
| BPp parent-rated | 0.3 | 0.093 | 0.019 | 4.847 | 0.000 | 0.019 | 3065 |
| BPp teacher-rated | 0.3 | 0.135 | 0.019 | 6.957 | 0.000 | 0.026 | 2953 |
| BPp child-rated | 0.3 | 0.074 | 0.019 | 3.793 | 0.000 | 0.026 | 2986 |
| Externalizing parent-rated | 0.3 | 0.129 | 0.019 | 6.815 | 0.000 | 0.024 | 3065 |
| Externalizing teacher-rated | 0.3 | 0.154 | 0.019 | 8.015 | 0.000 | 0.033 | 2953 |
| Externalizing child-rated | 0.3 | 0.086 | 0.019 | 4.422 | 0.000 | 0.028 | 2986 |
| Internalizing parent-rated | 0.3 | 0.040 | 0.019 | 2.107 | 0.035 | 0.012 | 3065 |
| Internalizing teacher-rated | 0.3 | 0.100 | 0.020 | 5.112 | 0.000 | 0.016 | 2953 |
| Internalizing child-rated | 0.3 | 0.054 | 0.019 | 2.759 | 0.006 | 0.022 | 2986 |
| **Cross-rater** |  |  |  |  |  |  |  |
| BPp childhood | 0.3 | 0.115 | 0.019 | 6.058 | 0.000 | 0.020 | 3065 |
| BPp adolescence | 0.3 | 0.069 | 0.019 | 3.583 | 0.000 | 0.024 | 2986 |
| BPp adulthood | 0.3 | 0.079 | 0.021 | 3.850 | 0.000 | 0.027 | 2648 |
| Externalizing childhood | 0.3 | 0.151 | 0.019 | 8.036 | 0.000 | 0.028 | 3065 |
| Externalizing adolescence | 0.3 | 0.080 | 0.019 | 4.121 | 0.000 | 0.026 | 2986 |
| Externalizing adulthood | 0.3 | 0.093 | 0.021 | 4.513 | 0.000 | 0.026 | 2648 |
| Internalizing childhood | 0.3 | 0.059 | 0.019 | 3.050 | 0.002 | 0.010 | 3065 |
| Internalizing adolescence | 0.3 | 0.045 | 0.020 | 2.295 | 0.022 | 0.017 | 2986 |
| Internalizing adulthood | 0.3 | 0.059 | 0.020 | 2.879 | 0.004 | 0.025 | 2648 |
| **Cross-age-and-rater (cross-age approach) *** |  |  |  |  |  |  |  |
| BPp | 0.3 | 0.119 | 0.019 | 6.263 | 0.000 | 0.033 | 3065 |
| Externalizing | 0.3 | 0.147 | 0.019 | 7.846 | 0.000 | 0.038 | 3065 |
| Internalizing | 0.3 | 0.075 | 0.019 | 3.938 | 0.000 | 0.025 | 3065 |
| **Cross-age-and-rater (cross-rater approach) **** |  |  |  |  |  |  |  |
| BPp | 0.3 | 0.102 | 0.019 | 5.381 | 0.000 | 0.032 | 3065 |
| Externalizing | 0.3 | 0.125 | 0.019 | 6.571 | 0.000 | 0.036 | 3065 |
| Internalizing | 0.3 | 0.063 | 0.019 | 3.311 | 0.001 | 0.025 | 3065 |
| **Single-trait cross-age** |  |  |  |  |  |  |  |
| Hyperactivity parent-rated | 0.3 | 0.122 | 0.019 | 6.359 | 0.000 | 0.018 | 3065 |
| Hyperactivity teacher-rated | 0.3 | 0.164 | 0.020 | 8.051 | 0.000 | 0.032 | 2744 |
| Hyperactivity child-rated | 0.3 | 0.087 | 0.021 | 4.236 | 0.000 | 0.018 | 2691 |
| Conduct parent-rated | 0.3 | 0.116 | 0.019 | 6.064 | 0.000 | 0.025 | 3065 |
| Conduct teacher-rated | 0.3 | 0.115 | 0.019 | 6.094 | 0.000 | 0.023 | 2953 |
| Conduct child-rated | 0.3 | 0.057 | 0.020 | 2.921 | 0.004 | 0.022 | 2986 |
| Emotional problems parent-rated | 0.3 | 0.014 | 0.019 | 0.747 | 0.455 | 0.010 | 3065 |
| Emotional problems teacher-rated | 0.3 | 0.025 | 0.020 | 1.268 | 0.205 | 0.012 | 2952 |
| Emotional problems child-rated | 0.3 | 0.017 | 0.020 | 0.885 | 0.376 | 0.013 | 2986 |
| Peer problems parent-rated | 0.3 | 0.030 | 0.020 | 1.524 | 0.128 | 0.005 | 3065 |
| Peer problems teacher-rated | 0.3 | 0.056 | 0.020 | 2.805 | 0.005 | 0.003 | 2953 |
| Peer problems child-rated | 0.3 | 0.032 | 0.020 | 1.617 | 0.106 | 0.009 | 2986 |
| **Single-trait cross-rater** |  |  |  |  |  |  |  |
| Hyperactivity childhood | 0.3 | 0.137 | 0.019 | 7.206 | 0.000 | 0.022 | 3065 |
| Hyperactivity adolescence | 0.3 | 0.076 | 0.022 | 3.483 | 0.001 | 0.013 | 2432 |
| Hyperactivity adulthood | 0.3 | 0.076 | 0.021 | 3.646 | 0.000 | 0.012 | 2648 |
| Conduct childhood | 0.3 | 0.119 | 0.019 | 6.254 | 0.000 | 0.024 | 3065 |
| Conduct adolescence | 0.3 | 0.069 | 0.020 | 3.505 | 0.000 | 0.025 | 2983 |
| Conduct adulthood | 0.3 | 0.085 | 0.021 | 4.093 | 0.000 | 0.013 | 2648 |
| Emotional problems childhood | 0.3 | 0.013 | 0.019 | 0.666 | 0.506 | 0.006 | 3065 |
| Emotional problems adolescence | 0.3 | 0.022 | 0.020 | 1.126 | 0.260 | 0.016 | 2984 |
| Emotional problems adulthood | 0.3 | 0.022 | 0.021 | 1.047 | 0.295 | 0.023 | 2647 |
| Peer problems childhood | 0.3 | 0.026 | 0.020 | 1.309 | 0.191 | 0.003 | 3060 |
| Peer problems adolescence | 0.3 | 0.037 | 0.020 | 1.847 | 0.065 | 0.007 | 2984 |
| Peer problems adulthood | 0.3 | 0.045 | 0.021 | 2.185 | 0.029 | 0.011 | 2648 |
| **Cross-age** |  |  |  |  |  |  |  |
| BPp parent-rated | 0.01 | 0.091 | 0.018 | 4.961 | 0.000 | 0.012 | 3065 |
| BPp teacher-rated | 0.01 | 0.123 | 0.019 | 6.657 | 0.000 | 0.020 | 2953 |
| BPp child-rated | 0.01 | 0.074 | 0.019 | 3.957 | 0.000 | 0.014 | 2986 |
| Externalizing parent-rated | 0.01 | 0.117 | 0.018 | 6.423 | 0.000 | 0.018 | 3065 |
| Externalizing teacher-rated | 0.01 | 0.137 | 0.018 | 7.429 | 0.000 | 0.024 | 2953 |
| Externalizing child-rated | 0.01 | 0.082 | 0.019 | 4.382 | 0.000 | 0.014 | 2986 |
| Internalizing parent-rated | 0.01 | 0.049 | 0.018 | 2.668 | 0.008 | 0.004 | 3065 |
| Internalizing teacher-rated | 0.01 | 0.096 | 0.019 | 5.130 | 0.000 | 0.013 | 2953 |
| Internalizing child-rated | 0.01 | 0.058 | 0.019 | 3.104 | 0.002 | 0.011 | 2986 |
| **Cross-rater** |  |  |  |  |  |  |  |
| BPp childhood | 0.01 | 0.104 | 0.018 | 5.675 | 0.000 | 0.014 | 3065 |
| BPp adolescence | 0.01 | 0.079 | 0.019 | 4.219 | 0.000 | 0.010 | 2986 |
| BPp adulthood | 0.01 | 0.081 | 0.020 | 4.065 | 0.000 | 0.016 | 2648 |
| Externalizing childhood | 0.01 | 0.132 | 0.018 | 7.267 | 0.000 | 0.021 | 3065 |
| Externalizing adolescence | 0.01 | 0.082 | 0.019 | 4.348 | 0.000 | 0.012 | 2986 |
| Externalizing adulthood | 0.01 | 0.093 | 0.020 | 4.652 | 0.000 | 0.015 | 2648 |
| Internalizing childhood | 0.01 | 0.057 | 0.018 | 3.096 | 0.002 | 0.004 | 3065 |
| Internalizing adolescence | 0.01 | 0.061 | 0.019 | 3.209 | 0.001 | 0.006 | 2986 |
| Internalizing adulthood | 0.01 | 0.062 | 0.020 | 3.151 | 0.002 | 0.015 | 2648 |
| **Cross-age-and-rater (cross-age approach) *** |  |  |  |  |  |  |  |
| BPp | 0.01 | 0.116 | 0.018 | 6.347 | 0.000 | 0.021 | 3065 |
| Externalizing | 0.01 | 0.136 | 0.018 | 7.493 | 0.000 | 0.026 | 3065 |
| Internalizing | 0.01 | 0.082 | 0.018 | 4.450 | 0.000 | 0.013 | 3065 |
| **Cross-age-and-rater (cross-rater approach) **** |  |  |  |  |  |  |  |
| BPp | 0.01 | 0.106 | 0.018 | 5.800 | 0.000 | 0.019 | 3065 |
| Externalizing | 0.01 | 0.122 | 0.018 | 6.662 | 0.000 | 0.022 | 3065 |
| Internalizing | 0.01 | 0.074 | 0.018 | 4.029 | 0.000 | 0.012 | 3065 |
| **Single-trait cross-age** |  |  |  |  |  |  |  |
| Hyperactivity parent-rated | 0.01 | 0.111 | 0.018 | 6.039 | 0.000 | 0.012 | 3065 |
| Hyperactivity teacher-rated | 0.01 | 0.140 | 0.020 | 7.153 | 0.000 | 0.021 | 2744 |
| Hyperactivity child-rated | 0.01 | 0.086 | 0.020 | 4.306 | 0.000 | 0.007 | 2691 |
| Conduct parent-rated | 0.01 | 0.098 | 0.018 | 5.316 | 0.000 | 0.019 | 3065 |
| Conduct teacher-rated | 0.01 | 0.106 | 0.018 | 5.856 | 0.000 | 0.018 | 2953 |
| Conduct child-rated | 0.01 | 0.042 | 0.019 | 2.194 | 0.028 | 0.011 | 2986 |
| Emotional problems parent-rated | 0.01 | 0.021 | 0.019 | 1.132 | 0.258 | 0.000 | 3065 |
| Emotional problems teacher-rated | 0.01 | 0.041 | 0.019 | 2.189 | 0.029 | 0.002 | 2952 |
| Emotional problems child-rated | 0.01 | 0.017 | 0.019 | 0.890 | 0.374 | 0.004 | 2986 |
| Peer problems parent-rated | 0.01 | 0.044 | 0.019 | 2.317 | 0.021 | 0.006 | 3065 |
| Peer problems teacher-rated | 0.01 | 0.051 | 0.019 | 2.656 | 0.008 | 0.004 | 2953 |
| Peer problems child-rated | 0.01 | 0.046 | 0.019 | 2.423 | 0.015 | 0.007 | 2986 |
| **Single-trait cross-rater** |  |  |  |  |  |  |  |
| Hyperactivity childhood | 0.01 | 0.122 | 0.018 | 6.626 | 0.000 | 0.015 | 3065 |
| Hyperactivity adolescence | 0.01 | 0.086 | 0.021 | 4.065 | 0.000 | 0.006 | 2432 |
| Hyperactivity adulthood | 0.01 | 0.079 | 0.020 | 3.921 | 0.000 | 0.005 | 2648 |
| Conduct childhood | 0.01 | 0.100 | 0.018 | 5.448 | 0.000 | 0.019 | 3065 |
| Conduct adolescence | 0.01 | 0.060 | 0.019 | 3.167 | 0.002 | 0.012 | 2983 |
| Conduct adulthood | 0.01 | 0.071 | 0.020 | 3.572 | 0.000 | 0.007 | 2648 |
| Emotional problems childhood | 0.01 | 0.012 | 0.019 | 0.671 | 0.502 | <.001 | 3065 |
| Emotional problems adolescence | 0.01 | 0.034 | 0.019 | 1.791 | 0.073 | 0.004 | 2984 |
| Emotional problems adulthood | 0.01 | 0.030 | 0.020 | 1.520 | 0.129 | 0.015 | 2647 |
| Peer problems childhood | 0.01 | 0.038 | 0.019 | 2.011 | 0.044 | 0.004 | 3060 |
| Peer problems adolescence | 0.01 | 0.052 | 0.019 | 2.698 | 0.007 | 0.006 | 2984 |
| Peer problems adulthood | 0.01 | 0.048 | 0.020 | 2.400 | 0.016 | 0.008 | 2648 |

| **Supporting table S4**. Behaviour problems composites: model fit indices and predictions from multiple regression, for males and females separately. | | | | | | | |
| --- | --- | --- | --- | --- | --- | --- | --- |
| Note. | | | | | | | |
| FRCT= fraction of SNPs; SE= standard error; P= p-value; R2= variance explained. | | | | | | | |
| **Composites** | **Multiple regression: males only** | | | | | | |
|  | **FRCT** | **BETA** | **SE** | **T** | **P** | **Adjusted R2** | **N** |
| **Cross-age** |  |  |  |  |  |  |  |
| BPp parent-rated | 1 | 0.091 | 0.029 | 3.107 | 0.002 | 0.014 | 1376 |
| BPp teacher-rated | 1 | 0.179 | 0.032 | 5.595 | 0.000 | 0.035 | 1322 |
| BPp child-rated | 1 | 0.053 | 0.028 | 1.914 | 0.056 | 0.012 | 1332 |
| Externalizing parent-rated | 1 | 0.118 | 0.030 | 3.974 | 0.000 | 0.015 | 1376 |
| Externalizing teacher-rated | 1 | 0.200 | 0.033 | 6.069 | 0.000 | 0.043 | 1322 |
| Externalizing child-rated | 1 | 0.066 | 0.028 | 2.340 | 0.019 | 0.017 | 1332 |
| Internalizing parent-rated | 1 | 0.049 | 0.029 | 1.676 | 0.094 | 0.013 | 1376 |
| Internalizing teacher-rated | 1 | 0.139 | 0.031 | 4.444 | 0.000 | 0.021 | 1322 |
| Internalizing child-rated | 1 | 0.035 | 0.028 | 1.275 | 0.203 | 0.008 | 1332 |
| **Cross-rater** |  |  |  |  |  |  |  |
| BPp childhood | 1 | 0.129 | 0.030 | 4.309 | 0.000 | 0.020 | 1376 |
| BPp adolescence | 1 | 0.053 | 0.029 | 1.845 | 0.065 | 0.011 | 1336 |
| BPp adulthood | 1 | 0.056 | 0.030 | 1.865 | 0.062 | 0.015 | 1126 |
| Externalizing childhood | 1 | 0.157 | 0.030 | 5.206 | 0.000 | 0.025 | 1376 |
| Externalizing adolescence | 1 | 0.063 | 0.029 | 2.172 | 0.030 | 0.015 | 1336 |
| Externalizing adulthood | 1 | 0.062 | 0.030 | 2.049 | 0.041 | 0.017 | 1126 |
| Internalizing childhood | 1 | 0.077 | 0.030 | 2.606 | 0.009 | 0.013 | 1376 |
| Internalizing adolescence | 1 | 0.033 | 0.029 | 1.125 | 0.261 | 0.009 | 1336 |
| Internalizing adulthood | 1 | 0.044 | 0.029 | 1.512 | 0.131 | 0.012 | 1126 |
| **Composites** | **Multiple regression: females only** | | | | | | |
|  | **Fraction of SNPs** | **BETA** | **SE** | **T** | **P** | **Adjusted R2** | **N** |
| **Cross-age** |  |  |  |  |  |  |  |
| BPp parent-rated |  | 0.087 | 0.025 | 3.473 | 0.001 | 0.023 | 1689 |
| BPp teacher-rated | 1 | 0.087 | 0.023 | 3.707 | 0.000 | 0.021 | 1631 |
| BPp child-rated | 1 | 0.088 | 0.027 | 3.259 | 0.001 | 0.039 | 1654 |
| Externalizing parent-rated | 1 | 0.131 | 0.024 | 5.354 | 0.000 | 0.031 | 1689 |
| Externalizing teacher-rated | 1 | 0.107 | 0.022 | 4.804 | 0.000 | 0.029 | 1631 |
| Externalizing child-rated | 1 | 0.100 | 0.027 | 3.725 | 0.000 | 0.036 | 1654 |
| Internalizing parent-rated | 1 | 0.029 | 0.025 | 1.139 | 0.255 | 0.013 | 1689 |
| Internalizing teacher-rated | 1 | 0.057 | 0.025 | 2.295 | 0.022 | 0.013 | 1631 |
| Internalizing child-rated | 1 | 0.067 | 0.027 | 2.452 | 0.014 | 0.038 | 1654 |
| **Cross-rater** |  |  |  |  |  |  |  |
| BPp childhood | 1 | 0.096 | 0.024 | 3.956 | 0.000 | 0.020 | 1689 |
| BPp adolescence | 1 | 0.072 | 0.026 | 2.785 | 0.005 | 0.037 | 1650 |
| BPp adulthood | 1 | 0.100 | 0.028 | 3.571 | 0.000 | 0.035 | 1522 |
| Externalizing childhood | 1 | 0.138 | 0.024 | 5.843 | 0.000 | 0.033 | 1689 |
| Externalizing adolescence | 1 | 0.087 | 0.026 | 3.323 | 0.001 | 0.036 | 1650 |
| Externalizing adulthood | 1 | 0.118 | 0.028 | 4.235 | 0.000 | 0.032 | 1522 |
| Internalizing childhood | 1 | 0.038 | 0.025 | 1.511 | 0.131 | 0.008 | 1689 |
| Internalizing adolescence | 1 | 0.044 | 0.026 | 1.672 | 0.095 | 0.028 | 1650 |
| Internalizing adulthood | 1 | 0.075 | 0.028 | 2.650 | 0.008 | 0.036 | 1522 |

| **Supporting table S5**. SDQ scales (observed traits): model fit indices and predictions from elastic net regularization. | | | | | | | | | | | | |
| --- | --- | --- | --- | --- | --- | --- | --- | --- | --- | --- | --- | --- |
| Note. | | | | | | | | | | | | |
| CV= cross-validated; SD= standard deviation; R2= variance explained; RMSE= root mean square error; train= training set (80%); test= hold-out set (20%). | | | | | | | | | | | | |
| **Trait** | **Rater** | **Elastic net regularization** | | | | | | | | | | |
|  |  | **FRCT** | **Mean CV**  **RMSE (train)** | **SD CV**  **RMSE (train)** | **Mean CV**  **R2 (train)** | **SD CV**  **R2 (train)** | **RMSE** | **R2** | **Alpha** | **Lambda** | **N (train)** | **N (test)** |
| Year 2 |  |  |  |  |  |  |  |  |  |  |  |  |
| Hyperactivity | Parent | 1 | 1.005 | 0.001 | 0.005 | 0.000 | 1.009 | 0.007 | 1 | 0.040 | 2182 | 544 |
| Conduct | Parent | 1 | 0.989 | 0.001 | 0.005 | 0.001 | 0.955 | 0.001 | 0.5 | 0.061 | 2178 | 543 |
| Emotional problems | Parent | 1 | 0.987 | 0.002 | 0.005 | 0.001 | 0.984 | 0.000 | 0.5 | 0.064 | 2186 | 544 |
| Year 3 |  |  |  |  |  |  |  |  |  |  |  |  |
| Hyperactivity | Parent | 1 | 1.006 | 0.001 | 0.007 | 0.001 | 1.004 | 0.001 | 0.4 | 0.065 | 2237 | 557 |
| Conduct | Parent | 1 | 0.985 | 0.001 | 0.016 | 0.000 | 0.988 | 0.003 | 0.1 | 0.089 | 2234 | 556 |
| Emotional problems | Parent | 1 | 0.990 | 0.001 | 0.005 | 0.001 | 1.005 | 0.014 | 0.5 | 0.063 | 2236 | 557 |
| Year 4 |  |  |  |  |  |  |  |  |  |  |  |  |
| Hyperactivity | Parent | 1 | 0.986 | 0.000 | 0.010 | 0.000 | 0.998 | 0.008 | 0.2 | 0.070 | 3176 | 792 |
| Conduct | Parent | 1 | 0.969 | 0.000 | 0.009 | 0.000 | 0.968 | 0.011 | 0.6 | 0.029 | 3176 | 791 |
| Emotional problems | Parent | 1 | 1.005 | 0.001 | 0.004 | 0.001 | 0.990 | 0.001 | 0.9 | 0.024 | 3176 | 791 |
| Peer problems | Parent | 1 | 1.002 | 0.001 | 0.003 | 0.000 | 0.996 | 0.000 | 1 | 0.032 | 3171 | 791 |
| Year 7 |  |  |  |  |  |  |  |  |  |  |  |  |
| Hyperactivity | Parent | 1 | 0.983 | 0.001 | 0.017 | 0.000 | 0.952 | 0.017 | 0.1 | 0.046 | 3251 | 812 |
| Hyperactivity | Teacher | 1 | 0.973 | 0.002 | 0.026 | 0.001 | 1.004 | 0.013 | 0.6 | 0.023 | 2695 | 672 |
| Conduct | Parent | 1 | 0.982 | 0.001 | 0.021 | 0.001 | 0.992 | 0.006 | 1 | 0.020 | 3253 | 812 |
| Conduct | Teacher | 1 | 1.276 | 0.001 | 0.018 | 0.001 | 1.277 | 0.004 | 0.5 | 0.049 | 2699 | 673 |
| Emotional problems | Parent | 1 | 0.976 | 0.000 | 0.009 | 0.001 | 1.017 | 0.003 | 0.1 | 0.054 | 3252 | 812 |
| Emotional problems | Teacher | 1 | 0.994 | 0.001 | 0.005 | 0.000 | 0.995 | 0.009 | 0.5 | 0.056 | 2684 | 669 |
| Peer problems | Parent | 1 | 1.002 | 0.001 | 0.003 | 0.001 | 0.995 | 0.000 | 0.6 | 0.047 | 3253 | 811 |
| Peer problems | Teacher | 1 | 0.991 | 0.001 | 0.006 | 0.001 | 1.015 | 0.000 | 0.2 | 0.056 | 2690 | 672 |
| Year 9 |  |  |  |  |  |  |  |  |  |  |  |  |
| Hyperactivity | Parent | 1 | 0.991 | 0.001 | 0.027 | 0.001 | 0.988 | 0.000 | 0.2 | 0.054 | 1551 | 384 |
| Hyperactivity | Teacher | 1 | 0.994 | 0.002 | 0.029 | 0.004 | 0.987 | 0.023 | 0.6 | 0.065 | 1283 | 320 |
| Hyperactivity | Child | 1 | 1.007 | 0.001 | 0.009 | 0.001 | 0.980 | 0.002 | 1 | 0.033 | 1532 | 380 |
| Conduct | Parent | 1 | 1.004 | 0.001 | 0.018 | 0.001 | 1.004 | 0.008 | 0.3 | 0.103 | 1551 | 385 |
| Conduct | Teacher | 1 | 0.936 | 0.002 | 0.032 | 0.003 | 0.909 | 0.000 | 1 | 0.026 | 1284 | 320 |
| Conduct | Child | 1 | 1.000 | 0.001 | 0.012 | 0.001 | 0.948 | 0.006 | 0.9 | 0.038 | 1529 | 380 |
| Peer problems | Parent | 1 | 1.013 | 0.002 | 0.008 | 0.001 | 0.997 | 0.002 | 0.3 | 0.063 | 1552 | 384 |
| Peer problems | Teacher | 1 | 0.997 | 0.002 | 0.006 | 0.000 | 0.995 | 0.002 | 1 | 0.045 | 1288 | 319 |
| Peer problems | Child | 1 | 0.998 | 0.002 | 0.008 | 0.001 | 0.960 | 0.004 | 0.3 | 0.080 | 1525 | 380 |
| Emotional problems | Parent | 1 | 1.004 | 0.002 | 0.006 | 0.000 | 0.986 | 0.010 | 1 | 0.053 | 1552 | 385 |
| Emotional problems | Teacher | 1 | 0.983 | 0.001 | 0.013 | 0.001 | 1.021 | 0.002 | 0.3 | 0.081 | 1282 | 320 |
| Emotional problems | Child | 1 | 1.002 | 0.001 | 0.008 | 0.000 | 0.989 | 0.033 | 0.6 | 0.065 | 1529 | 380 |
| Year 12 |  |  |  |  |  |  |  |  |  |  |  |  |
| Conduct | Teacher | 1 | 0.981 | 0.001 | 0.013 | 0.001 | 0.997 | 0.012 | 0.9 | 0.034 | 2272 | 565 |
| Conduct | Child | 1 | 0.990 | 0.001 | 0.012 | 0.001 | 0.974 | 0.018 | 0.3 | 0.032 | 2752 | 688 |
| Emotional problems | Parent | 1 | 0.997 | 0.001 | 0.007 | 0.000 | 1.029 | 0.000 | 0.4 | 0.071 | 2760 | 687 |
| Emotional problems | Teacher | 1 | 0.985 | 0.001 | 0.012 | 0.002 | 0.978 | 0.004 | 0.1 | 0.062 | 2265 | 565 |
| Emotional problems | Child | 1 | 0.998 | 0.001 | 0.016 | 0.001 | 0.982 | 0.005 | 1 | 0.020 | 2752 | 686 |
| Peer problems | Parent | 1 | 0.997 | 0.001 | 0.004 | 0.000 | 0.972 | 0.007 | 0.5 | 0.040 | 2759 | 688 |
| Peer problems | Teacher | 1 | 0.995 | 0.001 | 0.005 | 0.000 | 1.019 | 0.001 | 0.5 | 0.050 | 2271 | 566 |
| Peer problems | Child | 1 | 1.007 | 0.001 | 0.004 | 0.000 | 1.020 | 0.003 | 0.3 | 0.047 | 2753 | 687 |
| Year 16 |  |  |  |  |  |  |  |  |  |  |  |  |
| Hyperactivity | Child | 1 | 0.996 | 0.001 | 0.011 | 0.000 | 0.969 | 0.009 | 0.6 | 0.031 | 2388 | 596 |
| Hyperactivity | Parent | 1 | 0.992 | 0.001 | 0.009 | 0.001 | 0.990 | 0.007 | 0.2 | 0.067 | 2395 | 597 |
| Conduct | Child | 1 | 0.995 | 0.001 | 0.023 | 0.001 | 0.987 | 0.013 | 0.5 | 0.045 | 2389 | 595 |
| Conduct | Parent | 1 | 0.984 | 0.001 | 0.015 | 0.002 | 1.004 | 0.016 | 0.1 | 0.068 | 2400 | 598 |
| Emotional problems | Child | 1 | 1.005 | 0.001 | 0.011 | 0.001 | 0.999 | 0.014 | 0.7 | 0.037 | 2388 | 596 |
| Peer problems | Child | 1 | 0.998 | 0.000 | 0.009 | 0.001 | 1.029 | 0.001 | 0.3 | 0.052 | 2388 | 596 |
| Year 21 |  |  |  |  |  |  |  |  |  |  |  |  |
| Hyperactivity | Parent | 1 | 1.009 | 0.001 | 0.014 | 0.001 | 0.977 | 0.004 | 0.6 | 0.035 | 2496 | 622 |
| Hyperactivity | Child | 1 | 0.994 | 0.001 | 0.018 | 0.001 | 0.993 | 0.016 | 0.8 | 0.016 | 2240 | 557 |
| Conduct | Parent | 1 | 0.990 | 0.001 | 0.010 | 0.001 | 0.981 | 0.025 | 0.8 | 0.029 | 2496 | 623 |
| Conduct | Child | 1 | 0.977 | 0.001 | 0.017 | 0.002 | 0.998 | 0.002 | 1 | 0.015 | 2240 | 558 |
| Emotional problems | Parent | 1 | 0.989 | 0.001 | 0.016 | 0.001 | 1.005 | 0.021 | 0.2 | 0.100 | 2496 | 621 |
| Emotional problems | Child | 1 | 0.987 | 0.001 | 0.015 | 0.000 | 1.011 | 0.024 | 0.6 | 0.040 | 2240 | 557 |
| Peer problems | Parent | 1 | 0.985 | 0.001 | 0.010 | 0.001 | 0.972 | 0.012 | 0.2 | 0.067 | 2495 | 622 |
| Peer problems | Child | 1 | 0.985 | 0.001 | 0.023 | 0.002 | 0.965 | 0.005 | 0.1 | 0.099 | 2240 | 557 |
| Year 2 |  |  |  |  |  |  |  |  |  |  |  |  |
| Hyperactivity | Parent | 0.3 | 1.005 | 0.001 | 0.004 | 0.000 | 1.009 | 0.008 | 1.0 | 0.041 | 2182 | 544 |
| Conduct | Parent | 0.3 | 0.989 | 0.002 | 0.005 | 0.001 | 0.956 | 0.001 | 0.5 | 0.062 | 2178 | 543 |
| Emotional problems | Parent | 0.3 | 0.987 | 0.001 | 0.004 | 0.001 | 0.983 | 0.002 | 0.6 | 0.056 | 2186 | 544 |
| Year 3 |  |  |  |  |  |  |  |  |  |  |  |  |
| Hyperactivity | Parent | 0.3 | 1.006 | 0.001 | 0.007 | 0.000 | 1.004 | 0.002 | 0.5 | 0.066 | 2237 | 557 |
| Conduct | Parent | 0.3 | 0.986 | 0.001 | 0.015 | 0.000 | 0.988 | 0.003 | 0.1 | 0.089 | 2234 | 556 |
| Emotional problems | Parent | 0.3 | 0.991 | 0.001 | 0.004 | 0.000 | 1.007 | 0.010 | 0.6 | 0.054 | 2236 | 557 |
| Year 4 |  |  |  |  |  |  |  |  |  |  |  |  |
| Hyperactivity | Parent | 0.3 | 0.986 | 0.000 | 0.010 | 0.000 | 0.998 | 0.008 | 0.2 | 0.071 | 3176 | 792 |
| Conduct | Parent | 0.3 | 0.969 | 0.001 | 0.008 | 0.000 | 0.968 | 0.011 | 0.6 | 0.029 | 3176 | 791 |
| Emotional problems | Parent | 0.3 | 1.005 | 0.001 | 0.004 | 0.001 | 0.991 | 0.000 | 0.3 | 0.056 | 3176 | 791 |
| Peer problems | Parent | 0.3 | 1.002 | 0.001 | 0.003 | 0.000 | 0.996 | 0.000 | 1.0 | 0.032 | 3171 | 791 |
| Year 7 |  |  |  |  |  |  |  |  |  |  |  |  |
| Hyperactivity | Parent | 0.3 | 0.983 | 0.001 | 0.016 | 0.001 | 0.954 | 0.016 | 0.8 | 0.046 | 3251 | 812 |
| Hyperactivity | Teacher | 0.3 | 0.973 | 0.002 | 0.026 | 0.001 | 1.004 | 0.014 | 0.6 | 0.023 | 2695 | 672 |
| Conduct | Parent | 0.3 | 0.982 | 0.001 | 0.020 | 0.001 | 0.992 | 0.006 | 1.0 | 0.020 | 3253 | 812 |
| Conduct | Teacher | 0.3 | 1.276 | 0.001 | 0.017 | 0.001 | 1.275 | 0.005 | 0.5 | 0.050 | 2699 | 673 |
| Emotional problems | Parent | 0.3 | 0.976 | 0.000 | 0.009 | 0.001 | 1.017 | 0.002 | 0.1 | 0.053 | 3252 | 812 |
| Emotional problems | Teacher | 0.3 | 0.994 | 0.001 | 0.005 | 0.000 | 0.995 | 0.006 | 0.5 | 0.053 | 2684 | 669 |
| Peer problems | Parent | 0.3 | 1.002 | 0.001 | 0.003 | 0.001 | 0.995 | 0.000 | 0.5 | 0.047 | 3253 | 811 |
| Peer problems | Teacher | 0.3 | 0.991 | 0.001 | 0.006 | 0.000 | 1.015 | 0.000 | 0.6 | 0.024 | 2690 | 672 |
| Year 9 |  |  |  |  |  |  |  |  |  |  |  |  |
| Hyperactivity | Parent | 0.3 | 0.991 | 0.001 | 0.028 | 0.000 | 0.989 | 0.000 | 0.2 | 0.055 | 1551 | 384 |
| Hyperactivity | Teacher | 0.3 | 0.994 | 0.002 | 0.029 | 0.004 | 0.986 | 0.023 | 0.6 | 0.065 | 1283 | 320 |
| Hyperactivity | Child | 0.3 | 1.007 | 0.002 | 0.009 | 0.001 | 0.980 | 0.002 | 0.8 | 0.034 | 1532 | 380 |
| Conduct | Parent | 0.3 | 1.002 | 0.001 | 0.020 | 0.001 | 1.004 | 0.008 | 0.6 | 0.045 | 1551 | 385 |
| Conduct | Teacher | 0.3 | 0.935 | 0.002 | 0.034 | 0.002 | 0.908 | 0.000 | 0.9 | 0.027 | 1284 | 320 |
| Conduct | Child | 0.3 | 1.000 | 0.001 | 0.012 | 0.001 | 0.948 | 0.006 | 0.8 | 0.038 | 1529 | 380 |
| Emotional problems | Parent | 0.3 | 1.004 | 0.002 | 0.006 | 0.000 | 0.986 | 0.011 | 1.0 | 0.046 | 1552 | 385 |
| Emotional problems | Teacher | 0.3 | 0.982 | 0.001 | 0.014 | 0.001 | 1.022 | 0.001 | 0.6 | 0.035 | 1282 | 320 |
| Emotional problems | Child | 0.3 | 1.003 | 0.001 | 0.008 | 0.000 | 0.989 | 0.032 | 0.6 | 0.063 | 1529 | 380 |
| Peer problems | Parent | 0.3 | 1.012 | 0.002 | 0.008 | 0.001 | 0.998 | 0.001 | 0.3 | 0.062 | 1552 | 384 |
| Peer problems | Teacher | 0.3 | 0.996 | 0.002 | 0.006 | 0.000 | 0.995 | 0.001 | 1.0 | 0.047 | 1288 | 319 |
| Peer problems | Child | 0.3 | 0.999 | 0.002 | 0.008 | 0.001 | 0.959 | 0.006 | 0.4 | 0.079 | 1525 | 380 |
| Year 12 |  |  |  |  |  |  |  |  |  |  |  |  |
| Conduct | Teacher | 0.3 | 0.982 | 0.001 | 0.011 | 0.002 | 0.997 | 0.013 | 0.9 | 0.034 | 2272 | 565 |
| Conduct | Child | 0.3 | 0.991 | 0.001 | 0.012 | 0.001 | 0.974 | 0.018 | 0.1 | 0.074 | 2752 | 688 |
| Emotional problems | Parent | 0.3 | 0.995 | 0.000 | 0.009 | 0.000 | 1.030 | 0.000 | 0.3 | 0.078 | 2760 | 687 |
| Emotional problems | Teacher | 0.3 | 0.983 | 0.001 | 0.015 | 0.002 | 0.977 | 0.006 | 0.1 | 0.067 | 2265 | 565 |
| Emotional problems | Child | 0.3 | 0.999 | 0.001 | 0.015 | 0.001 | 0.983 | 0.004 | 1.0 | 0.019 | 2752 | 686 |
| Peer problems | Parent | 0.3 | 0.997 | 0.001 | 0.004 | 0.000 | 0.973 | 0.007 | 0.5 | 0.040 | 2759 | 688 |
| Peer problems | Teacher | 0.3 | 0.995 | 0.001 | 0.005 | 0.000 | 1.018 | 0.001 | 0.6 | 0.051 | 2271 | 566 |
| Peer problems | Child | 0.3 | 1.007 | 0.001 | 0.004 | 0.000 | 1.019 | 0.003 | 0.3 | 0.046 | 2753 | 687 |
| Year 16 |  |  |  |  |  |  |  |  |  |  |  |  |
| Hyperactivity | Child | 0.3 | 0.997 | 0.001 | 0.010 | 0.000 | 0.968 | 0.011 | 0.8 | 0.031 | 2388 | 596 |
| Hyperactivity | Parent | 0.3 | 0.993 | 0.001 | 0.009 | 0.000 | 0.992 | 0.004 | 0.2 | 0.067 | 2395 | 597 |
| Conduct | Child | 0.3 | 0.996 | 0.001 | 0.022 | 0.001 | 0.987 | 0.014 | 0.1 | 0.103 | 2389 | 595 |
| Conduct | Parent | 0.3 | 0.985 | 0.001 | 0.015 | 0.001 | 1.004 | 0.015 | 0.1 | 0.068 | 2400 | 598 |
| Emotional problems | Child | 0.3 | 1.007 | 0.001 | 0.007 | 0.001 | 1.001 | 0.008 | 0.8 | 0.029 | 2388 | 596 |
| Peer problems | Child | 0.3 | 0.999 | 0.001 | 0.008 | 0.001 | 1.029 | 0.000 | 0.3 | 0.052 | 2388 | 596 |
| Year 21 |  |  |  |  |  |  |  |  |  |  |  |  |
| Hyperactivity | Parent | 0.3 | 1.010 | 0.001 | 0.012 | 0.001 | 0.977 | 0.004 | 0.6 | 0.035 | 2496 | 622 |
| Hyperactivity | Child | 0.3 | 0.994 | 0.001 | 0.019 | 0.001 | 0.994 | 0.014 | 0.8 | 0.016 | 2240 | 557 |
| Conduct | Parent | 0.3 | 0.990 | 0.001 | 0.010 | 0.001 | 0.981 | 0.026 | 0.8 | 0.029 | 2496 | 623 |
| Conduct | Child | 0.3 | 0.978 | 0.001 | 0.016 | 0.001 | 0.998 | 0.002 | 0.1 | 0.079 | 2240 | 558 |
| Emotional problems | Parent | 0.3 | 0.989 | 0.001 | 0.016 | 0.001 | 1.005 | 0.020 | 1.0 | 0.019 | 2496 | 621 |
| Emotional problems | Child | 0.3 | 0.989 | 0.001 | 0.012 | 0.001 | 1.014 | 0.016 | 0.3 | 0.075 | 2240 | 557 |
| Peer problems | Parent | 0.3 | 0.985 | 0.001 | 0.010 | 0.001 | 0.972 | 0.012 | 0.2 | 0.067 | 2495 | 622 |
| Peer problems | Child | 0.3 | 0.985 | 0.001 | 0.023 | 0.002 | 0.964 | 0.006 | 0.1 | 0.098 | 2240 | 557 |
| Year 2 |  |  |  |  |  |  |  |  |  |  |  |  |
| Hyperactivity | Parent | 0.01 | 1.003 | 0.001 | 0.006 | 0.000 | 1.010 | 0.000 | 0.5 | 0.057 | 2182 | 544 |
| Conduct | Parent | 0.01 | 0.987 | 0.001 | 0.007 | 0.001 | 0.955 | 0.001 | 0.3 | 0.066 | 2178 | 543 |
| Emotional problems | Parent | 0.01 | 0.987 | 0.001 | 0.005 | 0.001 | 0.982 | 0.003 | 0.5 | 0.055 | 2186 | 544 |
| Year 3 |  |  |  |  |  |  |  |  |  |  |  |  |
| Hyperactivity | Parent | 0.01 | 1.007 | 0.001 | 0.006 | 0.000 | 1.004 | 0.002 | 1.0 | 0.024 | 2237 | 557 |
| Conduct | Parent | 0.01 | 0.988 | 0.001 | 0.010 | 0.001 | 0.986 | 0.003 | 0.2 | 0.078 | 2234 | 556 |
| Emotional problems | Parent | 0.01 | 0.992 | 0.001 | 0.004 | 0.000 | 1.009 | 0.008 | 1.0 | 0.030 | 2236 | 557 |
| Year 4 |  |  |  |  |  |  |  |  |  |  |  |  |
| Hyperactivity | Parent | 0.01 | 0.989 | 0.001 | 0.005 | 0.001 | 0.999 | 0.007 | 1.0 | 0.027 | 3176 | 792 |
| Conduct | Parent | 0.01 | 0.971 | 0.001 | 0.005 | 0.001 | 0.970 | 0.007 | 0.7 | 0.023 | 3176 | 791 |
| Emotional problems | Parent | 0.01 | 1.006 | 0.001 | 0.003 | 0.000 | 0.991 | 0.001 | 0.8 | 0.037 | 3176 | 791 |
| Peer problems | Parent | 0.01 | 1.001 | 0.000 | 0.004 | 0.001 | 0.996 | 0.000 | 1.0 | 0.037 | 3171 | 791 |
| Year 7 |  |  |  |  |  |  |  |  |  |  |  |  |
| Hyperactivity | Parent | 0.01 | 0.987 | 0.001 | 0.009 | 0.002 | 0.954 | 0.014 | 0.9 | 0.016 | 3251 | 812 |
| Hyperactivity | Teacher | 0.01 | 0.977 | 0.001 | 0.019 | 0.001 | 1.005 | 0.011 | 0.8 | 0.021 | 2695 | 672 |
| Conduct | Parent | 0.01 | 0.986 | 0.001 | 0.013 | 0.001 | 0.993 | 0.004 | 0.8 | 0.014 | 3253 | 812 |
| Conduct | Teacher | 0.01 | 1.279 | 0.001 | 0.012 | 0.001 | 1.272 | 0.010 | 1.0 | 0.019 | 2699 | 673 |
| Peer problems | Parent | 0.01 | 1.003 | 0.001 | 0.003 | 0.000 | 0.995 | 0.000 | 1.0 | 0.027 | 3253 | 811 |
| Peer problems | Teacher | 0.01 | 0.990 | 0.000 | 0.007 | 0.001 | 1.016 | 0.000 | 0.2 | 0.054 | 2690 | 672 |
| Emotional problems | Parent | 0.01 | 0.980 | 0.001 | 0.003 | 0.000 | 1.017 | 0.000 | 1.0 | 0.030 | 3252 | 812 |
| Emotional problems | Teacher | 0.01 | 0.995 | 0.001 | 0.004 | 0.000 | 0.995 | 0.011 | 0.6 | 0.043 | 2684 | 669 |
| Year 9 |  |  |  |  |  |  |  |  |  |  |  |  |
| Hyperactivity | Parent | 0.01 | 0.997 | 0.001 | 0.018 | 0.002 | 0.983 | 0.000 | 0.6 | 0.048 | 1551 | 384 |
| Hyperactivity | Teacher | 0.01 | 0.997 | 0.002 | 0.023 | 0.003 | 0.982 | 0.037 | 0.7 | 0.056 | 1283 | 320 |
| Hyperactivity | Child | 0.01 | 1.008 | 0.002 | 0.007 | 0.001 | 0.980 | 0.002 | 0.4 | 0.062 | 1532 | 380 |
| Conduct | Parent | 0.01 | 1.004 | 0.001 | 0.017 | 0.001 | 1.003 | 0.008 | 0.2 | 0.093 | 1551 | 385 |
| Conduct | Teacher | 0.01 | 0.938 | 0.001 | 0.029 | 0.001 | 0.899 | 0.005 | 0.6 | 0.052 | 1284 | 320 |
| Conduct | Child | 0.01 | 1.002 | 0.002 | 0.009 | 0.002 | 0.947 | 0.008 | 0.5 | 0.080 | 1529 | 380 |
| Peer problems | Parent | 0.01 | 1.011 | 0.001 | 0.011 | 0.001 | 1.005 | 0.000 | 0.2 | 0.061 | 1552 | 384 |
| Peer problems | Teacher | 0.01 | 0.996 | 0.002 | 0.006 | 0.000 | 0.996 | 0.003 | 1.0 | 0.045 | 1288 | 319 |
| Peer problems | Child | 0.01 | 1.002 | 0.002 | 0.006 | 0.000 | 0.961 | 0.012 | 1.0 | 0.047 | 1525 | 380 |
| Emotional problems | Parent | 0.01 | 1.005 | 0.002 | 0.007 | 0.001 | 0.987 | 0.001 | 1.0 | 0.039 | 1552 | 385 |
| Emotional problems | Teacher | 0.01 | 0.986 | 0.002 | 0.008 | 0.001 | 1.016 | 0.000 | 0.5 | 0.053 | 1282 | 320 |
| Emotional problems | Child | 0.01 | 1.007 | 0.002 | 0.007 | 0.001 | 0.995 | 0.000 | 1.0 | 0.040 | 1529 | 380 |
| Year 12 |  |  |  |  |  |  |  |  |  |  |  |  |
| Conduct | Teacher | 0.01 | 0.988 | 0.002 | 0.004 | 0.001 | 0.999 | 0.013 | 0.5 | 0.054 | 2272 | 565 |
| Conduct | Child | 0.01 | 0.994 | 0.001 | 0.005 | 0.001 | 0.978 | 0.014 | 0.9 | 0.027 | 2752 | 688 |
| Emotional problems | Parent | 0.01 | 0.998 | 0.001 | 0.006 | 0.000 | 1.030 | 0.001 | 0.5 | 0.053 | 2760 | 687 |
| Emotional problems | Teacher | 0.01 | 0.987 | 0.001 | 0.008 | 0.000 | 0.976 | 0.006 | 0.3 | 0.065 | 2265 | 565 |
| Emotional problems | Child | 0.01 | 1.004 | 0.000 | 0.006 | 0.000 | 0.984 | 0.001 | 0.4 | 0.059 | 2752 | 686 |
| Peer problems | Parent | 0.01 | 0.997 | 0.001 | 0.004 | 0.000 | 0.974 | 0.003 | 0.6 | 0.041 | 2759 | 688 |
| Peer problems | Teacher | 0.01 | 0.995 | 0.001 | 0.005 | 0.000 | 1.019 | 0.000 | 0.3 | 0.054 | 2271 | 566 |
| Peer problems | Child | 0.01 | 1.007 | 0.000 | 0.005 | 0.001 | 1.021 | 0.006 | 1.0 | 0.041 | 2753 | 687 |
| Year 16 |  |  |  |  |  |  |  |  |  |  |  |  |
| Hyperactivity | Child | 0.01 | 1.000 | 0.001 | 0.005 | 0.000 | 0.969 | 0.008 | 0.3 | 0.045 | 2388 | 596 |
| Hyperactivity | Parent | 0.01 | 0.994 | 0.001 | 0.007 | 0.000 | 0.995 | 0.001 | 0.2 | 0.055 | 2395 | 597 |
| Conduct | Child | 0.01 | 1.004 | 0.001 | 0.007 | 0.001 | 0.993 | 0.001 | 1.0 | 0.020 | 2389 | 595 |
| Conduct | Parent | 0.01 | 0.988 | 0.000 | 0.008 | 0.001 | 1.009 | 0.006 | 0.1 | 0.057 | 2400 | 598 |
| Emotional problems | Child | 0.01 | 1.008 | 0.001 | 0.005 | 0.001 | 1.001 | 0.010 | 0.4 | 0.060 | 2388 | 596 |
| Peer problems | Child | 0.01 | 0.999 | 0.000 | 0.007 | 0.001 | 1.032 | 0.001 | 0.3 | 0.048 | 2388 | 596 |
| Year 21 |  |  |  |  |  |  |  |  |  |  |  |  |
| Hyperactivity | Parent | 0.01 | 1.014 | 0.001 | 0.005 | 0.002 | 0.976 | 0.006 | 1.0 | 0.030 | 2496 | 622 |
| Hyperactivity | Child | 0.01 | 0.998 | 0.001 | 0.012 | 0.002 | 0.996 | 0.011 | 0.6 | 0.033 | 2240 | 557 |
| Conduct | Parent | 0.01 | 0.992 | 0.001 | 0.007 | 0.000 | 0.985 | 0.017 | 1.0 | 0.025 | 2496 | 623 |
| Conduct | Child | 0.01 | 0.983 | 0.001 | 0.007 | 0.001 | 0.995 | 0.003 | 0.2 | 0.054 | 2240 | 558 |
| Emotional problems | Parent | 0.01 | 0.991 | 0.001 | 0.012 | 0.001 | 1.011 | 0.009 | 0.6 | 0.031 | 2496 | 621 |
| Emotional problems | Child | 0.01 | 0.991 | 0.001 | 0.007 | 0.000 | 1.017 | 0.010 | 1.0 | 0.026 | 2240 | 557 |
| Peer problems | Parent | 0.01 | 0.987 | 0.000 | 0.007 | 0.000 | 0.976 | 0.004 | 0.4 | 0.052 | 2495 | 622 |
| Peer problems | Child | 0.01 | 0.990 | 0.001 | 0.014 | 0.001 | 0.963 | 0.005 | 0.6 | 0.033 | 2240 | 557 |

| **Supporting table S6**. Behaviour problems composites: univariate twin model fitting results. | | | | | | | | | |
| --- | --- | --- | --- | --- | --- | --- | --- | --- | --- |
| Note. | | | | | | | | | |
| Est= estimate; A= genetic influences; C= shared environmental influences; E= unique environmental influences; CI= confidence interval. | | | | | | | | | |
| * cross-age approach: combined individual behaviour problems at all ages (2-21) for parent, teacher and child ratings to create the first order factors of cross-age externalizing and internalizing, which were then combined across raters to create the second-order factors of cross-age-and-rater externalizing and internalizing. | | | | | | | | | |
| ** cross-rater approach: combined individual behaviour problems in adolescence and adulthood to create the first order factors of cross-rater externalizing and internalizing, which were then combined across developmental stages to create the second-order factors of cross-age-and-rater externalizing and internalizing. | | | | | | | | | |
| **Composites** | **Univariate twin model fitting results** | | | | | | | | |
|  | **A** | | | **C** | | | **E** | | |
|  | Est | Lower  95% CI | Upper 95% CI | Est | Lower  95% CI | Upper  95% CI | Est | Lower  95% CI | Upper  95% CI |
| **Cross-age** |  |  |  |  |  |  |  |  |  |
| BPp parent-rated | 0.592 | 0.545 | 0.641 | 0.265 | 0.218 | 0.310 | 0.143 | 0.133 | 0.154 |
| BPp teacher-rated | 0.721 | 0.662 | 0.740 | 0.000 | 0.000 | 0.051 | 0.279 | 0.260 | 0.300 |
| BPp child-rated | 0.483 | 0.398 | 0.567 | 0.077 | 0.007 | 0.146 | 0.440 | 0.411 | 0.471 |
| Externalizing parent-rated | 0.825 | 0.807 | 0.837 | 0.000 | 0.000 | 0.014 | 0.175 | 0.163 | 0.189 |
| Externalizing teacher-rated | 0.736 | 0.708 | 0.754 | 0.000 | 0.000 | 0.021 | 0.264 | 0.246 | 0.284 |
| Externalizing child-rated | 0.531 | 0.442 | 0.565 | 0.005 | 0.000 | 0.077 | 0.464 | 0.435 | 0.496 |
| Internalizing parent-rated | 0.480 | 0.430 | 0.530 | 0.325 | 0.278 | 0.370 | 0.195 | 0.182 | 0.210 |
| Internalizing teacher-rated | 0.653 | 0.580 | 0.701 | 0.027 | 0.000 | 0.089 | 0.320 | 0.298 | 0.345 |
| Internalizing child-rated | 0.463 | 0.378 | 0.547 | 0.093 | 0.048 | 0.161 | 0.445 | 0.416 | 0.476 |
| **Cross-rater** |  |  |  |  |  |  |  |  |  |
| BPp childhood | 0.654 | 0.601 | 0.708 | 0.173 | 0.121 | 0.223 | 0.173 | 0.161 | 0.186 |
| BPp adolescence | 0.543 | 0.474 | 0.612 | 0.145 | 0.130 | 0.204 | 0.312 | 0.290 | 0.335 |
| BPp adulthood | 0.515 | 0.431 | 0.599 | 0.106 | 0.034 | 0.175 | 0.379 | 0.352 | 0.409 |
| Externalizing childhood | 0.790 | 0.773 | 0.805 | 0.000 | 0.000 | 0.008 | 0.210 | 0.195 | 0.227 |
| Externalizing adolescence | 0.672 | 0.599 | 0.707 | 0.013 | 0.000 | 0.076 | 0.315 | 0.293 | 0.338 |
| Externalizing adulthood | 0.565 | 0.480 | 0.643 | 0.054 | 0.000 | 0.125 | 0.381 | 0.353 | 0.411 |
| Internalizing childhood | 0.514 | 0.459 | 0.570 | 0.259 | 0.208 | 0.309 | 0.227 | 0.211 | 0.244 |
| Internalizing adolescence | 0.511 | 0.438 | 0.584 | 0.142 | 0.079 | 0.203 | 0.347 | 0.324 | 0.373 |
| Internalizing adulthood | 0.502 | 0.414 | 0.589 | 0.092 | 0.018 | 0.164 | 0.406 | 0.377 | 0.438 |
| **Cross-trait-and-rater (cross-age approach) *** |  |  |  |  |  |  |  |  |  |
| BPp | 0.622 | 0.675 | 0.729 | 0.108 | 0.160 | 0.210 | 0.154 | 0.165 | 0.178 |
| Externalizing | 0.805 | 0.831 | 0.842 | 0.000 | 0.000 | 0.023 | 0.158 | 0.169 | 0.182 |
| Internalizing | 0.505 | 0.558 | 0.613 | 0.189 | 0.240 | 0.289 | 0.188 | 0.202 | 0.217 |
| **Cross-trait-and-rater (cross-rater approach) **** |  |  |  |  |  |  |  |  |  |
| BPp | 0.574 | 0.628 | 0.684 | 0.131 | 0.184 | 0.234 | 0.175 | 0.188 | 0.202 |
| Externalizing | 0.789 | 0.803 | 0.817 | 0.000 | 0.000 | 0.051 | 0.183 | 0.197 | 0.211 |
| Internalizing | 0.487 | 0.543 | 0.599 | 0.183 | 0.236 | 0.286 | 0.206 | 0.222 | 0.238 |
| **Single-trait cross-age** |  |  |  |  |  |  |  |  |  |
| Hyperactivity parent-rated | 0.646 | 0.613 | 0.676 | 0.000 | 0.000 | 0.003 | 0.354 | 0.324 | 0.387 |
| Hyperactivity teacher-rated | 0.712 | 0.688 | 0.734 | 0.000 | 0.000 | 0.010 | 0.288 | 0.266 | 0.312 |
| Hyperactivity child-rated | 0.429 | 0.391 | 0.465 | 0.000 | 0.000 | 0.017 | 0.571 | 0.535 | 0.609 |
| Conduct parent-rated | 0.617 | 0.565 | 0.670 | 0.206 | 0.156 | 0.255 | 0.177 | 0.165 | 0.190 |
| Conduct teacher-rated | 0.654 | 0.609 | 0.677 | 0.000 | 0.000 | 0.036 | 0.346 | 0.323 | 0.370 |
| Conduct child-rated | 0.425 | 0.357 | 0.458 | 0.000 | 0.000 | 0.051 | 0.575 | 0.542 | 0.609 |
| Emotional problems parent-rated | 0.554 | 0.492 | 0.618 | 0.173 | 0.116 | 0.228 | 0.272 | 0.254 | 0.292 |
| Emotional problems teacher-rated | 0.550 | 0.520 | 0.578 | 0.000 | 0.000 | 0.022 | 0.450 | 0.422 | 0.480 |
| Emotional problems child-rated | 0.462 | 0.373 | 0.492 | 0.000 | 0.000 | 0.069 | 0.538 | 0.508 | 0.570 |
| Peer problems parent-rated | 0.694 | 0.636 | 0.754 | 0.094 | 0.039 | 0.147 | 0.212 | 0.197 | 0.228 |
| Peer problems teacher-rated | 0.592 | 0.514 | 0.656 | 0.040 | 0.000 | 0.104 | 0.368 | 0.342 | 0.395 |
| Peer problems child-rated | 0.466 | 0.376 | 0.540 | 0.043 | 0.000 | 0.114 | 0.490 | 0.459 | 0.524 |
| **Single-trait cross-rater** |  |  |  |  |  |  |  |  |  |
| Hyperactivity childhood | 0.614 | 0.578 | 0.647 | 0.000 | 0.000 | 0.003 | 0.386 | 0.353 | 0.422 |
| Hyperactivity adolescence | 0.736 | 0.712 | 0.757 | 0.000 | 0.000 | 0.010 | 0.264 | 0.243 | 0.288 |
| Hyperactivity adulthood | 0.600 | 0.569 | 0.629 | 0.000 | 0.000 | 0.014 | 0.400 | 0.371 | 0.431 |
| Conduct childhood | 0.631 | 0.576 | 0.687 | 0.174 | 0.122 | 0.225 | 0.195 | 0.181 | 0.210 |
| Conduct adolescence | 0.578 | 0.496 | 0.625 | 0.021 | 0.000 | 0.088 | 0.401 | 0.375 | 0.430 |
| Conduct adulthood | 0.502 | 0.421 | 0.582 | 0.131 | 0.063 | 0.197 | 0.367 | 0.341 | 0.396 |
| Emotional problems childhood | 0.598 | 0.531 | 0.667 | 0.100 | 0.040 | 0.158 | 0.302 | 0.281 | 0.324 |
| Emotional problems adolescence | 0.495 | 0.403 | 0.538 | 0.012 | 0.000 | 0.084 | 0.493 | 0.462 | 0.527 |
| Emotional problems adulthood | 0.555 | 0.523 | 0.585 | 0.000 | 0.000 | 0.031 | 0.445 | 0.415 | 0.477 |
| Peer problems childhood | 0.704 | 0.640 | 0.759 | 0.040 | 0.000 | 0.097 | 0.256 | 0.238 | 0.275 |
| Peer problems adolescence | 0.561 | 0.487 | 0.635 | 0.093 | 0.029 | 0.154 | 0.346 | 0.323 | 0.372 |
| Peer problems adulthood | 0.702 | 0.648 | 0.724 | 0.000 | 0.000 | 0.047 | 0.298 | 0.276 | 0.320 |

| **Supporting table S7**. Behaviour problems composites: univariate twin model fitting results, for males and females separately. | | | | | | | | | |
| --- | --- | --- | --- | --- | --- | --- | --- | --- | --- |
| Note. | | | | | | | | | |
| Est= estimate; A= genetic influences; C= shared environmental influences; E= unique environmental influences; CI= confidence interval. | | | | | | | | | |
| * cross-age approach: combined individual behaviour problems at all ages (2-21) for parent, teacher and child ratings to create the first order factors of cross-age externalizing and internalizing, which were then combined across raters to create the second-order factors of cross-age-and-rater externalizing and internalizing. | | | | | | | | | |
| ** cross-rater approach: combined individual behaviour problems in adolescence and adulthood to create the first order factors of cross-rater externalizing and internalizing, which were then combined across developmental stages to create the second-order factors of cross-age-and-rater externalizing and internalizing. | | | | | | | | | |
| **Composites** | **Univariate twin model fitting results: males only** | | | | | | | | |
|  | **A** | | | **C** | | | **E** | | |
|  | Est | Lower  95% CI | Upper  95% CI | Est | Lower  95% CI | Upper  95% CI | Est | Lower  95% CI | Upper  95% CI |
| **Cross-age** |  |  |  |  |  |  |  |  |  |
| BPp parent-rated | 0.574 | 0.505 | 0.647 | 0.277 | 0.207 | 0.342 | 0.149 | 0.134 | 0.167 |
| BPp teacher-rated | 0.714 | 0.615 | 0.765 | 0.024 | 0.000 | 0.112 | 0.262 | 0.235 | 0.293 |
| BPp child-rated | 0.508 | 0.377 | 0.596 | 0.043 | 0.000 | 0.144 | 0.449 | 0.404 | 0.499 |
| Externalizing parent-rated | 0.828 | 0.793 | 0.846 | 0.000 | 0.000 | 0.031 | 0.172 | 0.154 | 0.193 |
| Externalizing teacher-rated | 0.748 | 0.672 | 0.774 | 0.000 | 0.000 | 0.067 | 0.252 | 0.226 | 0.281 |
| Externalizing child-rated | 0.521 | 0.404 | 0.565 | 0.000 | 0.000 | 0.088 | 0.479 | 0.435 | 0.526 |
| Internalizing parent-rated | 0.446 | 0.372 | 0.522 | 0.347 | 0.277 | 0.412 | 0.207 | 0.186 | 0.232 |
| Internalizing teacher-rated | 0.671 | 0.566 | 0.733 | 0.032 | 0.000 | 0.122 | 0.297 | 0.266 | 0.331 |
| Internalizing child-rated | 0.497 | 0.366 | 0.597 | 0.056 | 0.000 | 0.157 | 0.447 | 0.402 | 0.497 |
| **Cross-rater** |  |  |  |  |  |  |  |  |  |
| BPp childhood | 0.591 | 0.514 | 0.671 | 0.227 | 0.151 | 0.298 | 0.182 | 0.163 | 0.204 |
| BPp adolescence | 0.559 | 0.458 | 0.661 | 0.141 | 0.054 | 0.226 | 0.300 | 0.269 | 0.335 |
| BPp adulthood | 0.529 | 0.392 | 0.642 | 0.069 | 0.000 | 0.176 | 0.401 | 0.355 | 0.455 |
| Externalizing childhood | 0.788 | 0.762 | 0.810 | 0.000 | 0.000 | 0.023 | 0.212 | 0.190 | 0.238 |
| Externalizing adolescence | 0.658 | 0.553 | 0.731 | 0.043 | 0.000 | 0.132 | 0.299 | 0.268 | 0.334 |
| Externalizing adulthood | 0.563 | 0.423 | 0.637 | 0.027 | 0.000 | 0.136 | 0.410 | 0.363 | 0.464 |
| Internalizing childhood | 0.447 | 0.365 | 0.530 | 0.313 | 0.239 | 0.383 | 0.240 | 0.215 | 0.268 |
| Internalizing adolescence | 0.525 | 0.415 | 0.634 | 0.130 | 0.037 | 0.220 | 0.345 | 0.310 | 0.385 |
| Internalizing adulthood | 0.526 | 0.386 | 0.633 | 0.061 | 0.000 | 0.169 | 0.413 | 0.366 | 0.468 |
| **Composites** | **Univariate twin model fitting results: females only** | | | | | | | | |
|  | **A** | | | **C** | | | **E** | | |
|  | Est | Lower 95% CI | Upper 95% CI | Esti | Lower 95% CI | Upper 95% CI | Est | Lower 95% CI | Upper 95% CI |
| **Cross-age** |  |  |  |  |  |  |  |  |  |
| BPp parent-rated | 0.607 | 0.544 | 0.673 | 0.255 | 0.190 | 0.316 | 0.138 | 0.126 | 0.152 |
| BPp teacher-rated | 0.693 | 0.631 | 0.721 | 0.000 | 0.000 | 0.052 | 0.307 | 0.279 | 0.338 |
| BPp child-rated | 0.464 | 0.352 | 0.577 | 0.103 | 0.008 | 0.195 | 0.433 | 0.396 | 0.472 |
| Externalizing parent-rated | 0.821 | 0.795 | 0.838 | 0.000 | 0.000 | 0.022 | 0.179 | 0.162 | 0.198 |
| Externalizing teacher-rated | 0.711 | 0.681 | 0.739 | 0.000 | 0.000 | 0.025 | 0.289 | 0.261 | 0.319 |
| Externalizing child-rated | 0.521 | 0.404 | 0.583 | 0.024 | 0.000 | 0.119 | 0.455 | 0.417 | 0.497 |
| Internalizing parent-rated | 0.506 | 0.439 | 0.575 | 0.307 | 0.242 | 0.369 | 0.187 | 0.170 | 0.206 |
| Internalizing teacher-rated | 0.628 | 0.525 | 0.684 | 0.024 | 0.000 | 0.111 | 0.348 | 0.316 | 0.383 |
| Internalizing child-rated | 0.440 | 0.327 | 0.553 | 0.119 | 0.023 | 0.211 | 0.441 | 0.405 | 0.481 |
| **Cross-rater** |  |  |  |  |  |  |  |  |  |
| BPp childhood | 0.708 | 0.635 | 0.784 | 0.125 | 0.084 | 0.194 | 0.167 | 0.152 | 0.184 |
| BPp adolescence | 0.533 | 0.439 | 0.628 | 0.147 | 0.062 | 0.227 | 0.321 | 0.292 | 0.352 |
| BPp adulthood | 0.498 | 0.390 | 0.606 | 0.137 | 0.042 | 0.227 | 0.366 | 0.332 | 0.402 |
| Externalizing childhood | 0.789 | 0.766 | 0.810 | 0.000 | 0.000 | 0.012 | 0.211 | 0.190 | 0.234 |
| Externalizing adolescence | 0.672 | 0.589 | 0.701 | 0.000 | 0.000 | 0.072 | 0.328 | 0.299 | 0.359 |
| Externalizing adulthood | 0.556 | 0.447 | 0.661 | 0.080 | 0.000 | 0.172 | 0.364 | 0.331 | 0.401 |
| Internalizing childhood | 0.569 | 0.494 | 0.647 | 0.213 | 0.140 | 0.281 | 0.218 | 0.198 | 0.240 |
| Internalizing adolescence | 0.500 | 0.402 | 0.599 | 0.152 | 0.065 | 0.234 | 0.349 | 0.318 | 0.383 |
| Internalizing adulthood | 0.486 | 0.372 | 0.600 | 0.114 | 0.015 | 0.209 | 0.400 | 0.365 | 0.439 |

| **Supporting table S8**. SDQ scales (observed traits): univariate twin model fitting results. | | | | | | | | | | |
| --- | --- | --- | --- | --- | --- | --- | --- | --- | --- | --- |
| Note. | | | | | | | | | | |
| Est= estimate; A= genetic influences; C= shared environmental influences; E= unique environmental influences; CI= confidence interval. | | | | | | | | | | |
| **Trait** | **Rater** | **Univariate twin model fitting results** | | | | | | | | |
|  |  | **A** | | | **C** | | | **E** | | |
|  |  | Est | Lower  95% CI | Upper  95% CI | Est | Lower  95% CI | Upper  95% CI | Est | Lower  95% CI | Upper  95% CI |
| Year 2 |  |  |  |  |  |  |  |  |  |  |
| Hyperactivity | Parent | 0.638 | 0.611 | 0.663 | 0.000 | 0.000 | 0.005 | 0.362 | 0.337 | 0.389 |
| Conduct | Parent | 0.510 | 0.452 | 0.568 | 0.198 | 0.147 | 0.247 | 0.292 | 0.274 | 0.312 |
| Emotional problems | Parent | 0.557 | 0.529 | 0.584 | 0.000 | 0.000 | 0.013 | 0.443 | 0.416 | 0.471 |
| Year 3 |  |  |  |  |  |  |  |  |  |  |
| Hyperactivity | Parent | 0.557 | 0.523 | 0.589 | 0.000 | 0.000 | 0.003 | 0.443 | 0.411 | 0.477 |
| Conduct | Parent | 0.505 | 0.448 | 0.562 | 0.218 | 0.168 | 0.266 | 0.278 | 0.260 | 0.297 |
| Emotional problems | Parent | 0.525 | 0.495 | 0.553 | 0.000 | 0.000 | 0.014 | 0.475 | 0.447 | 0.504 |
| Year 4 |  |  |  |  |  |  |  |  |  |  |
| Hyperactivity | Parent | 0.349 | 0.314 | 0.383 | 0.000 | 0.000 | 0.003 | 0.651 | 0.617 | 0.686 |
| Conduct | Parent | 0.622 | 0.561 | 0.662 | 0.020 | 0.000 | 0.070 | 0.358 | 0.338 | 0.379 |
| Emotional problems | Parent | 0.539 | 0.472 | 0.591 | 0.030 | 0.000 | 0.083 | 0.432 | 0.408 | 0.457 |
| Peer problems | Parent | 0.640 | 0.583 | 0.696 | 0.045 | 0.000 | 0.093 | 0.315 | 0.298 | 0.334 |
| Year 7 |  |  |  |  |  |  |  |  |  |  |
| Hyperactivity | Parent | 0.468 | 0.437 | 0.498 | 0.000 | 0.000 | 0.000 | 0.532 | 0.502 | 0.563 |
| Hyperactivity | Teacher | 0.527 | 0.495 | 0.553 | 0.000 | 0.000 | 0.020 | 0.473 | 0.447 | 0.500 |
| Conduct | Parent | 0.586 | 0.539 | 0.634 | 0.175 | 0.131 | 0.217 | 0.239 | 0.226 | 0.254 |
| Conduct | Teacher | 0.712 | 0.666 | 0.729 | 0.000 | 0.000 | 0.039 | 0.288 | 0.271 | 0.307 |
| Emotional problems | Parent | 0.465 | 0.404 | 0.525 | 0.149 | 0.099 | 0.199 | 0.386 | 0.365 | 0.408 |
| Emotional problems | Teacher | 0.713 | 0.693 | 0.732 | 0.000 | 0.000 | 0.008 | 0.287 | 0.268 | 0.307 |
| Peer problems | Parent | 0.660 | 0.601 | 0.689 | 0.011 | 0.000 | 0.060 | 0.330 | 0.311 | 0.349 |
| Peer problems | Teacher | 0.669 | 0.602 | 0.690 | 0.001 | 0.000 | 0.058 | 0.330 | 0.310 | 0.352 |
| Year 9 |  |  |  |  |  |  |  |  |  |  |
| Hyperactivity | Parent | 0.701 | 0.670 | 0.729 | 0.000 | 0.000 | 0.006 | 0.299 | 0.271 | 0.330 |
| Hyperactivity | Teacher | 0.609 | 0.569 | 0.644 | 0.000 | 0.000 | 0.023 | 0.391 | 0.356 | 0.429 |
| Hyperactivity | Child | 0.388 | 0.343 | 0.430 | 0.000 | 0.000 | 0.021 | 0.612 | 0.570 | 0.655 |
| Conduct | Parent | 0.536 | 0.472 | 0.601 | 0.257 | 0.195 | 0.315 | 0.207 | 0.190 | 0.226 |
| Conduct | Teacher | 0.571 | 0.504 | 0.607 | 0.000 | 0.000 | 0.049 | 0.429 | 0.393 | 0.469 |
| Conduct | Child | 0.442 | 0.329 | 0.517 | 0.034 | 0.000 | 0.121 | 0.523 | 0.483 | 0.567 |
| Emotional problems | Parent | 0.449 | 0.363 | 0.535 | 0.193 | 0.120 | 0.264 | 0.358 | 0.330 | 0.388 |
| Emotional problems | Teacher | 0.496 | 0.434 | 0.537 | 0.000 | 0.000 | 0.044 | 0.504 | 0.463 | 0.547 |
| Emotional problems | Child | 0.392 | 0.275 | 0.477 | 0.044 | 0.000 | 0.134 | 0.564 | 0.522 | 0.608 |
| Peer problems | Parent | 0.587 | 0.513 | 0.662 | 0.151 | 0.083 | 0.216 | 0.262 | 0.241 | 0.286 |
| Peer problems | Teacher | 0.511 | 0.397 | 0.598 | 0.049 | 0.000 | 0.139 | 0.440 | 0.402 | 0.483 |
| Peer problems | Child | 0.319 | 0.197 | 0.435 | 0.077 | 0.000 | 0.168 | 0.604 | 0.559 | 0.652 |
| Year 12 |  |  |  |  |  |  |  |  |  |  |
| Conduct | Teacher | 0.566 | 0.477 | 0.604 | 0.009 | 0.000 | 0.080 | 0.425 | 0.396 | 0.457 |
| Conduct | Child | 0.376 | 0.288 | 0.462 | 0.065 | 0.000 | 0.132 | 0.560 | 0.528 | 0.593 |
| Emotional problems | Parent | 0.512 | 0.443 | 0.582 | 0.109 | 0.050 | 0.167 | 0.378 | 0.355 | 0.403 |
| Emotional problems | Teacher | 0.448 | 0.405 | 0.482 | 0.000 | 0.000 | 0.027 | 0.552 | 0.518 | 0.587 |
| Emotional problems | Child | 0.350 | 0.260 | 0.439 | 0.065 | 0.000 | 0.133 | 0.585 | 0.552 | 0.620 |
| Peer problems | Parent | 0.686 | 0.629 | 0.743 | 0.076 | 0.023 | 0.127 | 0.238 | 0.223 | 0.254 |
| Peer problems | Teacher | 0.525 | 0.447 | 0.555 | 0.000 | 0.000 | 0.060 | 0.475 | 0.445 | 0.508 |
| Peer problems | Child | 0.356 | 0.263 | 0.417 | 0.026 | 0.000 | 0.097 | 0.617 | 0.583 | 0.654 |
| Year 16 |  |  |  |  |  |  |  |  |  |  |
| Hyperactivity | Parent | 0.763 | 0.744 | 0.780 | 0.000 | 0.000 | 0.010 | 0.237 | 0.220 | 0.256 |
| Hyperactivity | Child | 0.385 | 0.349 | 0.420 | 0.000 | 0.000 | 0.016 | 0.615 | 0.580 | 0.651 |
| Conduct | Parent | 0.630 | 0.569 | 0.692 | 0.118 | 0.062 | 0.172 | 0.252 | 0.235 | 0.270 |
| Conduct | Child | 0.350 | 0.307 | 0.385 | 0.000 | 0.000 | 0.025 | 0.650 | 0.615 | 0.685 |
| Emotional problems | Child | 0.388 | 0.341 | 0.421 | 0.000 | 0.000 | 0.000 | 0.612 | 0.579 | 0.646 |
| Peer problems | Child | 0.391 | 0.293 | 0.448 | 0.022 | 0.000 | 0.096 | 0.587 | 0.552 | 0.625 |
| Year 21 |  |  |  |  |  |  |  |  |  |  |
| Hyperactivity | Parent | 0.612 | 0.585 | 0.638 | 0.000 | 0.000 | 0.012 | 0.388 | 0.362 | 0.415 |
| Hyperactivity | Child | 0.342 | 0.245 | 0.381 | 0.000 | 0.000 | 0.000 | 0.658 | 0.619 | 0.699 |
| Conduct | Parent | 0.514 | 0.441 | 0.586 | 0.113 | 0.052 | 0.173 | 0.373 | 0.349 | 0.399 |
| Conduct | Child | 0.209 | 0.082 | 0.277 | 0.022 | 0.000 | 0.116 | 0.769 | 0.723 | 0.819 |
| Emotional problems | Parent | 0.566 | 0.523 | 0.592 | 0.000 | 0.000 | 0.000 | 0.434 | 0.408 | 0.461 |
| Emotional problems | Child | 0.309 | 0.192 | 0.381 | 0.030 | 0.000 | 0.118 | 0.661 | 0.619 | 0.706 |
| Peer problems | Parent | 0.722 | 0.659 | 0.754 | 0.015 | 0.000 | 0.072 | 0.263 | 0.246 | 0.282 |
| Peer problems | Child | 0.397 | 0.295 | 0.433 | 0.000 | 0.000 | 0.000 | 0.603 | 0.567 | 0.643 |

| **Supporting table S9**. Weights for the individual polygenic scores from elastic net regularization. | | | | | | | | | | | | | | | | |
| --- | --- | --- | --- | --- | --- | --- | --- | --- | --- | --- | --- | --- | --- | --- | --- | --- |
| Note. | | | | | | | | | | | | | | | | |
| ADHD= attention deficit hyperactivity disorder; AN= anorexia Nervosa; ASD= autism spectrum disorder; BPD= bipolar disorder; DS= depressive symptoms; INS= insomnia; IRR= irritability; MDD= major depressive disorder; MS= mood swings; NEU= neuroticism; OCD= obsessive-compulsive disorder; PTSD= post-traumatic stress disorder; RT= risk taking; SCZ= schizophrenia; SWB= subjective wellbeing. | | | | | | | | | | | | | | | | |
| **Composites** | | **GPS weights for the multi-trait GPS** | | | | | | | | | | | | | | |
|  | ADHD | | AN | ASD | BPD | DS | INS | IRR | MDD | MS | NEU | OCD | PTSD | RT | SCZ | SWB |
| **Cross-age** |  | |  |  |  |  |  |  |  |  |  |  |  |  |  |  |
| BPp parent-rated | 0.069 | | 0.000 | 0.000 | 0.000 | 0.000 | 0.002 | 0.000 | 0.015 | 0.013 | 0.023 | 0.000 | 0.000 | 0.000 | 0.000 | 0.000 |
| BPp teacher-rated | 0.104 | | -0.010 | 0.000 | 0.000 | 0.015 | 0.000 | 0.000 | 0.000 | 0.000 | 0.000 | 0.003 | 0.000 | 0.037 | 0.000 | 0.004 |
| BPp child-rated | 0.067 | | -0.031 | -0.006 | -0.052 | 0.009 | 0.018 | -0.033 | 0.056 | 0.023 | 0.066 | 0.000 | 0.019 | 0.053 | 0.022 | -0.027 |
| Externalizing  parent-rated | 0.104 | | 0.000 | 0.000 | -0.016 | 0.000 | 0.015 | -0.015 | 0.011 | 0.049 | 0.000 | 0.000 | 0.000 | 0.014 | 0.000 | -0.018 |
| Externalizing  teacher-rated | 0.128 | | -0.008 | 0.000 | -0.006 | 0.021 | 0.000 | -0.005 | -0.001 | 0.000 | 0.000 | 0.021 | 0.000 | 0.050 | 0.000 | 0.028 |
| Externalizing  child-rated | 0.072 | | -0.028 | 0.000 | -0.027 | 0.000 | 0.022 | -0.034 | 0.018 | 0.068 | 0.015 | 0.000 | 0.003 | 0.079 | 0.014 | -0.048 |
| Internalizing  parent-rated | 0.036 | | 0.000 | 0.000 | -0.022 | 0.000 | 0.000 | -0.025 | 0.029 | 0.002 | 0.098 | 0.000 | 0.000 | 0.000 | 0.000 | 0.000 |
| Internalizing  teacher-rated | 0.057 | | 0.000 | 0.000 | 0.000 | 0.000 | 0.000 | 0.000 | 0.000 | 0.000 | 0.001 | 0.000 | 0.000 | 0.000 | 0.000 | 0.000 |
| Internalizing  child-rated | 0.051 | | -0.034 | 0.016 | -0.048 | 0.000 | 0.004 | -0.039 | 0.033 | 0.045 | 0.091 | 0.000 | 0.014 | 0.023 | 0.008 | -0.016 |
| **Cross-rater** |  | |  |  |  |  |  |  |  |  |  |  |  |  |  |  |
| BPp childhood | 0.094 | | -0.003 | 0.017 | -0.021 | 0.000 | 0.000 | -0.031 | 0.011 | 0.020 | 0.024 | 0.019 | 0.000 | 0.000 | 0.000 | -0.010 |
| BPp adolescence | 0.061 | | -0.018 | -0.003 | -0.009 | 0.000 | 0.022 | -0.015 | 0.032 | 0.038 | 0.059 | 0.000 | 0.011 | 0.052 | 0.000 | -0.008 |
| BPp adulthood | 0.060 | | 0.000 | 0.000 | -0.021 | 0.000 | 0.018 | 0.000 | 0.059 | 0.053 | 0.000 | 0.000 | 0.000 | 0.023 | 0.033 | 0.000 |
| Externalizing  childhood | 0.116 | | 0.000 | 0.000 | 0.000 | 0.000 | 0.000 | 0.000 | 0.000 | 0.014 | 0.005 | 0.000 | 0.000 | 0.000 | 0.000 | 0.000 |
| Externalizing  adolescence | 0.040 | | -0.002 | 0.000 | 0.000 | 0.000 | 0.014 | 0.000 | 0.011 | 0.070 | 0.000 | 0.000 | 0.000 | 0.083 | 0.000 | 0.000 |
| Externalizing  adulthood | 0.082 | | 0.000 | 0.000 | -0.044 | 0.000 | 0.020 | 0.000 | 0.045 | 0.043 | 0.000 | 0.000 | 0.000 | 0.048 | 0.042 | -0.001 |
| Internalizing  childhood | 0.031 | | 0.000 | 0.007 | 0.000 | 0.000 | 0.000 | 0.000 | 0.000 | 0.000 | 0.056 | 0.000 | 0.000 | 0.000 | 0.000 | 0.000 |
| Internalizing  adolescence | 0.036 | | -0.036 | 0.018 | -0.012 | 0.000 | 0.014 | -0.041 | 0.040 | 0.000 | 0.123 | 0.000 | 0.027 | 0.021 | 0.000 | -0.008 |
| Internalizing  adulthood | 0.027 | | 0.000 | 0.021 | -0.007 | 0.000 | 0.001 | 0.000 | 0.041 | 0.043 | 0.015 | 0.000 | 0.000 | 0.000 | 0.000 | 0.000 |
| **Cross-age-**  **and-rater**  **(cross-age**  **approach)** |  | |  |  |  |  |  |  |  |  |  |  |  |  |  |  |
| BPp | 0.097 | | -0.025 | 0.027 | -0.043 | 0.011 | 0.000 | -0.026 | 0.038 | 0.048 | 0.047 | 0.015 | 0.003 | 0.029 | -0.001 | -0.001 |
| Externalizing | 0.129 | | 0.000 | 0.000 | -0.047 | 0.000 | 0.010 | 0.000 | 0.017 | 0.044 | 0.000 | 0.001 | 0.000 | 0.046 | 0.000 | 0.000 |
| Internalizing | 0.066 | | 0.000 | 0.000 | -0.015 | 0.000 | 0.002 | 0.000 | 0.026 | 0.011 | 0.074 | 0.000 | 0.000 | 0.000 | 0.000 | 0.000 |
| **Single-trait**  **cross-age** |  | |  |  |  |  |  |  |  |  |  |  |  |  |  |  |
| Hyperactivity  parent-rated | 0.104 | | 0.000 | 0.000 | 0.000 | 0.000 | 0.000 | 0.000 | 0.000 | 0.000 | 0.000 | 0.000 | 0.000 | 0.007 | 0.000 | 0.000 |
| Hyperactivity  teacher-rated | 0.138 | | 0.000 | 0.005 | -0.017 | 0.000 | 0.000 | -0.012 | -0.020 | 0.000 | 0.000 | 0.005 | 0.019 | 0.051 | 0.000 | 0.015 |
| Hyperactivity  child-rated | 0.071 | | 0.000 | 0.000 | 0.000 | 0.000 | 0.000 | 0.000 | 0.004 | 0.000 | 0.000 | 0.000 | 0.000 | 0.033 | 0.000 | 0.000 |
| Conduct  parent-rated | 0.107 | | -0.024 | 0.016 | -0.017 | 0.007 | 0.010 | 0.000 | 0.037 | 0.037 | 0.000 | 0.000 | -0.022 | 0.017 | 0.023 | -0.024 |
| Conduct  teacher-rated | 0.081 | | 0.000 | 0.000 | 0.000 | 0.009 | 0.000 | 0.000 | 0.000 | 0.000 | 0.000 | 0.000 | -0.006 | 0.054 | 0.000 | 0.000 |
| Conduct  child-rated | 0.039 | | 0.000 | -0.002 | 0.000 | 0.000 | 0.013 | 0.000 | 0.000 | 0.038 | 0.000 | 0.000 | 0.000 | 0.095 | 0.000 | -0.045 |
| Emotional  problems  parent-rated | 0.009 | | 0.000 | 0.000 | -0.004 | 0.000 | 0.000 | -0.026 | 0.024 | 0.028 | 0.077 | 0.000 | 0.002 | -0.016 | 0.000 | 0.000 |
| Emotional  problems  teacher-rated | 0.006 | | 0.000 | 0.000 | -0.010 | 0.000 | 0.000 | -0.039 | 0.013 | 0.033 | 0.052 | 0.005 | 0.000 | -0.024 | 0.017 | 0.000 |
| Emotional  problems  child-rated | 0.021 | | -0.016 | 0.002 | -0.008 | 0.000 | -0.001 | -0.022 | 0.051 | 0.003 | 0.083 | 0.006 | 0.000 | 0.019 | 0.024 | -0.019 |
| Peer problems  parent-rated | 0.013 | | 0.000 | 0.033 | -0.001 | 0.000 | 0.000 | 0.000 | 0.016 | 0.000 | 0.025 | 0.000 | 0.002 | 0.000 | 0.000 | 0.000 |
| Peer problems  teacher-rated | 0.010 | | 0.000 | 0.023 | 0.000 | 0.000 | 0.000 | 0.000 | 0.000 | 0.000 | 0.000 | 0.000 | 0.000 | 0.000 | 0.000 | 0.000 |
| Peer problems  child-rated | 0.032 | | -0.010 | 0.007 | 0.000 | 0.000 | 0.000 | 0.000 | 0.010 | 0.020 | 0.031 | 0.000 | 0.000 | 0.000 | -0.041 | -0.007 |
| **Single-trait**  **cross-rater** |  | |  |  |  |  |  |  |  |  |  |  |  |  |  |  |
| Hyperactivity  childhood | 0.112 | | -0.008 | -0.003 | -0.007 | 0.000 | 0.025 | -0.031 | 0.000 | 0.030 | 0.000 | 0.034 | 0.026 | 0.029 | -0.031 | 0.001 |
| Hyperactivity  adolescence | 0.043 | | 0.000 | 0.000 | 0.000 | 0.000 | 0.000 | 0.000 | 0.000 | 0.057 | 0.000 | 0.000 | 0.000 | 0.039 | 0.009 | 0.000 |
| Hyperactivity  adulthood | 0.064 | | 0.021 | 0.000 | -0.006 | -0.001 | 0.000 | 0.000 | 0.000 | 0.022 | 0.000 | 0.000 | 0.000 | 0.037 | 0.024 | 0.000 |
| Conduct  childhood | 0.077 | | 0.000 | 0.027 | 0.000 | 0.028 | 0.000 | 0.000 | 0.012 | 0.001 | 0.000 | 0.000 | 0.000 | 0.000 | 0.000 | 0.000 |
| Conduct  adolescence | 0.060 | | -0.003 | -0.017 | -0.021 | 0.008 | 0.012 | 0.000 | 0.021 | 0.042 | 0.015 | 0.000 | 0.000 | 0.077 | 0.014 | -0.025 |
| Conduct  adulthood | 0.069 | | 0.000 | 0.000 | 0.000 | 0.000 | 0.008 | 0.001 | 0.036 | 0.017 | 0.000 | 0.000 | 0.000 | 0.031 | 0.000 | 0.000 |
| Emotional  problems childhood | 0.000 | | 0.000 | 0.000 | 0.000 | 0.000 | 0.000 | 0.000 | 0.000 | 0.000 | 0.066 | 0.000 | 0.000 | -0.012 | 0.000 | 0.000 |
| Emotional  problems adolescence | 0.004 | | -0.015 | 0.000 | 0.000 | 0.000 | 0.014 | -0.031 | 0.029 | 0.023 | 0.115 | 0.000 | 0.000 | 0.000 | 0.000 | 0.000 |
| Emotional  problems adulthood | 0.000 | | 0.000 | 0.001 | 0.000 | 0.000 | 0.000 | 0.000 | 0.073 | 0.014 | 0.060 | 0.000 | 0.000 | 0.000 | 0.000 | 0.000 |
| Peer problems  childhood | 0.007 | | 0.000 | 0.029 | -0.008 | 0.000 | 0.000 | 0.000 | 0.000 | 0.000 | 0.013 | 0.000 | 0.000 | 0.000 | 0.000 | 0.000 |
| Peer problems  adolescence | 0.031 | | -0.017 | 0.017 | -0.002 | 0.000 | 0.000 | 0.000 | 0.008 | 0.000 | 0.023 | 0.000 | 0.004 | 0.000 | 0.000 | -0.001 |
| Peer problems  adulthood | 0.031 | | 0.000 | 0.000 | 0.000 | 0.006 | 0.020 | 0.000 | 0.023 | 0.039 | 0.000 | 0.000 | 0.000 | -0.039 | 0.000 | 0.000 |

| **Supporting table S10**. Cross-age and cross-rater behaviour problems composites: bivariate twin model fitting results. | | | | | | | | | | |
| --- | --- | --- | --- | --- | --- | --- | --- | --- | --- | --- |
| **Note**. | | | | | | | | | | |
| Est= estimate; bivA= genetic correlation; bivC= shared environmental correlation; bivE= unique environmental correlation; CI= confidence interval. | | | | | | | | | | |
| **Composite 1** | **Composite 2** | **Bivariate twin model fitting results** | | | | | | | | |
|  |  | **bivA** | | | **bivC** | | | **bivE** | | |
|  |  | Est | Lower 95% CI | Upper 95% CI | Est | Lower 95% CI | Upper 95% CI | Est | Lower 95% CI | Upper 95% CI |
| **Cross-age** | **Cross-rater** |  |  |  |  |  |  |  |  |  |
| BPp parent-rated | BPp childhood | 0.943 | 0.933 | 0.952 | 0.992 | 0.966 | 1.000 | 0.836 | 0.821 | 0.849 |
| BPp parent-rated | BPp adolescence | 0.717 | 0.669 | 0.765 | 0.840 | 0.840 | 0.996 | 0.379 | 0.338 | 0.418 |
| BPp parent-rated | BPp adulthood | 0.664 | 0.602 | 0.727 | 0.880 | 0.675 | 1.000 | 0.337 | 0.292 | 0.381 |
| BPp teacher-rated | BPp childhood | 0.662 | 0.620 | 0.702 | -1.000 | -1.000 | 1.000 | 0.448 | 0.409 | 0.485 |
| BPp teacher-rated | BPp adolescence | 0.500 | 0.438 | 0.565 | 1.000 | 0.994 | 1.000 | 0.225 | 0.179 | 0.270 |
| BPp teacher-rated | BPp adulthood | 0.408 | 0.329 | 0.498 | 0.999 | -1.000 | NA | 0.039 | -0.012 | 0.091 |
| BPp child-rated | BPp childhood | 0.504 | 0.425 | 0.584 | 0.682 | 0.341 | 1.000 | 0.110 | 0.064 | 0.156 |
| BPp child-rated | BPp adolescence | 0.860 | 0.823 | 0.896 | 0.951 | 0.784 | 1.000 | 0.744 | 0.723 | 0.764 |
| BPp child-rated | BPp adulthood | 0.715 | 0.664 | 0.760 | 1.000 | 0.777 | 1.000 | 0.644 | 0.616 | 0.671 |
| Externalizing parent-rated | Externalizing childhood | 0.960 | 0.955 | 0.965 | 1.000 | 0.943 | 1.000 | 0.900 | 0.890 | 0.908 |
| Externalizing parent-rated | Externalizing adolescence | 0.722 | 0.700 | 0.748 | 1.000 | -1.000 | 1.000 | 0.292 | 0.250 | 0.333 |
| Externalizing parent-rated | Externalizing adulthood | 0.683 | 0.651 | 0.725 | 1.000 | 0.909 | 1.000 | 0.277 | 0.232 | 0.322 |
| Externalizing teacher-rated | Externalizing childhood | 0.621 | 0.586 | 0.657 | -1.000 | -1.000 | 1.000 | 0.430 | 0.390 | 0.469 |
| Externalizing teacher-rated | Externalizing adolescence | 0.447 | 0.407 | 0.493 | 1.000 | -1.000 | 1.000 | 0.169 | 0.123 | 0.214 |
| Externalizing teacher-rated | Externalizing adulthood | 0.399 | 0.337 | 0.467 | 0.371 | -1.000 | 1.000 | 0.038 | -0.013 | 0.088 |
| Externalizing child-rated | Externalizing childhood | 0.515 | 0.480 | 0.564 | 1.000 | -1.000 | 1.000 | 0.091 | 0.046 | 0.135 |
| Externalizing child-rated | Externalizing adolescence | 0.866 | 0.835 | 0.898 | -0.994 | -1.000 | 1.000 | 0.751 | 0.731 | 0.771 |
| Externalizing child-rated | Externalizing adulthood | 0.719 | 0.667 | 0.772 | 1.000 | -1.000 | 1.000 | 0.624 | 0.594 | 0.652 |
| Internalizing parent-rated | Internalizing childhood | 0.902 | 0.886 | 0.919 | 0.996 | 0.973 | 1.000 | 0.811 | 0.795 | 0.827 |
| Internalizing parent-rated | Internalizing adolescence | 0.704 | 0.644 | 0.761 | 0.836 | 0.716 | 0.998 | 0.424 | 0.385 | 0.462 |
| Internalizing parent-rated | Internalizing adulthood | 0.624 | 0.554 | 0.700 | 0.980 | 0.754 | 1.000 | 0.339 | 0.293 | 0.383 |
| Internalizing teacher-rated | Internalizing childhood | 0.575 | 0.513 | 0.633 | 0.595 | 0.174 | 1.000 | 0.408 | 0.368 | 0.447 |
| Internalizing teacher-rated | Internalizing adolescence | 0.539 | 0.465 | 0.615 | 0.865 | 0.581 | 1.000 | 0.280 | 0.236 | 0.324 |
| Internalizing teacher-rated | Internalizing adulthood | 0.393 | 0.303 | 0.499 | 0.703 | -1.000 | 1.000 | 0.049 | -0.002 | 0.100 |
| Internalizing child-rated | Internalizing childhood | 0.473 | 0.380 | 0.568 | 0.685 | 0.430 | 1.000 | 0.127 | 0.081 | 0.172 |
| Internalizing child-rated | Internalizing adolescence | 0.835 | 0.794 | 0.878 | 0.964 | 0.802 | 1.000 | 0.714 | 0.690 | 0.735 |
| Internalizing child-rated | Internalizing adulthood | 0.737 | 0.685 | 0.786 | 1.000 | 0.916 | 1.000 | 0.646 | 0.619 | 0.672 |

***Supporting references***

Allegrini, A. G., Karhunen, V., Coleman, J. R., Selzam, S., Rimfeld, K., von Stumm, S., ... & Plomin, R. (2020). Multivariable GE interplay in the prediction of educational achievement. *PLoS genetics*, *16*(11), e1009153.

Behar, L. B. (1977): The Preschool Behavior Questionnaire; Journal of Abnormal Child Psychology, 5, 265–275.

Cheesman, R., Purves, K. L., Pingault, J. B., Breen, G., Plomin, R., & Eley, T. C. (2018). Extracting stability increases the SNP heritability of emotional problems in young people. *Translational psychiatry*, *8*(1), 1-9.

Demontis, D., Walters, R. K., Martin, J., Mattheisen, M., Als, T. D., Agerbo, E., ... & Cerrato, F. (2019). Discovery of the first genome-wide significant risk loci for attention deficit/hyperactivity disorder. *Nature Genetics*, *51*(1), 63-75.

Duncan, L. E., Ratanatharathorn, A., Aiello, A. E., Almli, L. M., Amstadter, A. B., Ashley-Koch, A. E., ... & Bradley, B. (2018). Largest GWAS of PTSD (N= 20 070) yields genetic overlap with schizophrenia and sex differences in heritability. *Molecular Psychiatry*, *23*(3), 666-673.

Fushiki, T. (2011). Estimation of prediction error by using K-fold cross-validation. *Statistics and Computing*, *21*(2), 137-146.

Genetics IOCDF, Arnold, P. D., Askland, K. D., Barlassina, C., Bellodi, L., Bienvenu, O. J., Black, D., ... & Cappi, C. (2018). Revealing the complex genetic architecture of obsessive-compulsive disorder using meta-analysis. *Molecular Psychiatry*, *23*(5), 1181-1181.

Goodman, R. (1997): The Strengths and Difficulties Questionnaire: a research note; Journal of Child Psychology and Psychiatry, 38, 581-586.

Grove, J., Ripke, S., Als, T. D., Mattheisen, M., Walters, R. K., Won, H., ... & Awashti, S. (2019). Identification of common genetic risk variants for autism spectrum disorder. *Nature Genetics*, *51*(3), 431-444.

Hastie, T., & Qian, J. (2014). Glmnet vignette. *Retrieved June*, *9*(2016), 1-30.

Hemani, G., & Yang, J. (2017). Gcta-greml power calculator.

Howard, D. M., Adams, M. J., Clarke, T. K., Hafferty, J. D., Gibson, J., Shirali, M., ... & Alloza, C. (2019). Genome-wide meta-analysis of depression identifies 102 independent variants and highlights the importance of the prefrontal brain regions. *Nature Neuroscience*, *22*(3), 343-352.

Jansen, P. R., Watanabe, K., Stringer, S., Skene, N., Bryois, J., Hammerschlag, A. R., ... & Savage, J. E. (2019). Genome-wide analysis of insomnia in 1,331,010 individuals identifies new risk loci and functional pathways. *Nature Genetics*, *51*(3), 394-403.

Kuhn, M. (2012). The caret package. *R Foundation for Statistical Computing, Vienna, Austria. URL https://cran. r-project. org/package= caret*

Linnér, R. K., Biroli, P., Kong, E., Meddens, S. F. W., Wedow, R., Fontana, M. A., ... & Nivard, M. G. (2019). Genome-wide association analyses of risk tolerance and risky behaviours in over 1 million individuals identify hundreds of loci and shared genetic influences. *Nature Genetics*, *51*(2), 245-257.

Luciano, M., Hagenaars, S. P., Davies, G., Hill, W. D., Clarke, T. K., Shirali, M., ... & Adams, M. J. (2018). Association analysis in over 329,000 individuals identifies 116 independent variants influencing neuroticism. *Nature Genetics*, *50*(1), 6-11.

McGue, M., & Bouchard, T. J. (1984). Adjustment of twin data for the effects of age and sex. Behavior genetics, 14(4), 325-343.

Neale, M. C., Hunter, M. D., Pritikin, J. N., Zahery, M., Brick, T. R., Kirkpatrick, R. M., ... & Boker, S. M. (2016). OpenMx 2.0: Extended structural equation and statistical modeling. *Psychometrika*, *81*(2), 535-549.

Okbay, A., Baselmans, B. M., De Neve, J. E., Turley, P., Nivard, M. G., Fontana, M. A., ... & Gratten, J. (2016). Genetic variants associated with subjective well-being, depressive symptoms, and neuroticism identified through genome-wide analyses. *Nature Genetics*, *48*(6), 624-633.

Pardiñas, A. F., Holmans, P., Pocklington, A. J., Escott-Price, V., Ripke, S., Carrera, N., ... & Han, J. (2018). Common schizophrenia alleles are enriched in mutation-intolerant genes and in regions under strong background selection. *Nature Genetics*, *50*(3), 381-389.

Pavlou, M., Ambler, G., Seaman, S., De Iorio, M., & Omar, R. Z. (2016). Review and evaluation of penalised regression methods for risk prediction in low‐dimensional data with few events. *Statistics in medicine*, *35*(7), 1159-1177.

Price, T. S., Freeman, B., Craig, I., Petrill, S. A., Ebersole, L., & Plomin, R. (2000). Infant zygosity can be assigned by parental report questionnaire data. Twin Research and Human Genetics, 3(3), 129-133.

R Core Team (2020). R: A language and environment for statistical computing. R Foundation for Statistical Computing, Vienna, Austria. URL <https://www.R-project.org/>.

Revelle, W. (2020) psych: Procedures for Personality and Psychological Research, Northwestern University, Evanston, Illinois, USA, <https://CRAN.R-project.org/package=psych> Version = 2.0.9,.

Rimfeld, K., Malanchini, M., Spargo, T., Spickernell, G., Selzam, S., McMillan, A., ... & Plomin, R. (2019). Twins early development study: A genetically sensitive investigation into behavioural and cognitive development from infancy to emerging adulthood. *Twin Research and Human Genetics*, *22*(6), 508-513.

Rosseel, Y. (2012). Lavaan: An R package for structural equation modeling and more. Version 0.5–12 (BETA). *Journal of statistical software*, *48*(2), 1-36.

Seed, C. (2017). Hail: An Open-Source Framework for Scalable Genetic Data. *Neale Lab http://www. nealelab. is/blog/2017/7/19/rapid-gwas-of-thousands-of-phenotypes-for-337000-samples-in-the-uk-biobank*.

Stahl, E. A., Breen, G., Forstner, A. J., McQuillin, A., Ripke, S., Trubetskoy, V., ... & de Leeuw, C. A. (2019). Genome-wide association study identifies 30 loci associated with bipolar disorder. *Nature Genetics*, *51*(5), 793-803.

Viechtbauer, W., & Viechtbauer, M. W. (2015). Package ‘metafor’. *The Comprehensive R Archive Network. Package ‘metafor’. http://cran. r-project. org/web/packages/metafor/metafor. pdf*.

Visscher, P. M., Hemani, G., Vinkhuyzen, A. A., Chen, G. B., Lee, S. H., Wray, N. R., ... & Yang, J. (2014). Statistical power to detect genetic (co) variance of complex traits using SNP data in unrelated samples. PLoS Genet, 10(4), e1004269.

Watson, H. J., Yilmaz, Z., Thornton, L. M., Hübel, C., Coleman, J. R., Gaspar, H. A., ... & Medland, S. E. (2019). Genome-wide association study identifies eight risk loci and implicates metabo-psychiatric origins for anorexia nervosa. *Nature Genetics*, *51*(8), 1207-1214.

Wray, N. R., Ripke, S., Mattheisen, M., Trzaskowski, M., Byrne, E. M., Abdellaoui, A., ... & Bacanu, S. A. (2018). Genome-wide association analyses identify 44 risk variants and refine the genetic architecture of major depression. *Nature Genetics*, *50*(5), 668-681.

Zou, H., & Hastie, T. (2005). Regularization and variable selection via the elastic net *Journal of the Royal Statistical Society: Series B (Statistical Methodology)*, *67*(2), 301-320.
